# The global randomization test: A Mendelian randomization falsification test for the exclusion restriction assumption

**DOI:** 10.1101/2022.05.03.22274459

**Authors:** Louise AC Millard, George Davey Smith, Kate Tilling

**Affiliations:** MRC Integrative Epidemiology Unit (IEU) at the University of Bristol, Bristol, United Kingdom; Population health Sciences, Bristol Medical School, University of Bristol, Bristol, United Kingdom

**Keywords:** Mendelian randomization, exclusion restriction assumption, falsification test, selection bias, horizontal pleiotropy

## Abstract

Mendelian randomization may give biased causal estimates if the instrument affects the outcome not solely via the exposure of interest (violating the exclusion restriction assumption). We demonstrate use of a global randomization test as a falsification test for the exclusion restriction assumption. Using simulations, we explored the statistical power of the randomization test to detect an association between a genetic instrument and a covariate set due to a) selection bias or b) horizontal pleiotropy, compared to three approaches examining associations with individual covariates: i) Bonferroni correction for the number of covariates, and ii) correction for the effective number of independent covariates and iii) an r^2^ permutation-based approach. We conducted proof-of-principle analyses in UK Biobank, using CRP as the exposure and coronary heart disease (CHD) as the outcome. In simulations, power of the randomization test was higher than the other approaches for detecting selection bias when the correlation between the covariates was low (R^2^< 0.1), and at least as powerful as the other approaches across all simulated horizontal pleiotropy scenarios. In our applied example, we found strong evidence of selection bias using all approaches (e.g., global randomization test p<0.002). We identified 51 of the 58 CRP genetic variants as horizontally pleiotropic, and estimated effects of CRP on CHD attenuated somewhat to the null when excluding these from the genetic risk score (OR=0.956 [95% CI: 0.918, 0.996] versus 0.970 [95% CI: 0.900, 1.046] per 1-unit higher log CRP levels). The global randomization test can be a useful addition to the MR researcher’ s toolkit.

## INTRODUCTION

Mendelian randomization (MR) is a valuable approach to test for causal effects using observational data, generally using a genetic instrumental variable (IV) to proxy for the exposure of interest [1–4]. However, three core assumptions need to be made, to be able to test for a causal effect using MR, and violations of these assumptions may bias results [5]. These three assumptions are: 1) the IV is associated with the exposure (relevance assumption), 2) there is no unmeasured (i.e., unaccounted for) confounding between the IV and the outcome (independence assumption) and 3) the association of the IV and the outcome is entirely via the exposure (exclusion restriction assumption). To estimate the magnitude of (not just test for) an effect a further assumption of monotonicity or homogeneity is required [4,6]. The independence assumption may be violated by confounding due to population stratification, by dynastic effects and assortative mating [7]. The exclusion restriction assumption may be violated due to horizontal pleiotropy, where the genetic variant affects the outcome along pathways that are not via the exposure, or linkage disequilibrium. Selection bias can also violate the exclusion restriction assumption by inducing a pathway between the IV and confounders through conditioning on a collider [8].

While only the relevance assumption can be directly tested (by testing the strength of the association of the exposure with the IV), the independence and exclusion restriction assumptions can be investigated with sensitivity analyses and falsification tests that test for evidence that these assumptions do not hold. A common falsification test for the independence assumption is to test for covariate prevalence difference (also known as covariate balance), by testing the association of the IV with a set of potential confounders. Provided these factors are not on the causal path between the IV and exposure, or the exposure and outcome, the IV should not be associated with these factors if the independence assumption holds [9]. For example, cis CRP genetic variants were not found to be related to risk factors for cardiovascular disease [10,11]. Bias can also be estimated, as the covariate prevalence difference divided by the exposure prevalence difference, and displayed in confounding bias plots [9,12]. This is useful when a researcher wants to compare the potential bias due to confounding in an IV analysis with that of a conventional multivariable regression, as the bias in the causal MR estimate depends also on the strength of the effect of the IV on the exposure [9]. For example, confounding bias plots have been used to assess the potential bias in IV studies of myopia [13] and education [13,14].

A recent study proposed an approach to compare balance or bias of an IV analysis with what would be expected from a randomized experiment [15]. Given a set of covariates, C, their approach – which we refer to as the global randomization test – uses permutation testing to test whether a binary instrument Z is as-if randomized according to p(Z|C), by comparing the observed test statistic (e.g., covariate bias or balance) with that which we would expect if this were true (i.e. no difference in C across values of Z). They suggest the Mahalanobis distance can be used as a global measure of balance and bias, across the set of covariates tested. In their study [15] they assume that C are measured before Z and X are assigned, hence assume that there is no alternate path between the IV and outcome via C rather than X (i.e. the exclusion restriction assumption holds). However, in an MR setting with a genetic IV, an association between Z and a covariate C may be because of violations of either the independence or the exclusion restriction assumptions (or both) (see Figure 1). Thus, in MR studies the randomization test has potential to be used as a falsification test for both these assumptions, depending on the MR analysis in question.

**Figure 1:**
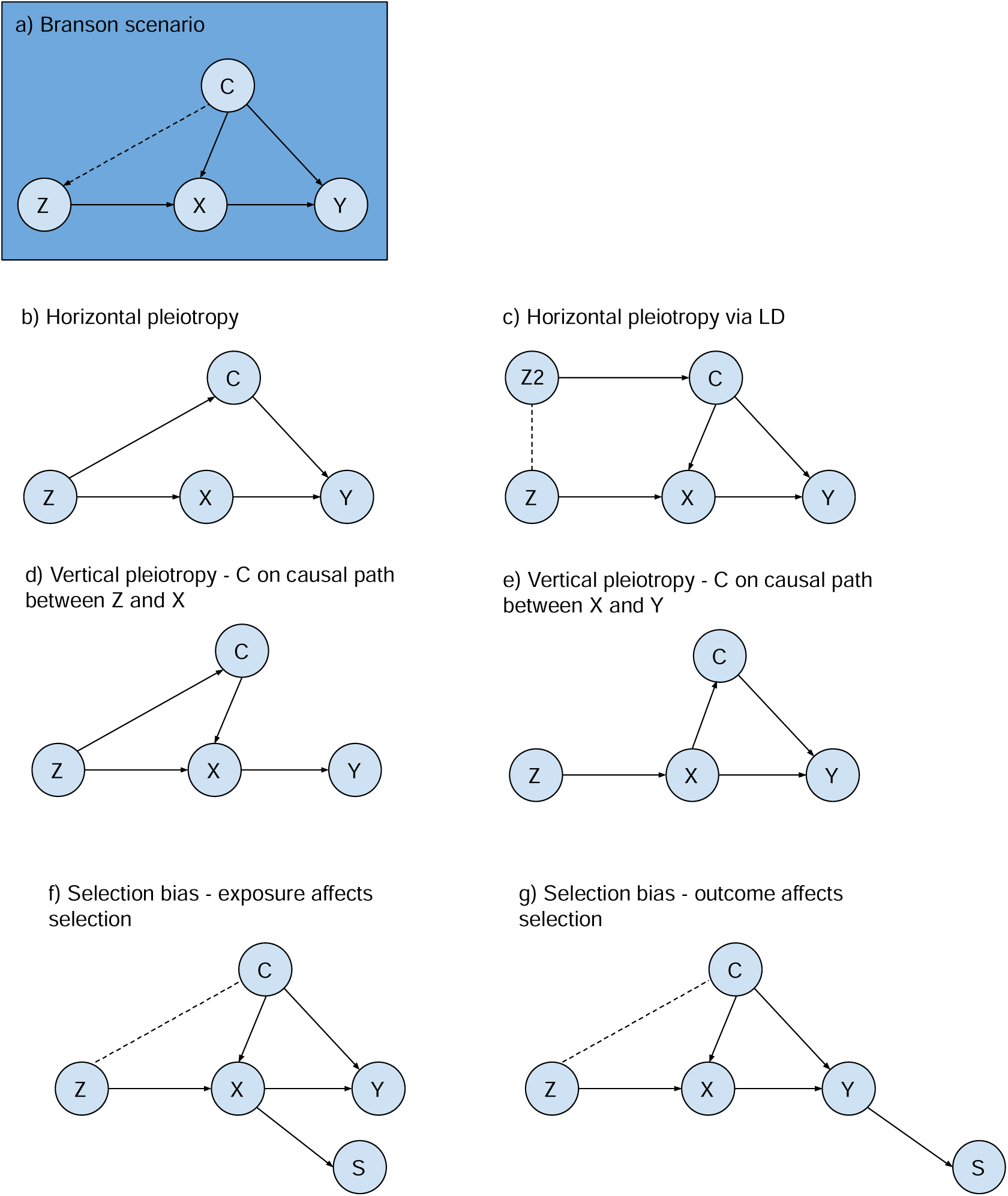
Example scenarios where covariate imbalance may be seen. Z: instrumental variable; X: exposure; Y: outcome; C: covariates; S: selection. Branson scenario (a) assumes covariates C are measured prior to Z and X and may affect both X and Y (dashed arrow indicates association tested using the randomization test). b-g) Example MR scenarios through which covariate imbalance can occur. In figure (b) dashed line indicates linkage disequilibrium (LD) between genetic variants). Figures (f) and (g) show two example scenarios through which covariates C become correlated with instrument Z due to selection bias [29].

In this paper we show how the global randomization test can be useful in MR studies, to identify potential bias due to horizontal pleiotropy or non-random selection. We demonstrate the statistical power in these scenarios using simulations and demonstrate how this approach can be used in practice using proof of principle applications.

## METHODS

### Overview of the randomization test procedure

The global randomization test approach as presented in [15] has the following steps:

1. Define set of covariates to test – this depends on the specific scenario (see applied examples).
2. Calculate the test statistic *T*, the Mahalanobis distance (with values [0,∞, which is a global measure of balance and bias across all covariates tested.
3. Permute the genetic IV N_p_ times and for each calculate the test statistic t, where N_p_ is specified by the researcher. We use N_p_ =5000 in our simulations and applied examples below.
4. Calculate the P value as the proportion of permutations with a test statistic *t* at least as strong as *T, i*.*e*. |*t* ≥ *T*:|/N_*p*_.

We generalize the approach in [15] (that focused on binary IVs) to continuous, ordinal and binary IVs, as described in the following section.

*Generalising the Mahalanobis distance to allow continuous, ordered categorical and binary variables* We use the Mahalanobis distance as a global measure of balance defined for an IV with two categories as:

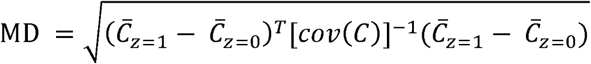

Where C is a *m*× *n* matrix of *m* participants and *n* covariates, and 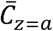 is a vector containing the mean of the covariates for the subset of participants where z=a.

Since MD is affinely invariant, this is also a global measure of bias (i.e. changing prevalence difference 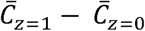 to bias measure 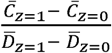 in the above equation would result in the same MD value).

To generalize to IVs with three categories (i.e., SNP dosages) and continuous IVs (i.e., genetic risk scores), we generalize this equation to:

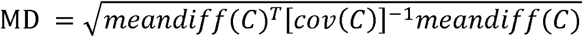

Where *meandiff(C)* is a 1xn vector of the mean difference of each covariate per 1 unit higher IV. This assumes a linear relationship between the IV and covariates.

We estimate the mean difference using the correlation between z and each covariate C_*i*_ :

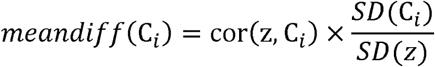

where *cor(A, B)* is the Pearson’ s r2 between A and B, SD(X) is the standard deviation of X. This approach also assumes a linear relationship between the IV and covariates, but is ∼15 times faster than estimating the mean difference using linear regression (particularly helpful for this work as the randomization test uses permutation testing and we conduct extensive simulations). Note that the MD is invariant to the scale of covariates in C, but not invariant to the scale of Z, and the resultant global randomization test P value is independent of both.

### Simulations

We conduct simulations to explore scenarios where the global randomization test may be useful when testing for horizontal pleiotropy and selection bias. We conduct separate simulations to test for selection bias and horizontal pleiotropy, illustrating how the covariates can be chosen to test each scenario. We report the aims, data-generating mechanisms, estimands, methods, and performance measures of our simulations (the ADEMP approach) [16].

#### Simulation A: Assessing statistical power to detect potential selection bias

This simulation is based on the situation where a researcher wants to determine whether the GRS relates to covariates that are unlikely to be downstream effects of the GRS, such that an association would indicate possible selection bias.

##### Aim

To compare statistical power of the global randomization test compared to alternative tests that test each covariate individually, across different (1) number of covariates that affect selection, 2) number of covariates than do not affect selection, and 3) correlations between covariates.

##### Data generating mechanism

The directed acyclic graph (DAG) on which our data generating mechanism is based is shown in Figure 2a. We set the proportion selected to 5.5%, based on the UK Biobank recruitment rate where 9.2 million invited and 5.5% of those joined the study. We use a sample size of 920,000, 10% of the number invited in UK Biobank to keep the simulation manageable [17]. A set of covariates C including those affecting selection C_*s*_ and those not affecting selection 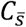 are included. We vary the number of covariates affecting selection *N*_*cs*_, the number of covariates not affecting selection 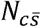, the correlation between all variables in C, 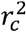 the variance of X explained by Z, 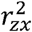, and the pseudo variance of S explained by C_*s*_ and X, 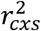. We use a fully factorial design, running our simulation with all combinations of the following values: 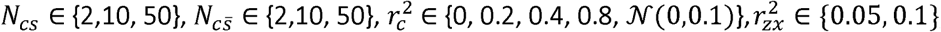, and 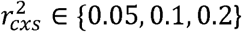. For the 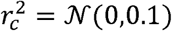 setting the covariate correlations are generated from a normal distribution with mean=0 and standard deviation (SD)=0.1, to reflect correlations reported previously [18]. All covariates in C, and exposure X are continuous with mean=0 and SD=1. Instrument Z is assumed to be a normally distributed genetic risk score with mean=0 and SD=1.

**Figure 2:**
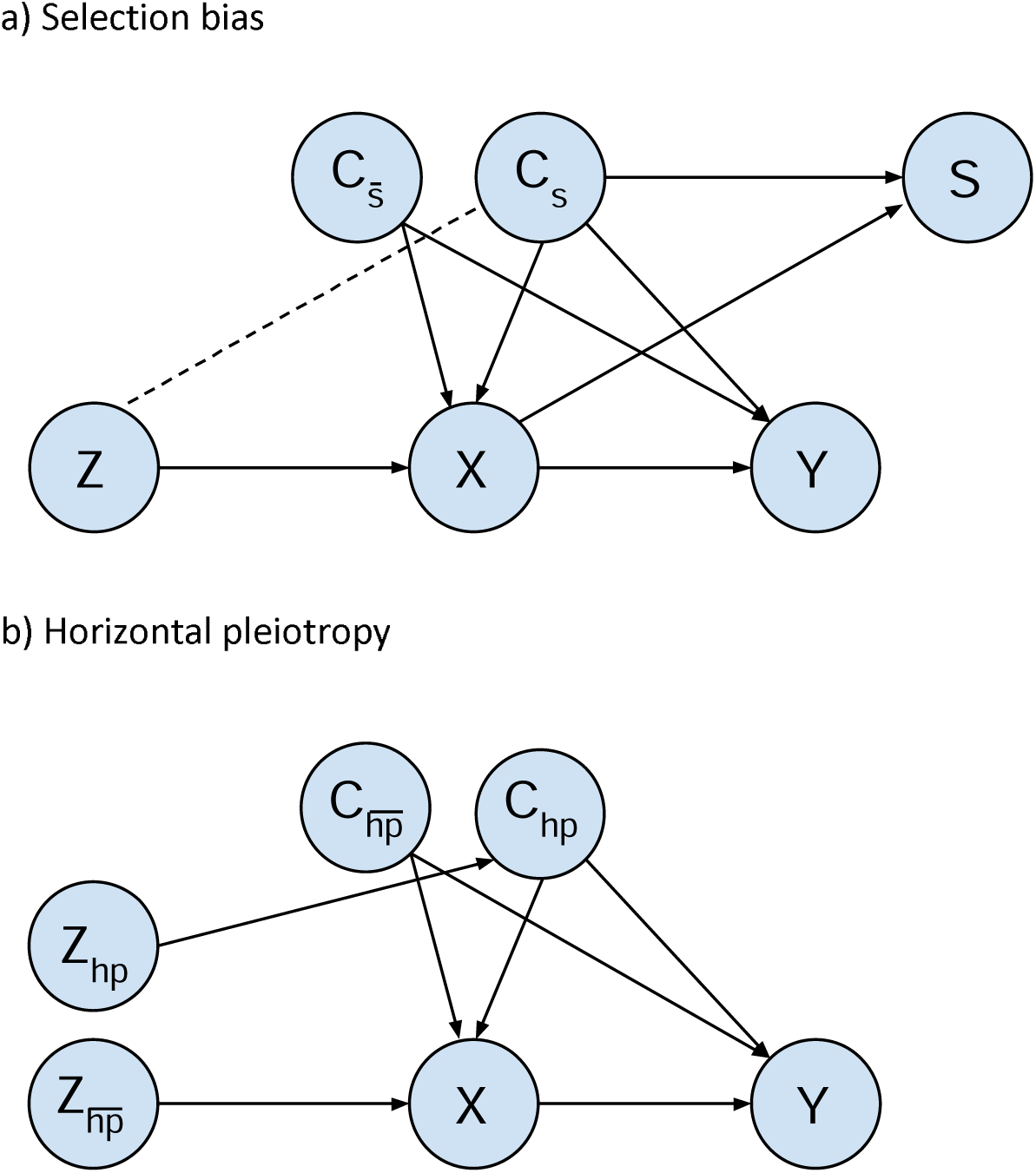
DAGs for simulation data generating mechanisms. DAG (a): Covariates *c*_*s*_ and 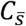 are confounders of X and Y. Covariates *c*_*s*_ and exposure X affect selection (S) inducing an association between instrument Z and *c*_*s*_. X, *c*_*s*_ and 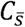 may affect Y but effects on Y do not impact associations between Z and *c*_*s*_ tested by global randomization test. The total effect of the following paths on the DAG is kept constant irrespective of the number of covariates in *c*_*s*_ and 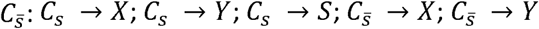. Dashed line indicated a statistical association induced through conditioning on *S*. DAG (b): Covariates *c*_*hp*_ and 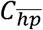 are confounders of X and Y. In this DAG we depict a horizontally pleiotropic instrument that affects Y both via and not via X. While this isn’ t necessarily the case (i.e. Z_h_p might affect Y via X only, or directly (i.e. not via X)) here we are showing an example – the exact relationship between the instruments and X and Y doesn’ t impact the randomization test because the randomization test only tests the association between each instrument and the covariate set, and X the relationships with X and Y does not impact the strength of this association (unlike in the selection bias example where e.g. the effect of Z on X impacts the magnitude of the selection inducted association between Z and the covariate set).

We fix the total effect of the following relationships. For variable X, we fixed the total effect of all covariates C_*s*_ and 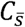 on X, to r^2^=0.1, with equal contribution by each set (i.e., r^2^=0.05 for each set irrespective of N*cs* and 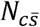). This fixes the strength of effect of the covariates on selection indirectly via X. Further details of this process are given in Supplementary section S1. A DAG with simulation parameters is shown in Supplementary figure 1a. The outcome Y is not modelled as, according to our DAG in Figure 2a, the association between the SNPs and covariates induced by conditioning on selection does not depend on our definition of Y.

##### Estimand or other target

Our target is the test of the null hypothesis of no association between the GRS and covariate set.

##### Methods

We compare the global randomization test with 3 alternative approaches to test the association of the IV with C:

a. individual tests of each covariate with Bonferroni correction, referred to as test-Bonf.
b. individual tests of each covariate with correction for the effective number of tests performed, referred to as test-indep.
c. permutation testing where the test statistic is the maximum r^2^ of each of the covariates with the IV, referred to as test-r^2^perm.

To calculate (a) and (b) we first regress the IV on each of the covariates using univariable regression, and find the lowest p value of these results, *pvalue*_*min*_. The test-Bonf p value is then calculated as Min(1, *pvalue*_*min*_ x N_c_). The p value for test-indep in calculated as Min(1, *pvalue*_*min*_ x N_*i*_) N_*i*_, where NI is the estimated effective number of independent tests calculated using spectral decomposition [19,20].

##### Performance measures

We evaluate statistical power using rejection percentage [16], which is the proportion of simulation repetitions, *n*_*sim*_, where the null hypothesis is rejected (see further details in Supplementary section S2). We set *n*_*sim*_ =500 in all our simulations. The four tests (global randomization test, test-Bonf, and test-indep and test-r^2^perm) are applied to the same simulated dataset in each simulation repetition.

We repeated these simulations, including only half the covariates in *C*_*s*_ and 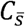 to represent the scenario where only a subset of these covariates is either available or hypothesized to affect selection. We also repeated these simulations in the whole sample (I.e., with no selection) to check that we observe ∼5% type-1 error, i.e., around 5% of the permutations incorrectly identify an association between the IV and covariate set.

#### Simulation B: Assessing statistical power to detect potential horizontal pleiotropy

This simulation is based on the situation where a researcher has a GRS but wants to determine whether any of the SNPs included may affect the outcome via horizontally pleiotropic pathways rather than (solely) via the exposure of interest.

##### Aim

To compare statistical power of the global randomization test with alternatives that test the association of a SNP with each covariate individually, to identify whether the SNP acts via a horizontally pleiotropic pathway. We evaluate this across different: 1) numbers of covariates affected and not affected by a horizontally pleiotropic SNP, and 2) magnitude of effect of a horizontally pleiotropic SNP on covariates on the horizontal pleiotropy pathway.

##### Data generating mechanisms

The DAG on which our DGM is based is shown in Figure 2b. We use a sample size of 500,000 reflecting the size of UK Biobank. We include one horizontally pleiotropic SNP,

*Z*_*hp*_, and one non-horizontally pleiotropic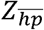. We generate a set of covariates C including those affected by *Z*_*hp*_, and not affected by *Z*_*hp*_, denoted *C*_*hp*_ and 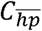, respectively. We vary the number of covariates in *c*_*hp*_ and 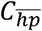, denoted *N*_*chp*_ and 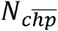, the variance of each covariate in *C*_*hp*_ explained by 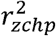, and the correlation between all variables in 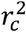. We use a fully factorial design where 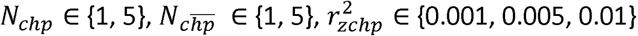 and 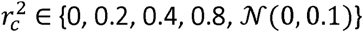. The exposure X and outcome Y are not modelled as, according to our DAG in Figure 2b, the association between the SNPs and covariates are not dependent on X or Y.

All covariates in C are continuous with mean=0 and SD=1. Each SNP is a 3-category ordinal variable (representing SNP dosages) assuming allele frequencies of 0.8 and 0.2 and assuming Heidi Weinberg Equilibrium (such that P_dosage0_=0.64, P_dosage1_=0.32, and P_dosage2_=0.04). A DAG with simulation parameters is shown in Supplementary figure 1b.

#### Estimand or other target

Our target is the test of the null hypothesis of no association between *Z*_*hp*_ and the covariates.

#### Methods

As in simulation A, we compare 4 approaches to test the association of the IV with *C*: the global randomization test, test-Bonf, and test-indep and test-r^2^perm.

#### Performance measures

We evaluate statistical power using rejection percentage [16].

We also estimated these performance measures using 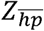 to check that we observe ∼5% type-1 error, i.e., around 5% of the permutations incorrectly identify an association between 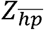 and *C*.

## Applied examples

### Study population

UK Biobank is a prospective cohort of 503 325 men and women in the UK aged between 37–73 years (99.5% were between 40 and 69 years). This cohort includes a large and diverse range of data from blood, urine and saliva samples and health and lifestyle questionnaires. UK Biobank received ethical approval from the UK National Health Service’ s National Research Ethics Service (ref 11/NW/0382). This research was conducted under UK Biobank application number 16729, using phenotypic dataset ID 48196.

Of the 463,005 UK Biobank participants with genetic passing quality control [21], we removed 77,758 minimally related participants, 48,233 non-white British participants, and 39 participants who had since withdrawn from the study. Our sample therefore included 336,975 participants. A data flow diagram is provided in Supplementary figure 2.

### Example 1: Testing for evidence of selection bias

We assess the potential for selection bias in Mendelian randomization studies in UK Biobank that use C-reactive protein (CRP) as the exposure of interest. A previous GWAS meta-analysis (that did not include UK Biobank) identified 58 SNPs robustly associated with CRP [22]. We generate the CRP GRS as a weighted sum of the 58 SNPs, weighted by the effect size of the CRP-increasing allele of each SNP on CRP.

We use two sets of covariates, a restricted set and a more liberal set. The restricted set comprises just age and sex – two factors that cannot be on the causal path between the (constituent SNPs of the) CRP GRS and outcome. The liberal set were chosen as phenotypes that, given our exposure of interest (CRP) we believe are not likely to be either upstream determinants of CRP (or CRP GRS) or downstream effects, such that if an association exists between this set and the CRP GRS we would think it is more likely that this is due to selection bias rather than a causal effect of one or more of the CRP SNPs on (one or more of) these traits. This second set additionally includes socio-economic factors (Townsend deprivation index, age completed full time education), north and east coordinates of home location, and height [23]. Age, sex, home location and education were self-reported at baseline. Sex was validated against genetic sex. Height was measured as baseline, to the nearest cm using a Seca 202 device. The age the participant completed full time education was used as a measure of education level. Townsend deprivation index (a score representing the deprivation of the participant’ s neighbourhood) was calculated immediately prior to participants joining UK Biobank using their self-reported postcode of residence. This gave 7 variables included in the liberal covariate set.

We ran the global randomization test and alternative approaches to test for an association of the CRP GRS with the restricted and liberal covariate sets, respectively. We also repeated these analyses using the rs2794520 cis CRP SNP only, to explore detection of selection bias using the SNP set (in this case just one SNP) that is unlikely to be horizontally pleiotropic.

### Example 2: Testing for evidence of horizontal pleiotropy among CRP SNPs

We used the global randomization test to identify CRP-associated SNPs that may have horizontally pleiotropic effects on coronary heart disease (CHD). We formed our covariate set using a previous study [10] that found little evidence of an association of CIS CRP SNPs with a set of CHD risk factors. These risk factors are therefore unlikely to be on the causal pathway between CRP levels and CHD, such that associations with other CRP-associated SNPs (or a combined CRP GRS) would be most likely due to this SNP being horizontally pleiotropic. These risk factors can therefore be used as the covariate set in the randomization test, to test for evidence of horizontal pleiotropy.

Our covariate set comprised the subset of these phenotypes that were measured in the full UKB sample (e.g., some such as LDL cholesterol were only available in the NMR metabolomics UKB subsample), namely: BMI, systolic blood pressure (SBP), diastolic blood pressure (DBP), total cholesterol, HDL cholesterol, apolipoprotein A1, apolipoprotein B, albumin, lipoprotein A, Leukocyte count, glucose, smoking pack years, weight and waist hip ratio. Details of the covariates are provided in Supplementary section S3. CHD events were ascertained using both self-reported data and linkage to mortality data and hospital inpatient records (see Supplementary section S4 for further details).

We estimated the causal effect of CRP on CHD using two-stage IV probit regression, first using all CRP SNPs and then using only those not identified as horizontally pleiotropic, using a nominal threshold of P<0.05. We use log transformed CRP levels (mg/L) and take the exponent of 1.6 times the estimates, to approximate the association in terms of the change of odds per 1 unit higher log CRP levels [24]. We repeated analyses using a threshold of P<0.001, to assess the sensitivity of results to the stringency of SNP selection.

Analyses were performed in R version 4.0.3, Stata version 15 and Matlab r2015a, and all of our analysis code are available at https://github.com/MRCIEU/MR-randomization-test. Git tag v0.1 corresponds to the version of the analyses presented here.

## RESULTS

### Simulation results

#### Using the randomization test to detect selection bias

Figure 3 shows the results of our selection bias simulations, with an IV strength of r^2^=0.05, including all covariates in the tests of association. Results for IV strength r^2^=0.1 and including half the covariates in the tests of association are shown in Supplementary figure 3. The approach with the highest statistical power depended on the scenario, with the randomization tending to have greater statistical power with low covariate correlations (usually for both the r^2^=0 and r^2^= 𝒩 (0,0.1)).

**Figure 3:**
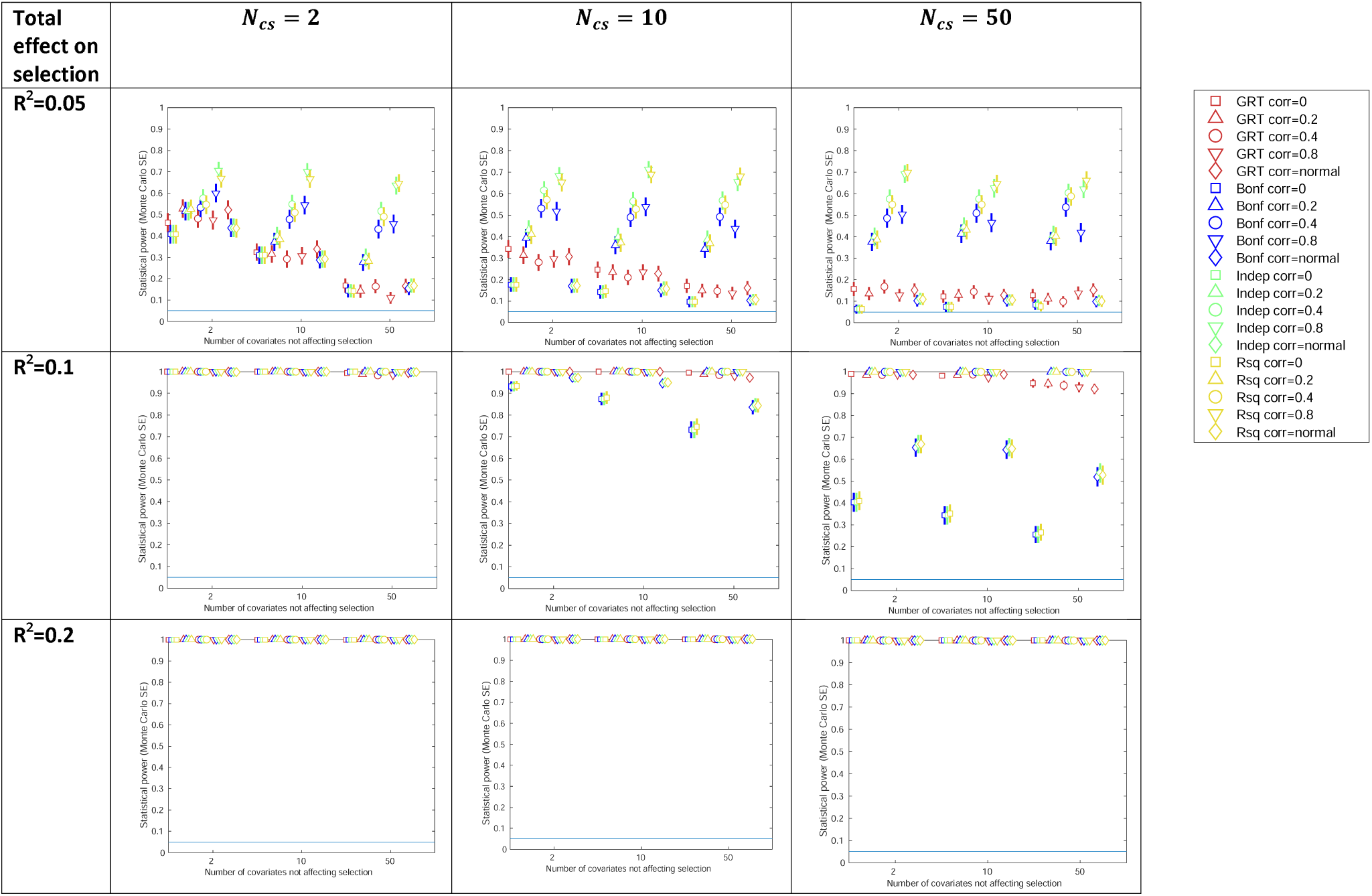
Results of selection bias simulations for instrument strength r^2^=0.05 and all covariates included in test. GRT: global randomization test; SE: standard error. Total effect on selection: the total effect of covariates c_s_ and X on selection S. N c_s_: The number of Covariates affecting selection. Confidence intervals are +/-1.96*MCSE (Monte Carlo standard error).

The statistical power of the global randomization test changed relatively little across covariate correlations, compared with the test-r^2^perm, test-Bonf and test-indep approaches, which were sensitive to this. For example, when there were 10 covariates affecting selection and 2 covariates not affecting selection and a total effect of effect on selection of r^2^=0.05, power ranged between 0.280 (MCSE=0.02) and 0.342 (MCSE=0.02) for the global randomization test (correlation between covariates r^2^=0.4 and 0, respectively), and between 0.172 (MCSE=0.02) and 0.682 (MCSE=0.02) for the test-indep approach (correlation between covariates r^2^=0 and 0.8, respectively).

The statistical power of the global randomization test was well controlled (i.e., with ∼5% type-1 error when no selection bias is present) across all scenarios of our selection bias simulation (see Supplementary figure 4).

#### Using the randomization test to detect horizontal pleiotropy

Figure 4 shows the results of our horizontal pleiotropy simulations. In all except one simulated scenario, statistical power of the global randomization test was either comparable to the power of the alternative tests or had greater power. For example, with an effect of the horizontally pleiotropic SNP of 0.001 on each covariate (Figure 4a), and 5 horizontal pleiotropy covariates *N*_*chp*_ and 5 non 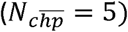, when the covariates were uncorrelated (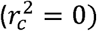 we estimated similar statistical power across all approaches (e.g.0.146 (MCSE=0.02) and 0.118 (MCSE=0.01) for the global randomization and test-indep approaches). In contrast, when the covariates were generated with normally distributed correlation (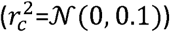), power of the global randomization was larger than the other tests 0.262 (MCSE=0.02) compared with e.g., 0.134 (MCSE=0.02) for the test-indep approach).

**Figure 4:**
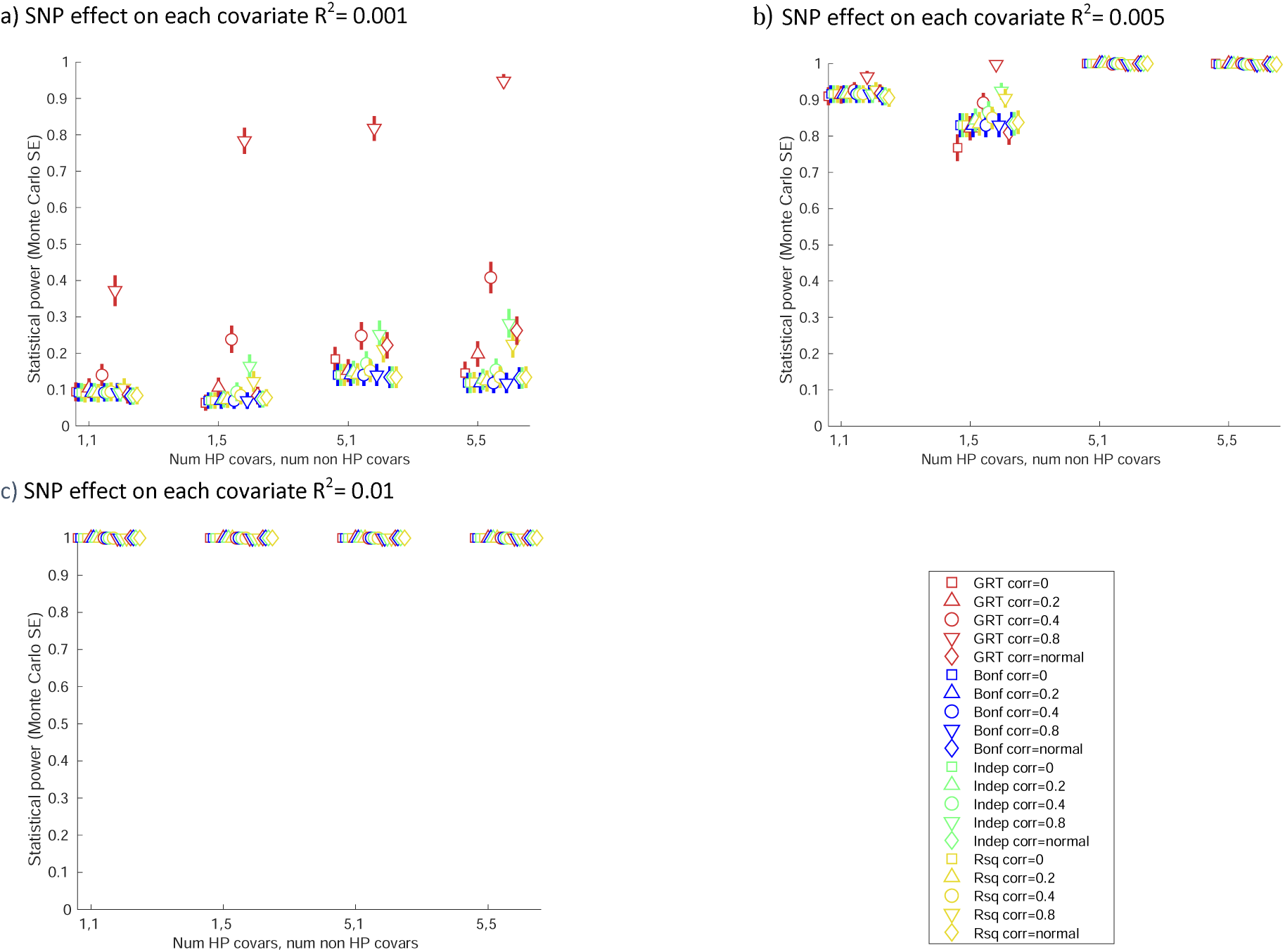
Results of horizontal pleiotropy simulations. GRT: global randomization test; SE: standard error. Confidence intervals are +/- 1.96*MCSE (Monte Carlo standard error).

The statistical power of the global randomization test was well controlled (i.e., with ∼5% type-1 error for non-horizontal pleiotropy SNPs) across all scenarios of our horizontal pleiotropy simulation (see Supplementary figure 6).

## Applied examples

### Detecting selection bias for CRP GRS in UK Biobank

Table 1 shows the results of our CRP selection bias analysis in UK Biobank. We did not detect an association of the CRP GRS with the restricted covariate set (containing only age and sex) using any approach (e.g., p=0.813 using the global randomization test). Using the liberal covariate set we detected an association with the CRP GRS using all approaches (e.g., p<0.002 and p=0.010 for the global randomization test and test-r^2^-perm approaches, respectively). In contrast, we only detected an association with the CRP cis SNP using test-r^2^-perm (p=0.004).

**Table 1:** Results of CRP selection bias applied example.

|  |  | P values |  |  |  |
| --- | --- | --- | --- | --- | --- |
|  |  | CRP GRS |  | CRP cis SNP rs2794520 |  |
|  |  | Restricted covariate set | Liberal covariate set | Restricted covariate set | Liberal covariate set |
| Global Randomization test |  | 0.813 | <0.002 | 0.937 | 0.072 |
| R<sup>2</sup> permutation test (R<sup>2</sup>perm) |  | 0.784 | 0.010 | 0.925 | 0.004 |
| Bonferroni corrected (test-Bonf) * |  | 1.000 (2 tests) | 8.88x10<sup>-9</sup> (7 tests) | 1.000 (2 tests) | 0.110 (7 tests) |
| Independent (test-indep) * |  | 1.000 (2 tests) | 8.88x10<sup>-9</sup> (7 tests) | 1.000 (2 tests) | 0.110 (7 tests) |
| Covariates | Age | 0.840 | 0.840 | 0.923 | 0.923 |
|  | Sex | 0.537 | 0.537 | 0.732 | 0.732 |
|  | Height |  | <0.001 |  | 0.912 |
|  | Home location – northing |  | <0.001 |  | 0.285 |
|  | Home location – easting |  | 0.944 |  | 0.016 |
|  | Years in full time education |  | 0.587 |  | 0.222 |
|  | Townsend deprivation index |  | 0.966 |  | 0.553 |

\* P values calculated as $\min(1, \text{pvalue}_{\min} \times \text{numTests})$ , where $\text{pvalue}_{\min}$ is the lowest p-value of the covariates and numTests is the number of covariates for Bonferroni, and calculated with spectral decomposition for the independent versions. The P values of individual traits are generated with linear regression (IV as independent variable, covariate as dependent variable). The number of tests shown in brackets indicates the effective number of independent phenotypes in the covariate sets, i.e., the effective number of tests to use in the correction for multiple tests.
Baseline education category “None of the above”.

### Detecting horizontally pleiotropic CRP SNPs in UK Biobank

Of the 58 CRP-associated genetic variants, 51 were found to be associated with our defined covariate set using the global randomization test (using a threshold of P<0.05). This compares to 51 identified using the test-Bonf and test-indep approaches, and 46 identified using the r^2^perm approach (see Supplementary table 4). Using a GRS composed of all 58 SNPs, higher genetically predicted CRP levels are associated with a lower risk of CAD (odds ratio [OR]: 0.956 [95% CI: 0.918, 0.996] per 1-unit higher log CRP levels). Using a GRS composed of only the 7 SNPs not identified as horizontally pleiotropic using the global randomization test, and estimates attenuated slightly to the null (e.g., OR: 0.970 [95% CI: 0.900, 1.046] per 1-unit higher log CRP levels). Results of sensitivity analyses using the P<0.001 threshold were similar to the main results (see Supplementary figure 7).

## DISCUSSION

In this study, we have adapted a recently proposed test of association for use in MR studies. The global randomization approach tests the association of a set of covariates with a trait of interest jointly, accounting for the correlation between these covariates, rather than testing the association of the trait of interest with each covariate individually. While the original study [15] proposed the global randomization test as a test for assumption 2 (no association between IV and confounders), assuming that the IV relevance assumption (IV instruments the exposure) and exclusion restriction are valid, in an MR setting, only the relevance assumption is directly testable. The exclusion restriction assumption (in addition to the independence assumption) may not be valid. We therefore focused on demonstrating ways in which the global randomization test can be used to identify violations of the exclusion restriction assumption, using violations due to selection bias and horizontal pleiotropy as examples. We compared the statistical power of this test to that of individual tests of the IV with each covariate with correction for the multiple tests performed using a) Bonferroni, and b) effective number of independent tests (calculated using spectral decomposition). In contrast to these traditional tests of covariate imbalance, the global randomization test uses a permutation-based approach to test the association of a set of covariates jointly with an IV. We also explored an alternative permutation-based approach, using the highest correlation between the covariates and IV as the test statistic.

We used simulations to investigate the statistical power of these approaches to detect selection bias and horizontal pleiotropy under different scenarios. Our selection bias simulations suggested that the global randomization test tends to have better power compared to the alternative approaches, when covariate correlations are lower. While we do not have enough information to suggest a cutoff in general, our simulations suggest the global randomization test (with MD test statistic) could be used when covariate correlations are below 0.1. In our horizontal pleiotropy simulations, the global randomization test had either similar or better power compared to the alternative approaches across all except one of the simulated scenarios. We would therefore recommend use of the global randomization test to test for horizontal pleiotropy, but we note we have not assessed every different scenario in our simulations.

We demonstrated how the global randomization test can be used in practice with two applied examples. The first sought to investigate whether MR analyses of CRP may be biased due to non-random selection in UK Biobank. We used a restricted and liberal covariate set, the former containing just age and sex, while the latter contained 5 additional variables that are unlikely to be downstream determinants of CRP genetic variants. We found evidence using all test approaches (including the global randomization test) suggested that non-random selection may bias MR estimates of CRP. The second applied example sought to identify horizontally pleiotropic CRP-associated SNPs, when estimating the effect of CRP on coronary artery disease. The global randomization test identified 51 of the SNPs as potentially horizontally pleiotropic and estimates attenuated to the null after excluding these SNPs, although confidence intervals were wide.

Our results suggest that the global randomization test may be a useful falsification test in MR. However, one potential challenge in applying this test in this setting is the choice of covariate set. In an MR setting, most phenotypes can theoretically be downstream of a genetic variant (age and sex being two key exceptions), such that it may be difficult to identify candidate covariates to include in the global randomization test, where we believe with confidence that these covariates are not on the causal pathway between the IV and exposure, or between the exposure and outcome. In short, to use the test the researcher needs to assume that, if the IV associates with the candidate covariate set, this is more likely to be due to an invalid IV assumption rather than because these covariates are on the vertically pleiotropic pathway. Box 1 summarises the approach researchers can take to use the global randomization test to explore violations of the exclusion restriction assumption due to selection bias or horizontal pleiotropy. Where covariates may associate with the IV due to both selection bias and horizontal pleiotropy, it may be useful to first conduct a combined test for these using a combined covariate set. The value of this compared to testing for selection bias and horizontal pleiotropy could be investigated in future work.

**Box 1:** Overview of process to use the global randomization test

**Box 1: Overview of process to use the global randomization test**
|  | Testing for evidence of selection bias | Testing for evidence of horizontal pleiotropy |
| --- | --- | --- |
| 1. Choose covariates for inclusion in covariate set | Covariates should be selected that are thought to be unaffected by the genetic instrument (and hence also not affected by the exposure phenotype), but that are potential determinants of selection. | Covariates should be selected that are not on the causal path between genetic instrument and exposure phenotype and are also not downstream effects of the exposure. |
| 2. Choose test of association between genetic instrument and covariate set | If mean $r^2$ between covariates $< 0.1$:<br>- Use global randomization test.<br>Otherwise:<br>- Use individual tests of each covariate with correction for the effective number of tests performed (test-indep). | Use global randomization test. |
| 3. Choose follow-up analysis | Option 1: If confident that covariates are not consequences of the genetic instrument, conduct MR analysis conditioning on them, as if condition on all causes of selection you remove the selection bias.<br>Option 2: Test associations of covariate set with outcome. If covariates are not related to the outcome, then bias due to selection may be negligible.<br>Option 3: Conduct simulations to explore the magnitude of the selection bias that would be needed to give the observed (or null) effect, assuming no (or positive or negative effect) true effect. | Include SNPs not identified as associated with the covariate set in genetic IV, and re-estimate causal effect.<br>Compare estimate and confidence interval with estimate using all SNPs. |
| 4. Triangulation | Test association of covariate set in a second cohort with weaker or different selection mechanism. For example, if this second cohort has less selection and the association between the IV and covariate set weakens then this strengthens the evidence for selection bias in the original cohort. | Test association of covariate set in a second cohort with weaker or different selection mechanism. For example, if this second cohort has less selection and the association between the IV and covariate set weakens then this suggests that this association is at least in part driven by selection bias rather than (solely by) horizontal pleiotropy. |

In addition to the approaches using a covariate set that we focus on in this study, there are other falsification tests for exclusion restriction assumption. Heterogeneity of effects across SNPs can be tested for, as if exclusion restriction assumption holds (in addition to the relevance and independence assumptions) the estimated effect should be consistent across IVs. Tests for this include the Hansen test, or by splitting the SNPs into distinct sets and comparing the estimated effects of the exposure using these sets [25]. These approaches test for evidence of horizontal pleiotropy without needing to specify the set of covariates for the particular SNP, through which this pleiotropy may act. However, these may have low statistical power because they are comparing estimates of effect on the outcome across instruments. Steiger filtering is an approach that removes SNPs from a GRS where they explain more variation in the outcome than the exposure, such that their effect on the exposure may be more likely to be via the outcome rather than vice-versa [26]. As we have shown, the global randomization test can be used in a complementary way, to identify SNPs that may be invalid because they correlate with factors that we do not believe could be on the causal pathway (i.e., between IV and exposure or between exposure and outcome), and hence can be removed from a GRS. This ‘ randomization filtering’ can be used as a sensitivity analysis in MR studies.

Our study has a number of strengths and limitations. Strengths include the fact that we explored the value of the global randomization test using both simulations and applied examples. We tried to simulate realistic scenarios by basing aspects such as the recruitment rate on real data (in this case UK Biobank). However, we were only being able to simulate a limited number of scenarios, such that we cannot infer how the statistical power of the global randomization test compares to the alternative approaches beyond these. We generalized the Mahalanobis distance used in [15] to allow continuous and ordinal instruments as well as binary. Our approach assumes a linear relationship between the IV and each of the covariates, such that non-linear associations may have been missed. We used the global randomization test to test the association of a single genetic variant or independent SNPs combined into a GRS, with a covariate set. It may be possible to extend this to incorporate multiple correlated SNPs, for example, those used in cis-MR studies [27]. We did not adjust for any covariates (e.g., genetic principal components) in our examples, but in future work using this approach the genetic instrument and covariates could be regressed on potential confounders and then the residuals from those regressions can be used in the test process.

In summary, the global randomization test can be used as a first step for identifying potential violations of Mendelian randomization assumptions. Where an association is identified that suggests horizontal pleiotropy, a researcher can then investigate this further, for example, by using instruments for variables in the covariate set to test for an effect of these on the outcome of interest [28]. The choice of covariate set used with the global randomization test needs careful consideration in the context of the specific exposure and outcome being examined. While we have focused on falsification tests for the exclusion restriction assumption, this approach may also be useful as a falsification test for the independence assumption, for example, testing for confounding via dynastic effects.

## Data Availability

UK Biobank data can be accessed by bona fide researchers through the UK Biobank data access portal.

## STATEMENTS AND DECLARATIONS

### Author contributions

LACM led the design of the study, performed all analyses, wrote the first version of the manuscript, critically reviewed and revised the manuscript, and approved the final version of the manuscript as submitted. GDS contributed to the design of the study, critically reviewed and revised the manuscript, and approved the final version of the manuscript as submitted. KT conceptualized the study, contributed to the design of the study, critically reviewed and revised the manuscript, and approved the final version of the manuscript as submitted.

### Ethics approval

UK Biobank received ethical approval from the UK National Health Service’ s National Research Ethics Service (ref 11/NW/0382). This research was approved under UK Biobank application number 16729.

