## Supplementary information for "The global randomization test: A Mendelian randomization falsification test for the exclusion restriction assumption"

Millard et al.

**SUPPLEMENTARY INFORMATION**

### SUPPLEMENTARY TEXT

#### Supplementary section S1: Details of selection bias simulation data generating mechanism

*Controlling the variance of X explained by* $C_{s}$ *and* $C_{\bar{s}}$ *and Z*

To control the variance of X explained by $C_{s}$ and $C_{\bar{s}}$ and Z, we first generate two intermediate variables that combine the covariates in $C_{s}$ and $C_{\bar{s}}$, respectively, into two intermediate variables:

$$I_{CS}=\sum_{i=1}^{N_{cs}} C_{s}\left( i \right)$$

$$I_{C\bar{S}}=\sum_{i=1}^{N_{cs}} C_{\bar{s}}(i)$$

Then, X is generated as:

$$X=\beta_{z}Z+\beta_{ICS}I_{CS}+ \beta_{IC\bar{S}}I_{C\bar{S}}+ \beta_{\in}\in$$

Where $\in$ is normally distributed random error.

The total variance of X can be defined in terms of the variances and covariances of Z, $I_{CS}$ and $I_{C\bar{S}}$:

$$var\left( total \right)={\beta_{z}}^{2}var\left( Z \right)+{\beta_{ICS}}^{2}var\left( I_{CS} \right)+ {\beta_{IC\bar{S}}}^{2}var\left( I_{C\bar{S}} \right)+2\beta_{z}\beta_{ICS}\mathrm{cov}\left( Z,I_{CS} \right)+ 2\beta_{Z}\beta_{IC\bar{S}}\mathrm{cov}\left( Z,I_{C\bar{S}} \right)+2\beta_{ICS}\beta_{IC\bar{S}}\mathrm{cov}\left( I_{CS},I_{C\bar{S}} \right)$$

And given that Z and C are independent $\mathrm{cov}\left( Z,I_{CS} \right)=0$ and $\mathrm{cov}\left( Z,I_{C\bar{S}} \right)=0$ such that:

$$var\left( total \right)={\beta_{z}}^{2}var\left( Z \right)+{\beta_{ICS}}^{2}var\left( I_{CS} \right)+ {\beta_{IC\bar{S}}}^{2}var\left( I_{C\bar{S}} \right)+2\beta_{ICS}\beta_{IC\bar{S}}\mathrm{cov}\left( I_{CS},I_{C\bar{S}} \right)$$

We fix the variance of X explained by Z (i.e. the total effect of Z on X), to $r_{zx}^{2}$ $\in$ {0.05, 0.1}:

$$\beta_{z}= \sqrt{r_{zx}^{2}}$$

Then set ${\beta=\beta}_{ICS}=\beta_{IC\bar{S}}$ such that the total variance explained by $I_{CS}$ and $I_{C\bar{S}}$ combined is 10%:

$$0.1=\beta^{2}var\left( I_{CS} \right)+ \beta^{2}var\left( I_{C\bar{S}} \right)+2\beta^{2}\mathrm{cov}\left( I_{CS},I_{C\bar{S}} \right)$$

$$\beta=\sqrt{\frac{0.1}{var(I_{CS})+var(I_{C\bar{S}})+2(\mathrm{cov}\left( I_{CS},I_{C\bar{S}} \right))}}$$

The variance of X explained by the error term is 0.8, such that $\beta_{\in}=\surd0.8$.

*Controlling the variance of S explained by* $C_{s}$ *and X*

For the selection binary variable S, we also fix the total effects. We assumed a total effect of X and $C_{s}$ on S of r^2^ = {0.05, 0.1, 0.2}. We assume 5.5% are selected into our sample (the proportion of those invited who agreed to participate [8]). To generate S we use intermediate covariate $I_{CS}$ to first generate a continuous selection variable, $S_{cont}$, with mean zero and sd=1:

$$S_{cont}=\beta x+\beta I_{CS}$$

Where

$$\beta=\sqrt{\frac{1}{var(X)+var(I_{CS})+2(\mathrm{cor}\left( X,I_{CS} \right))}}$$

i.e. we are assuming X and $I_{CS}$ explain 100% of the variance of $S_{cont}$. We then generate S using $S_{cont}$, changing the odds ratio and intercept in this model accordingly to give r^2^ = {0.05, 0.1, 0.2}, where the probability of being selected, $P_{S}$, is given by:

$$P_{S}=\frac{\exp\left( \log\left( OR \right)\times S_{cont}+intercept \right)}{1+exp(\log\left( OR \right)\times S_{cont}+intercept)}$$

#### Supplementary section S2: Simulation performance measure

We will evaluate statistical power using rejection percentage [7]:

$$RP= \frac{1}{n_{sim}}\sum_{i=1}^{n_{sim}} 1(p_{i}\leq0.05)$$

Where *p_i_* is the p-value of simulation iteration *I,* and *n_sim_* is the number of simulation repetitions, which we set to 500.

Monte Carlo standard error (SE) is estimated as:

$$\sqrt{\frac{RP \times(1-RP)}{n_{sim}}}$$

#### Supplementary section S3: Details of covariates used in horizontal pleiotropy applied example

Weight and height were both measured at baseline assessment. Weight was measured (to the nearest 100 g) in light clothing and unshod using a Tanita BC418MA body composition analyser and height to the nearest cm using a Seca 202 device. We used the mean SBP and DBP, respectively, from two resting automated measures, measured using an Omron HEM-7105IT digital blood pressure monitor. Blood samples were collected at baseline and total cholesterol, HDL cholesterol, apolipoprotein A1, apolipoprotein B, albumin, lipoprotein A, leukocyte count and glucose were measured by immunoturbidimetric analysis on a Beckman Coulter AU5800. Waist and hip circumference were measured at baseline and used to derive waist hip ratio. Age participants started smoking, stopped smoking and the number of cigarettes smoked per day were used to derive a measure of smoking pack years, with those who have never smoked assigned the value zero.

#### Supplementary section S4: Derivation of coronary heart disease phenotype

We derived a binary variable denoting coronary heart disease, where participants with a date of first occurrence for any of the following diseases were assigned as having coronary artery disease:

- Angina pectoris (UK Biobank field ID 131296)
- Acute myocardial infarction (UK Biobank field ID 131298)
- Subsequent myocardial infarction (UK Biobank field ID 131300)
- Certain current complications following acute myocardial infarction (UK Biobank field ID 131302)
- Other acute ischaemic heart diseases (UK Biobank field ID 131304)
- Chronic ischaemic heart disease (UK Biobank field ID 131306)

,

### SUPPLEMENTARY FIGURES

#### Supplementary figure 1: DAG for simulations with parameters used

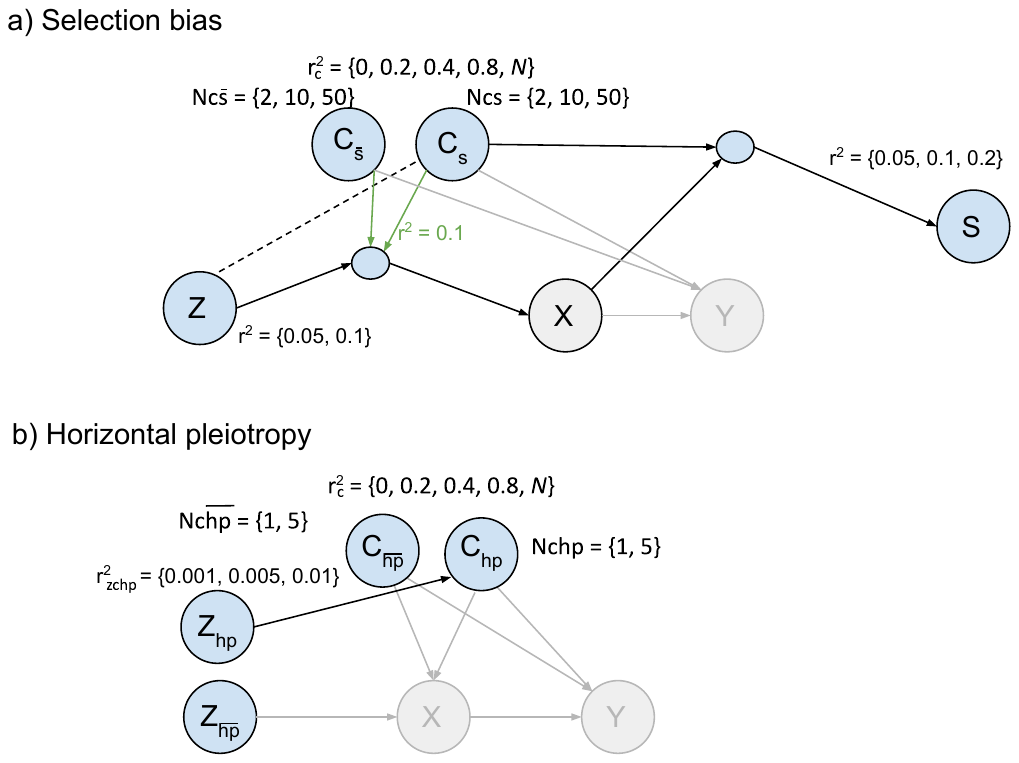

DAGs show the simulation parameters used in a fully factorial design. The grey edges and nodes are those that do not impact the simulation, so the relationships depicted by these edges do not need to be defined and the variables depicted by these nodes do not need to be generated.

The green r^2^=0.1 is the total effect of the covariates on X (with r^2^=0.05 from $C_{s}$ and $C_{\bar{s}}$, respectively).

#### Supplementary figure 2: Sample for applied UK Biobank examples

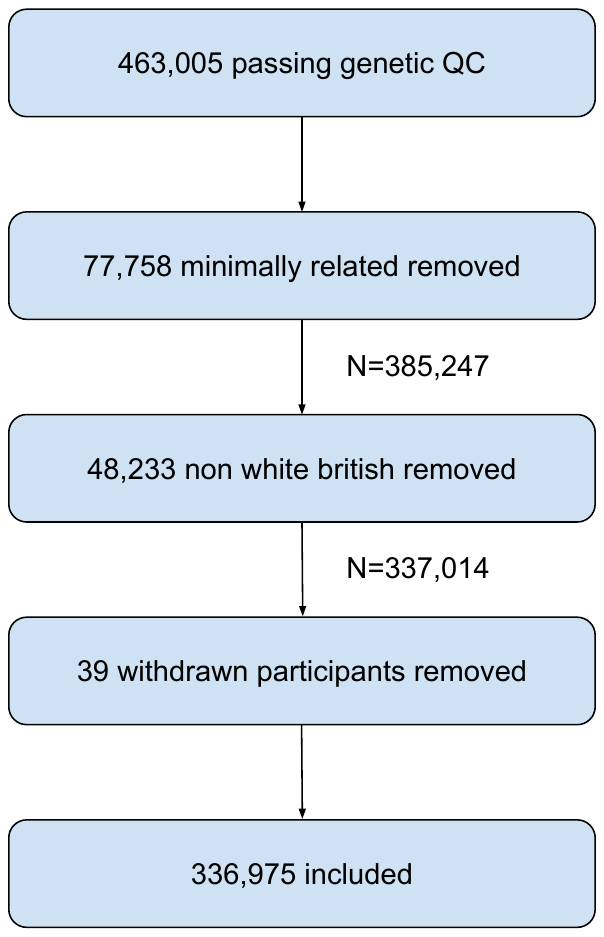

#### Supplementary figure 3: Results of selection bias simulations

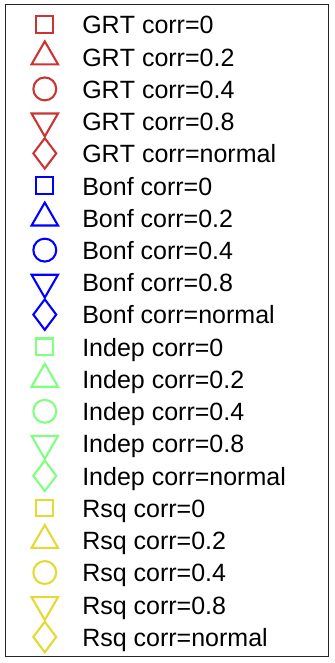
a) Instrument strength rsq=0.05, half covariates included in test

| **Total effect on selection** | $\boldsymbol{N}_{\boldsymbol{cs}}\boldsymbol{=2}$ | $\boldsymbol{N}_{\boldsymbol{cs}}\boldsymbol{=10}$ | $\boldsymbol{N}_{\boldsymbol{cs}}\boldsymbol{=50}$ |
| --- | --- | --- | --- |
| **R^2^=0.05** | 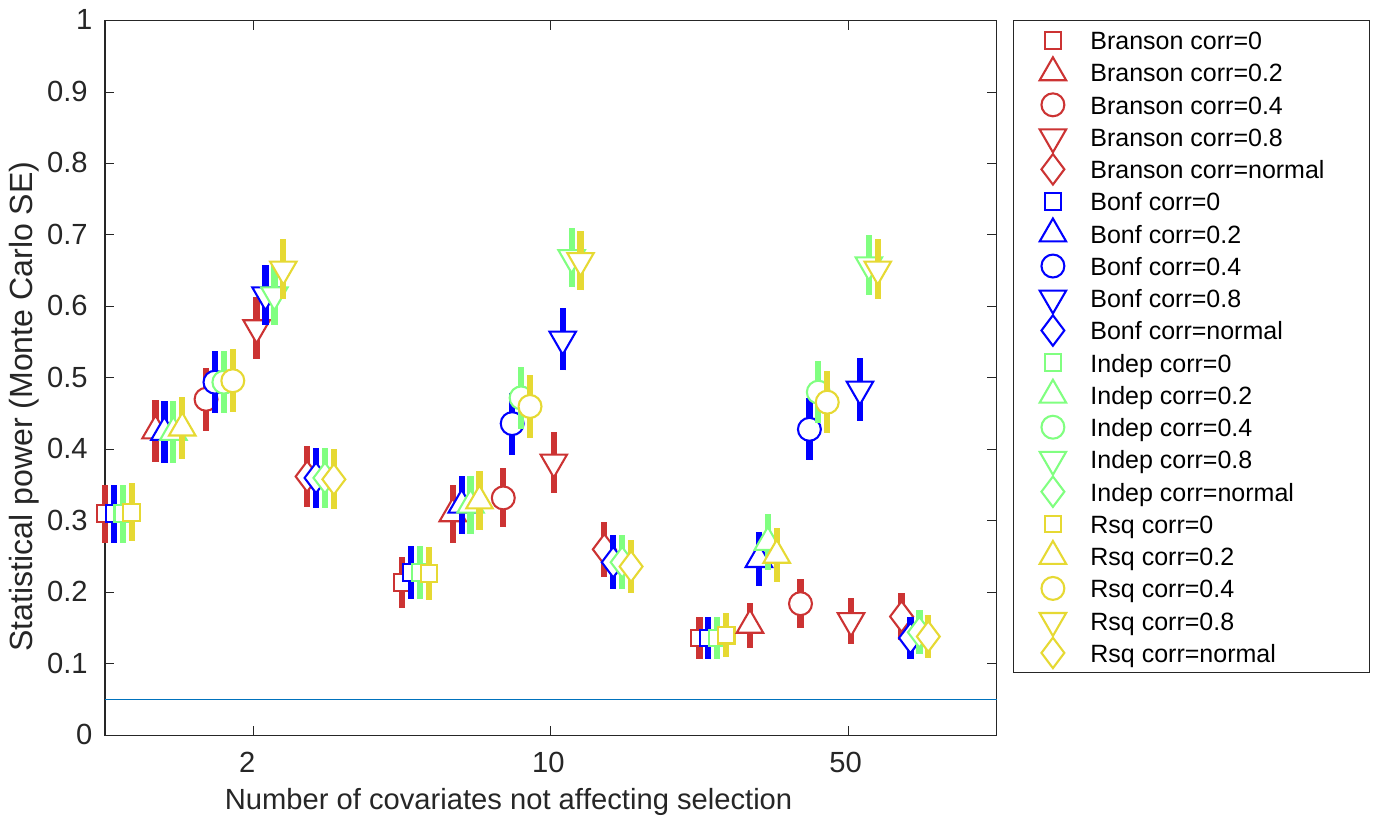 | 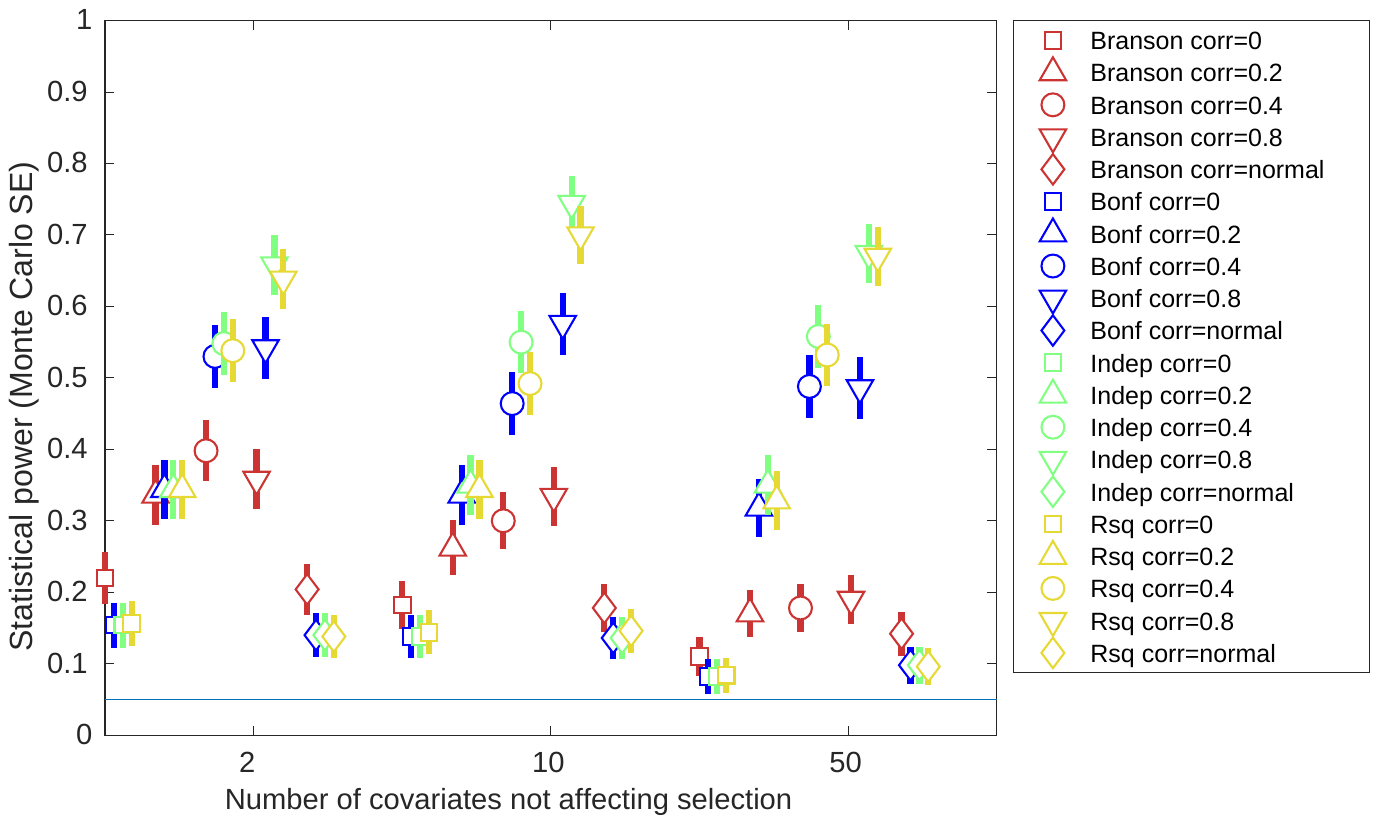 | 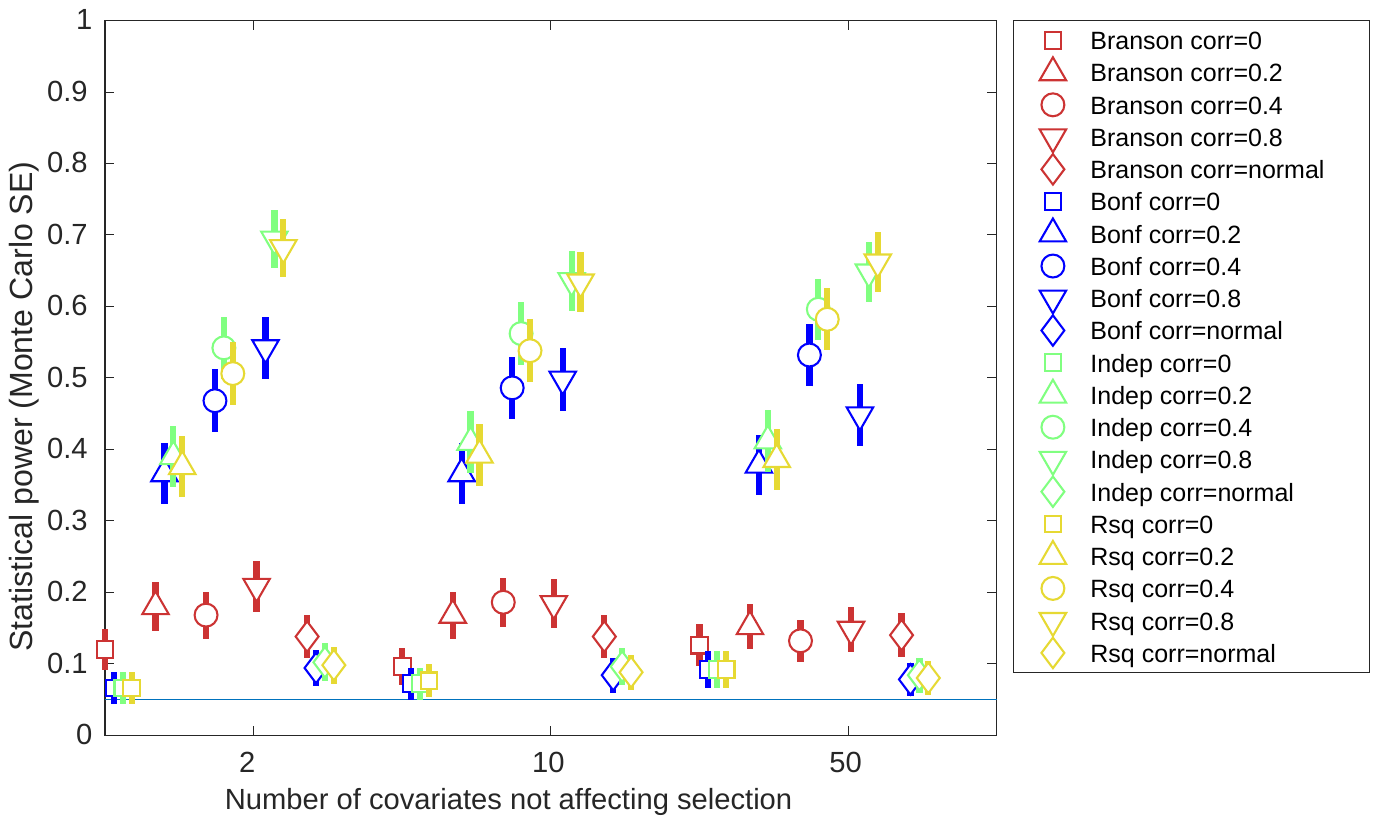 |
| **R^2^=0.1** | 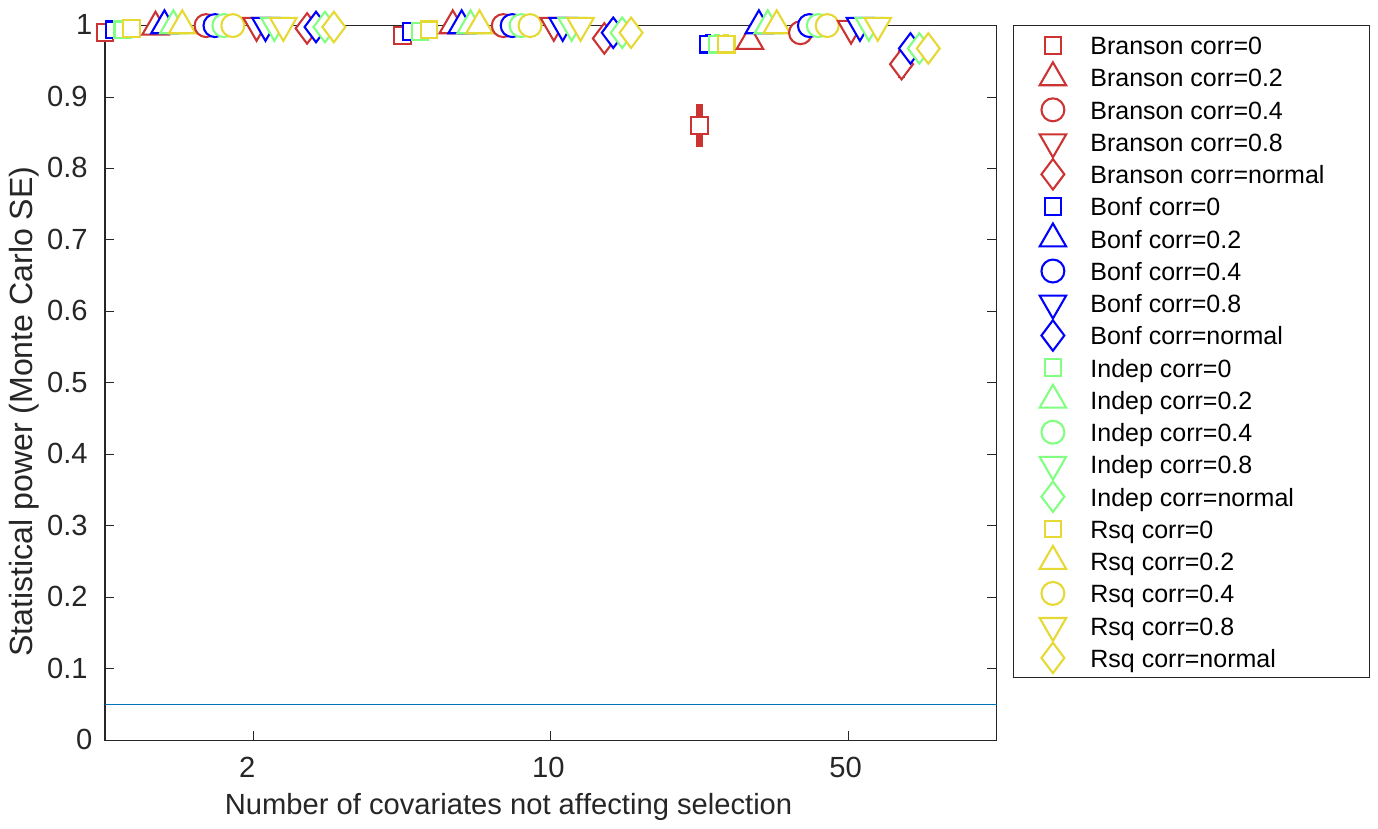 | 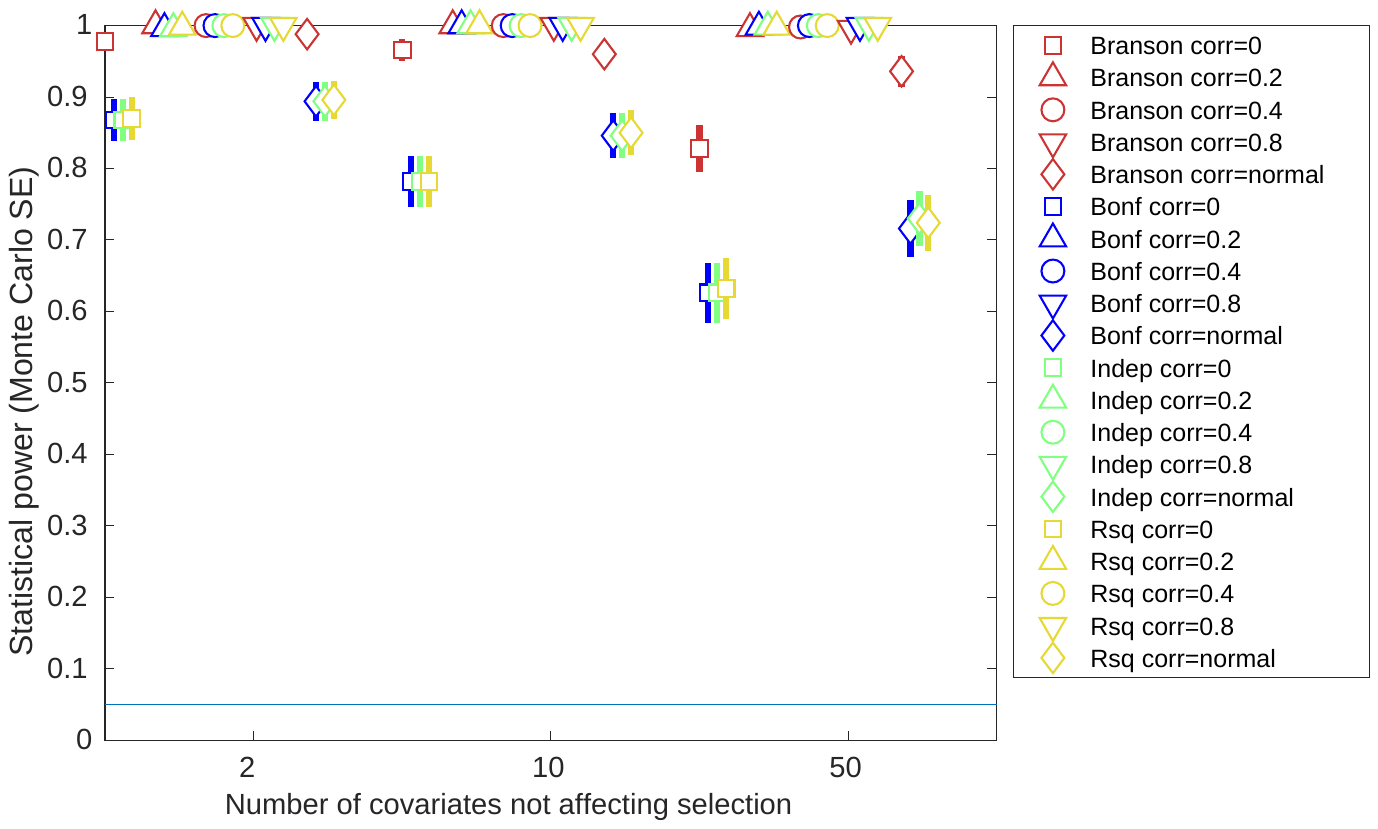 | 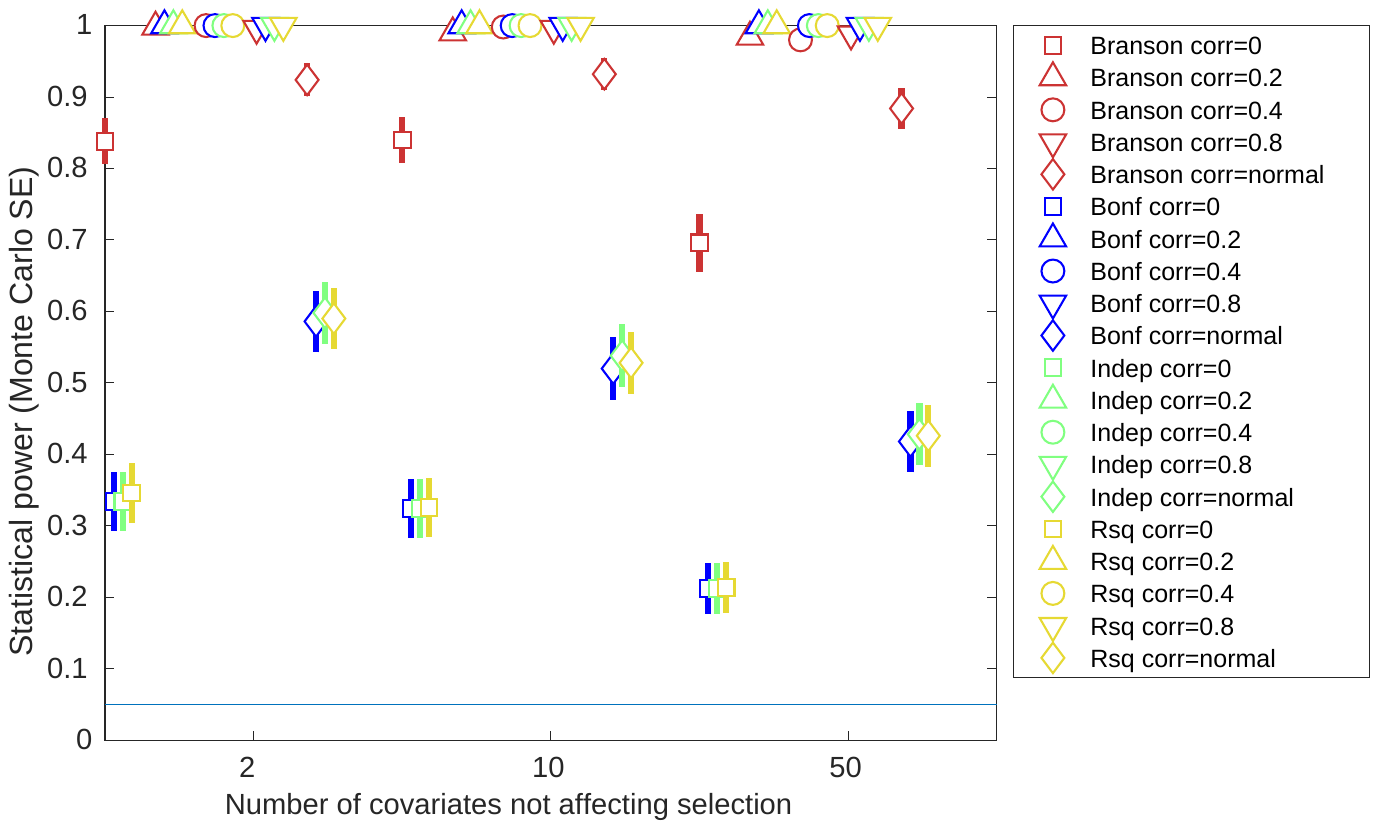 |
| **R^2^=0.2** | 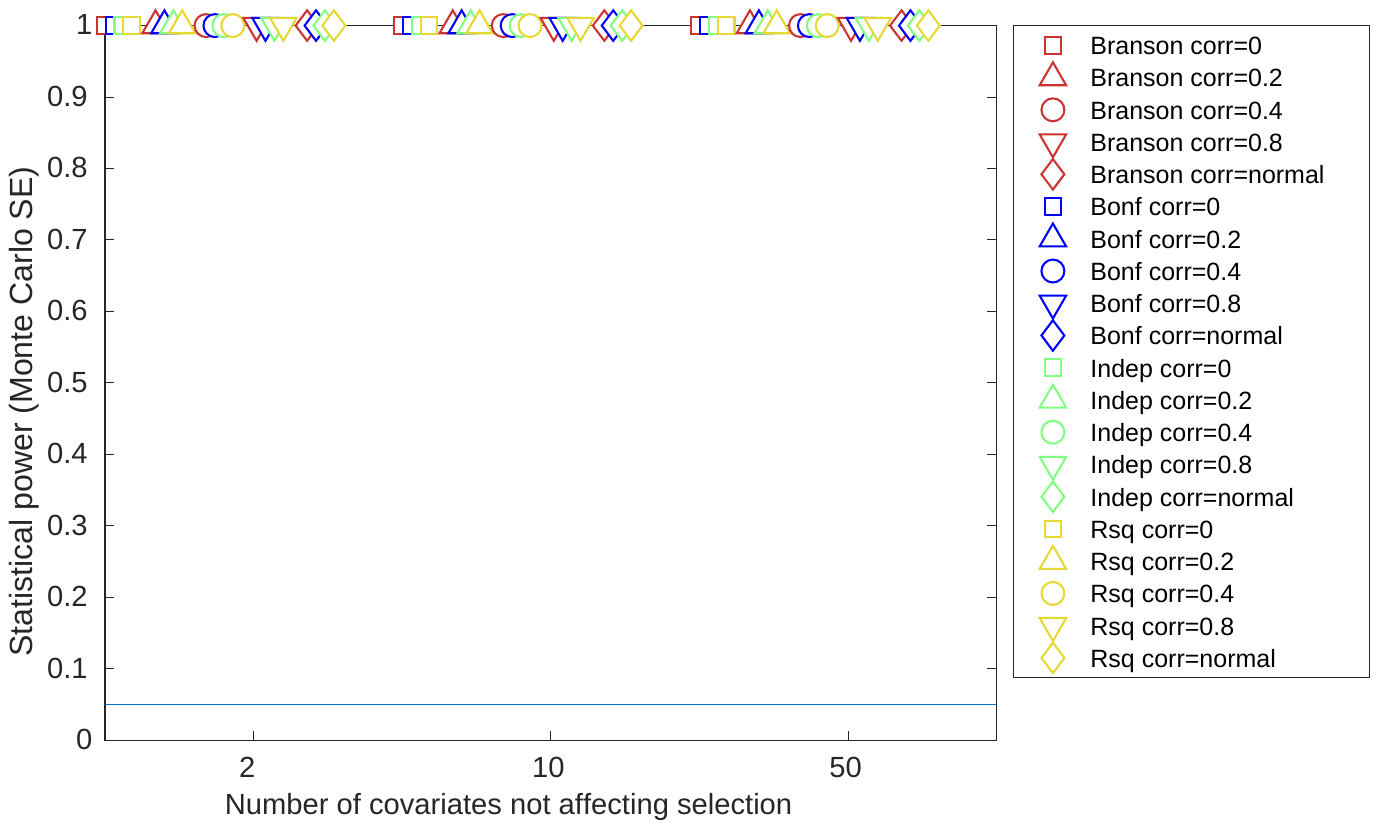 | 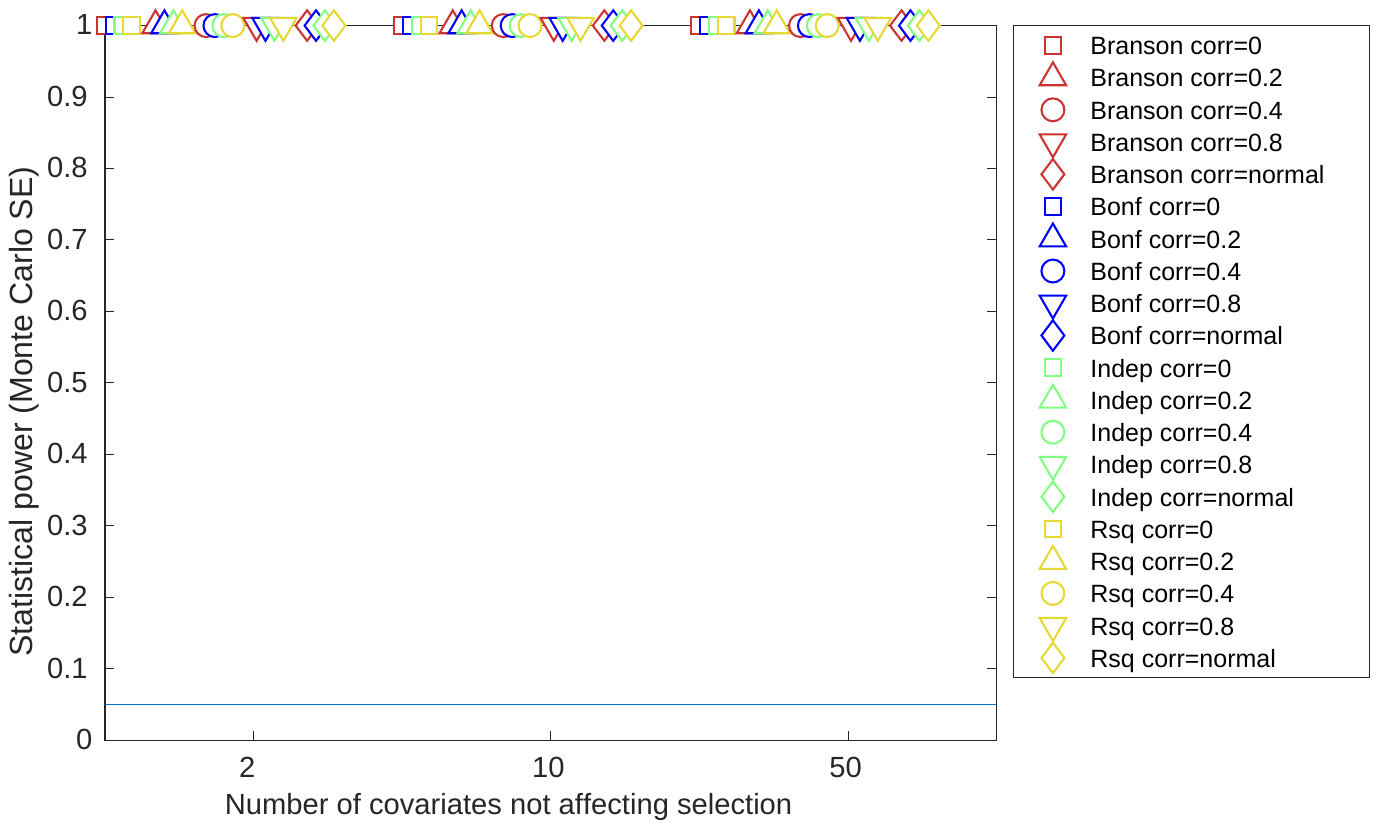 | 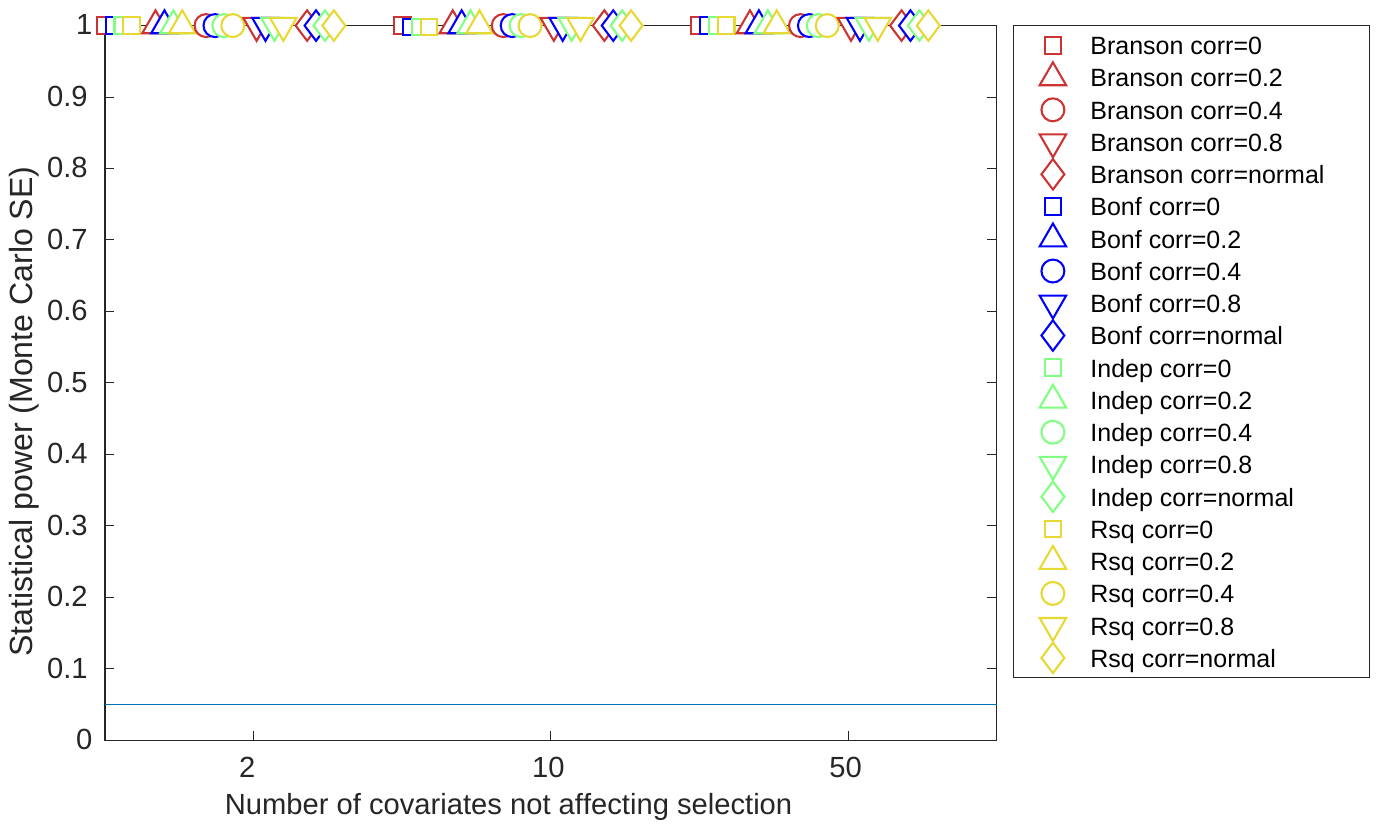 |

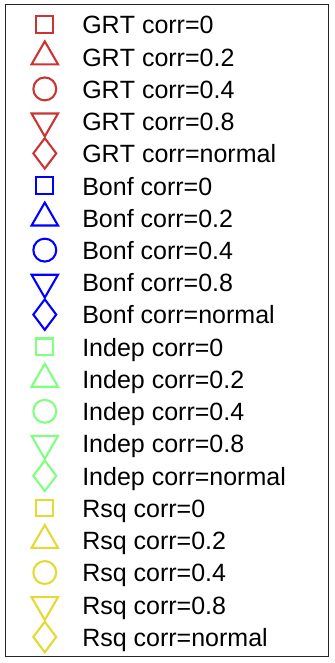
b) Instrument strength rsq=0.1, all covariates included in test

| **Total effect on selection** | $\boldsymbol{N}_{\boldsymbol{cs}}\boldsymbol{=2}$ | $\boldsymbol{N}_{\boldsymbol{cs}}\boldsymbol{=10}$ | $\boldsymbol{N}_{\boldsymbol{cs}}\boldsymbol{=50}$ |
| --- | --- | --- | --- |
| **R^2^=0.05** | 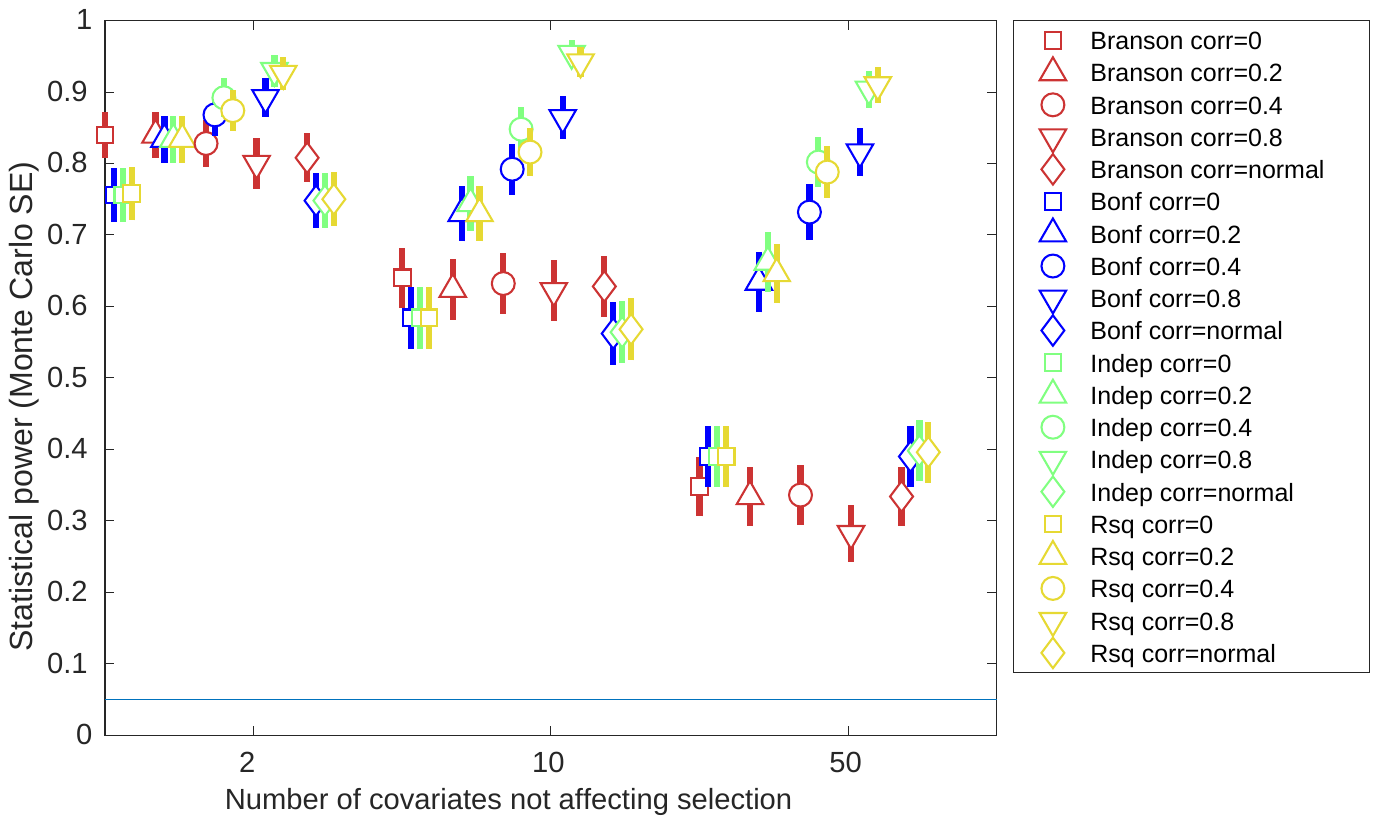 | 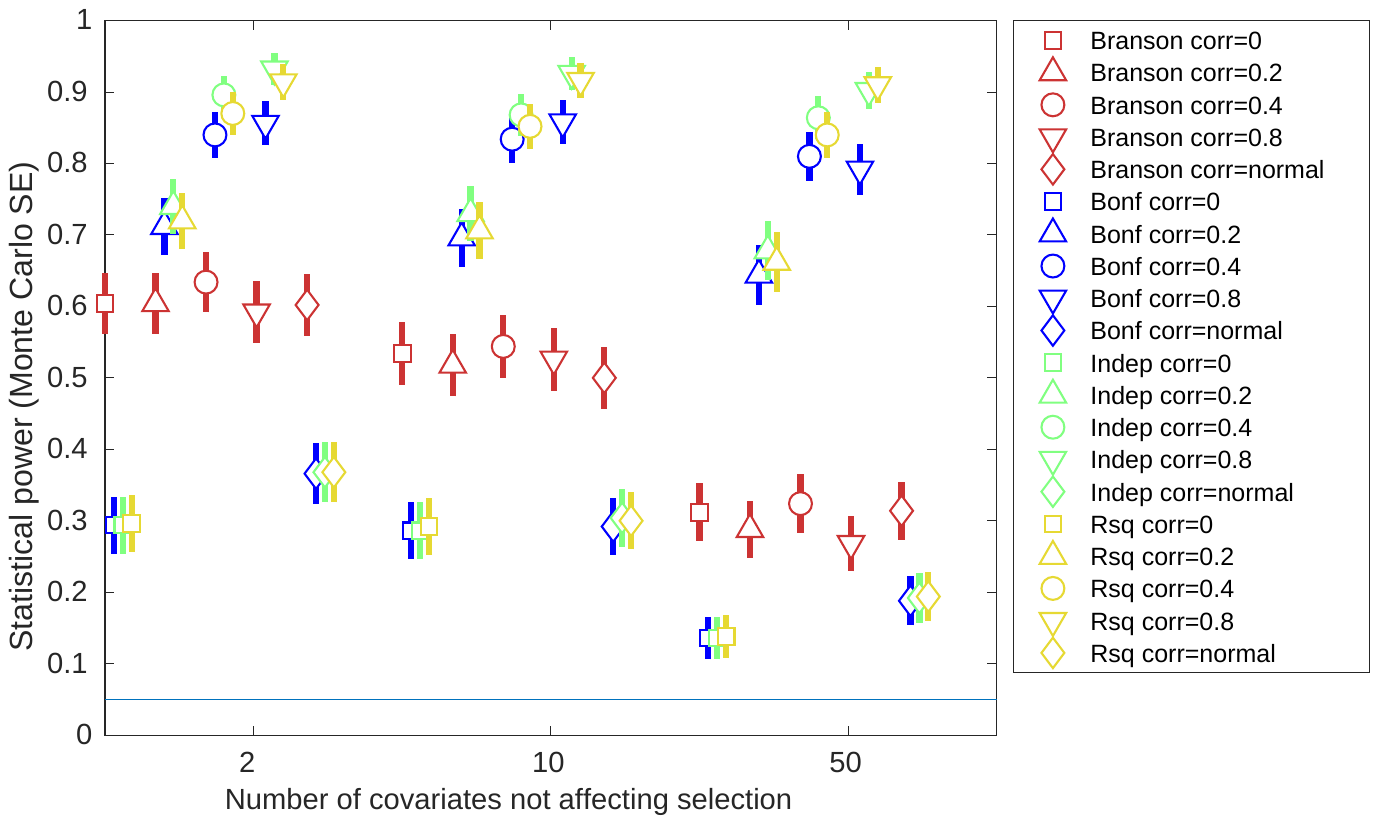 | 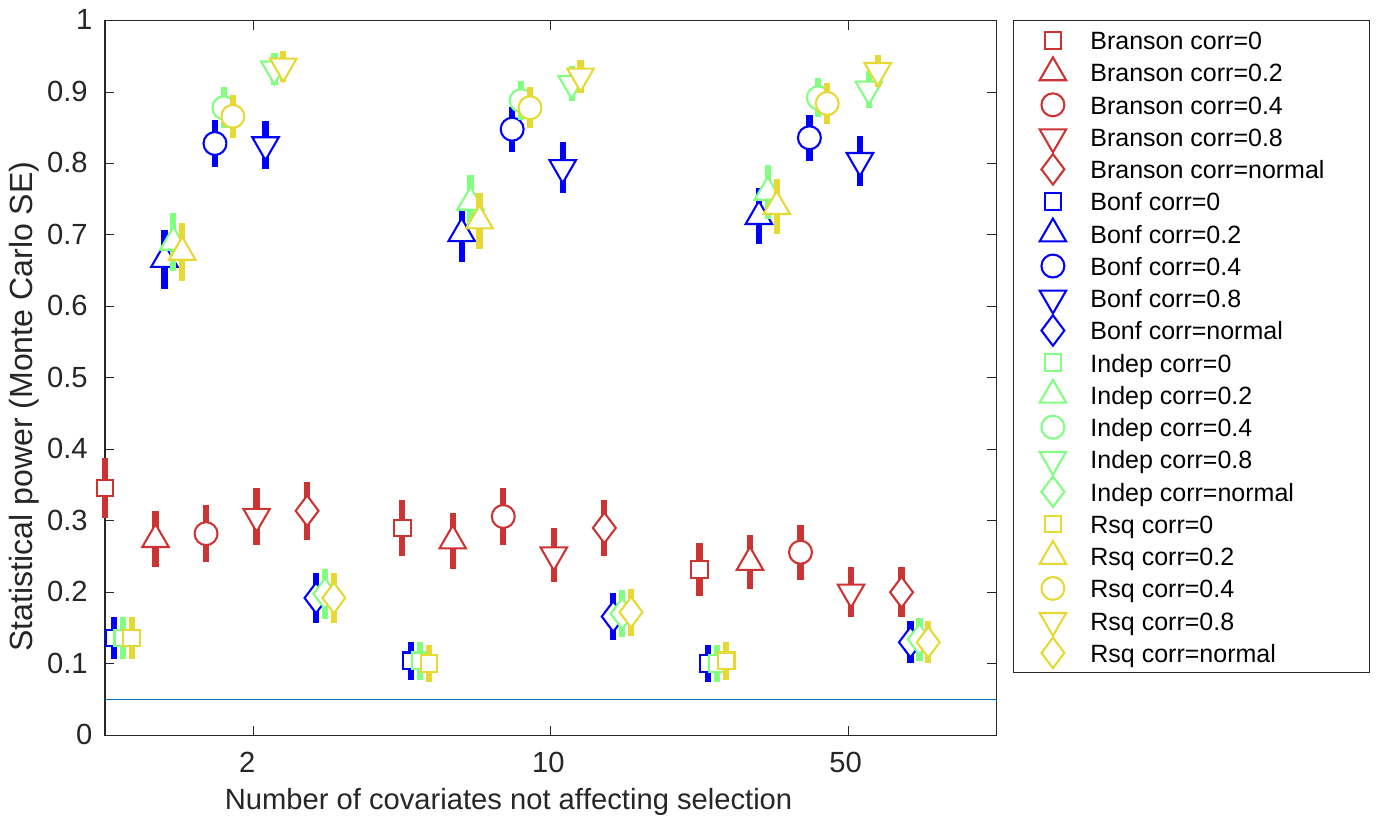 |
| **R^2^=0.1** | 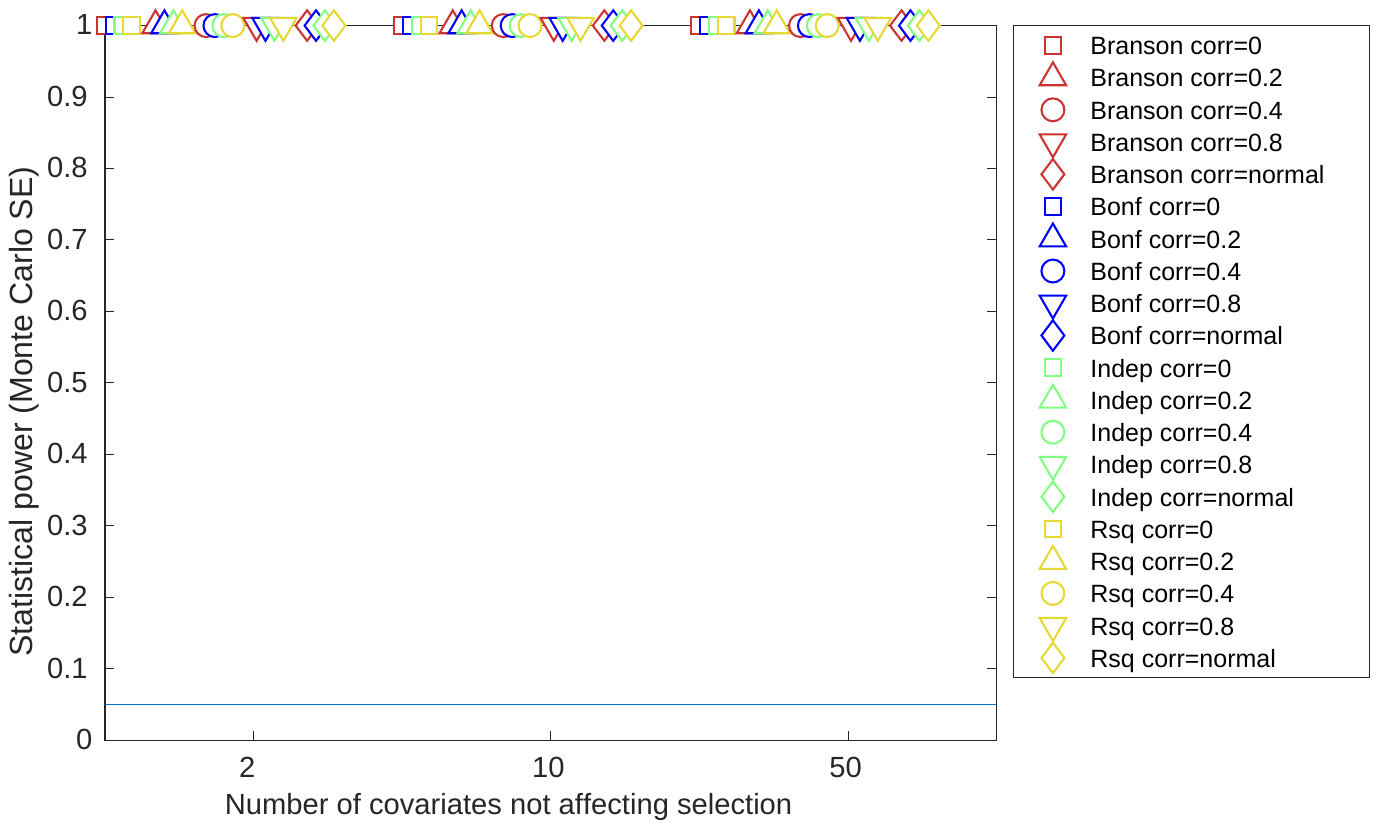 | 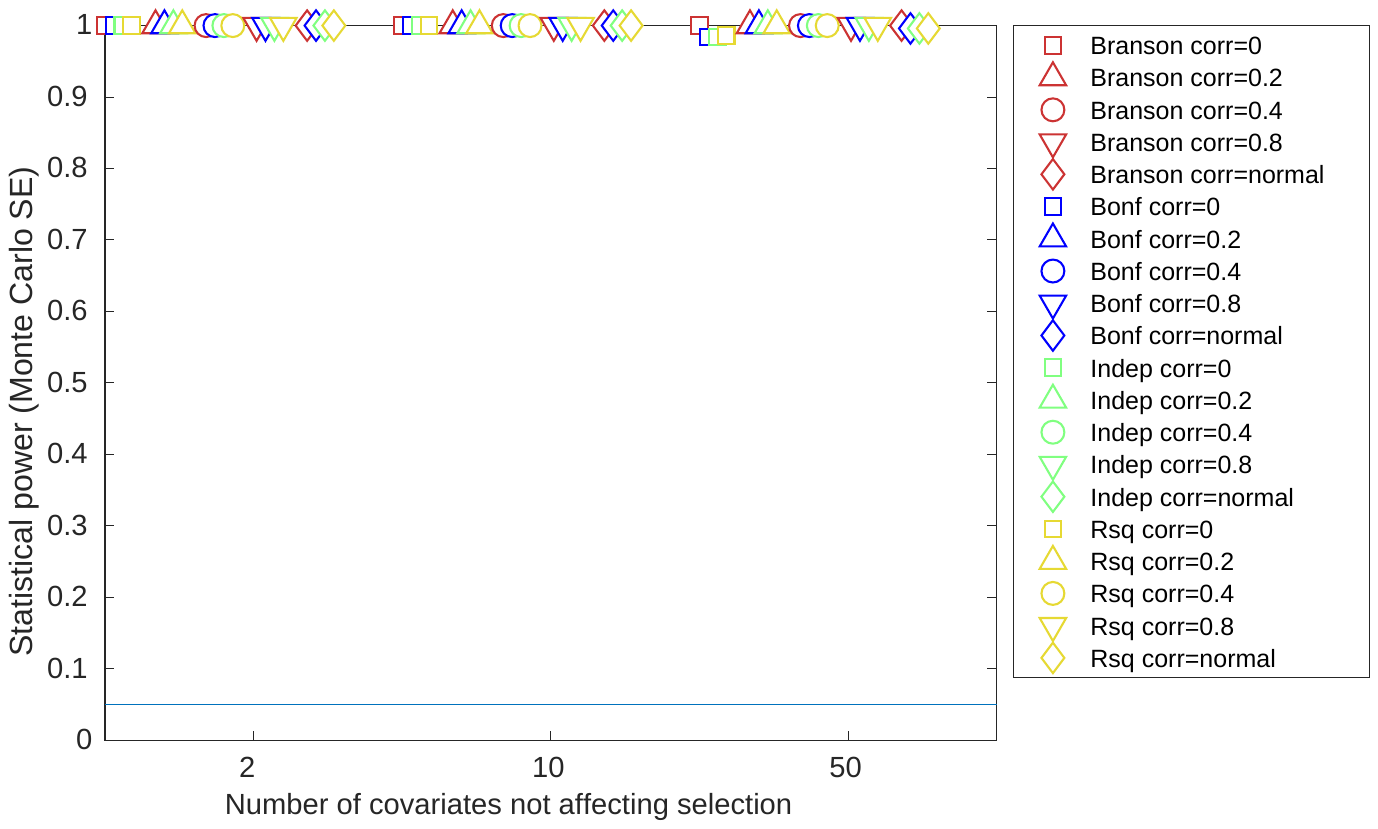 | 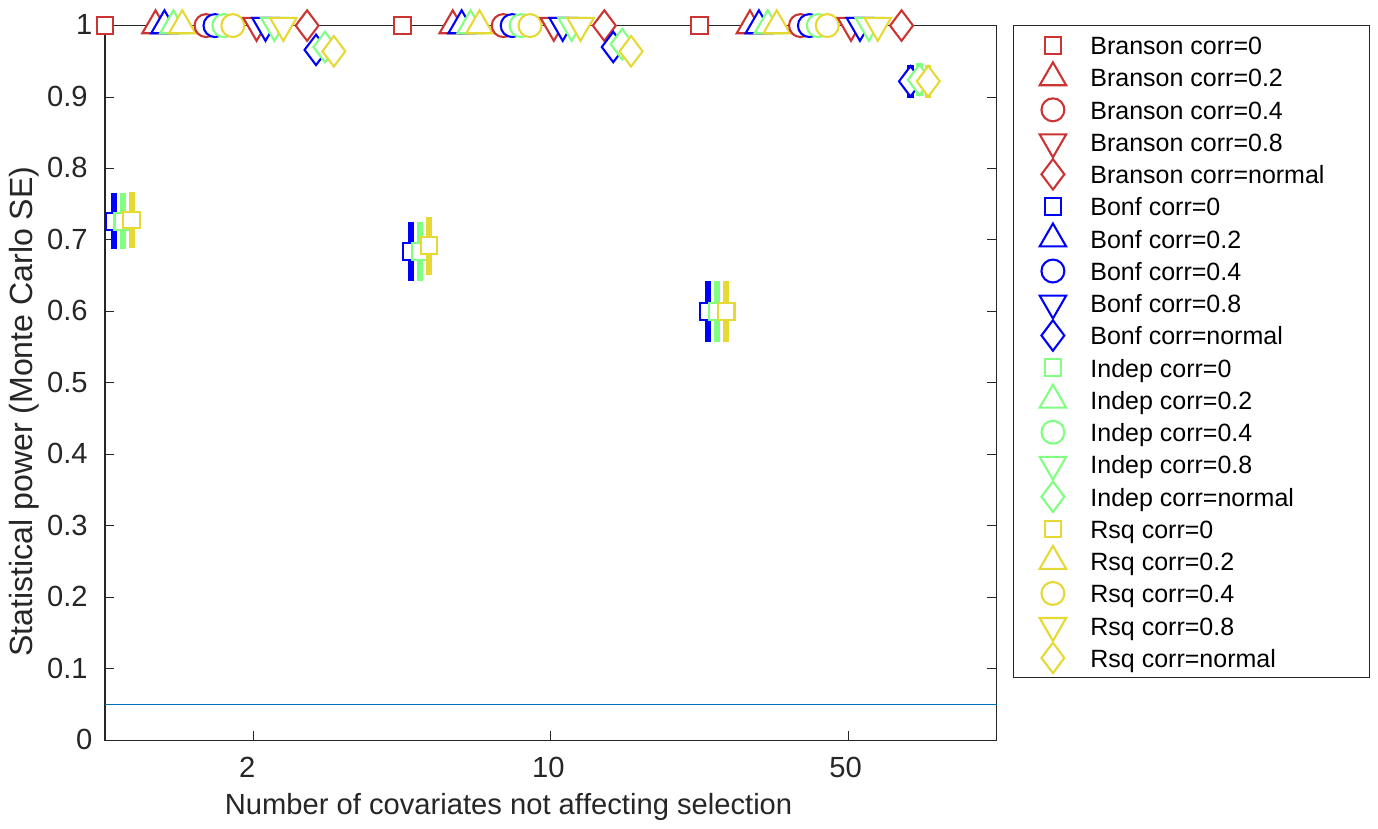 |
| **R^2^=0.2** | 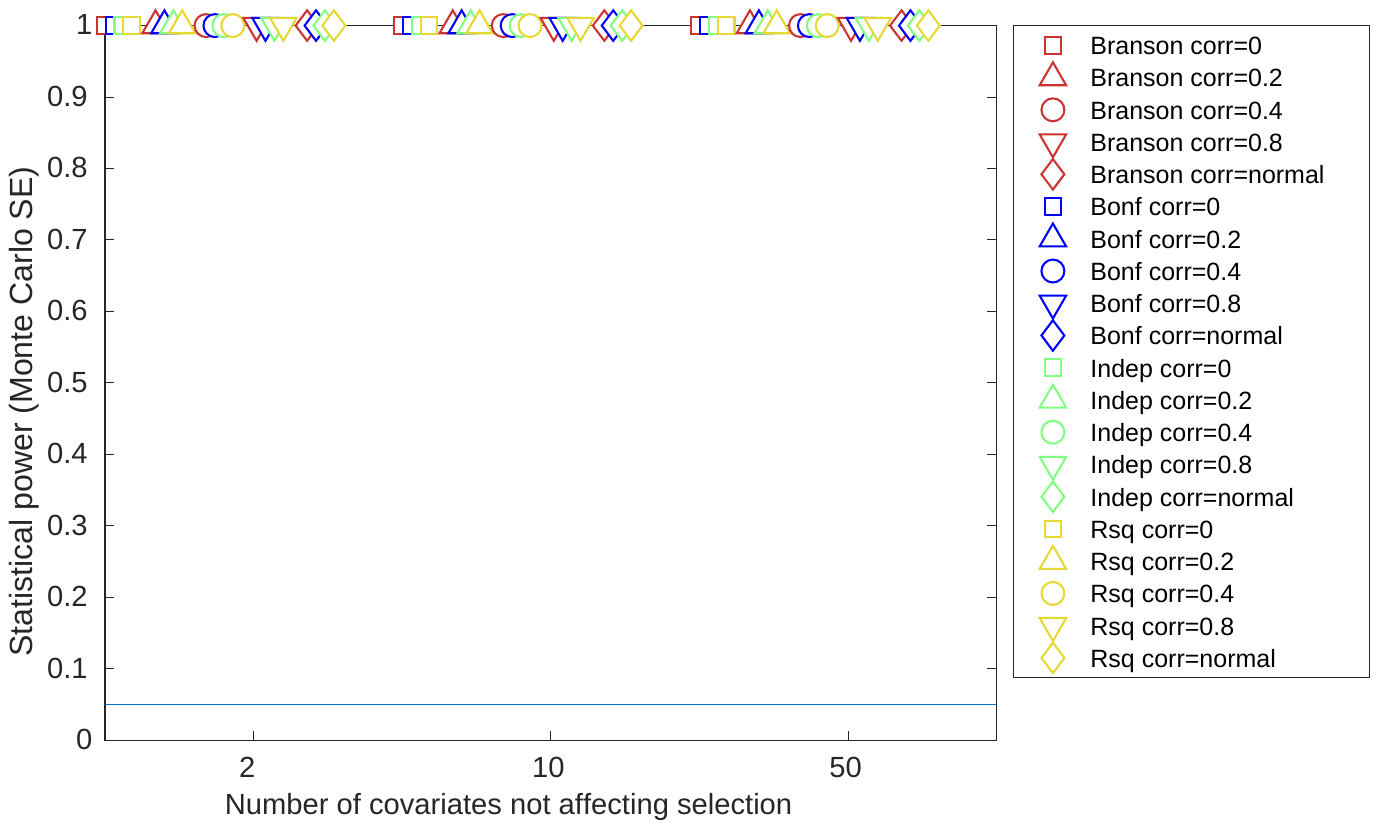 | 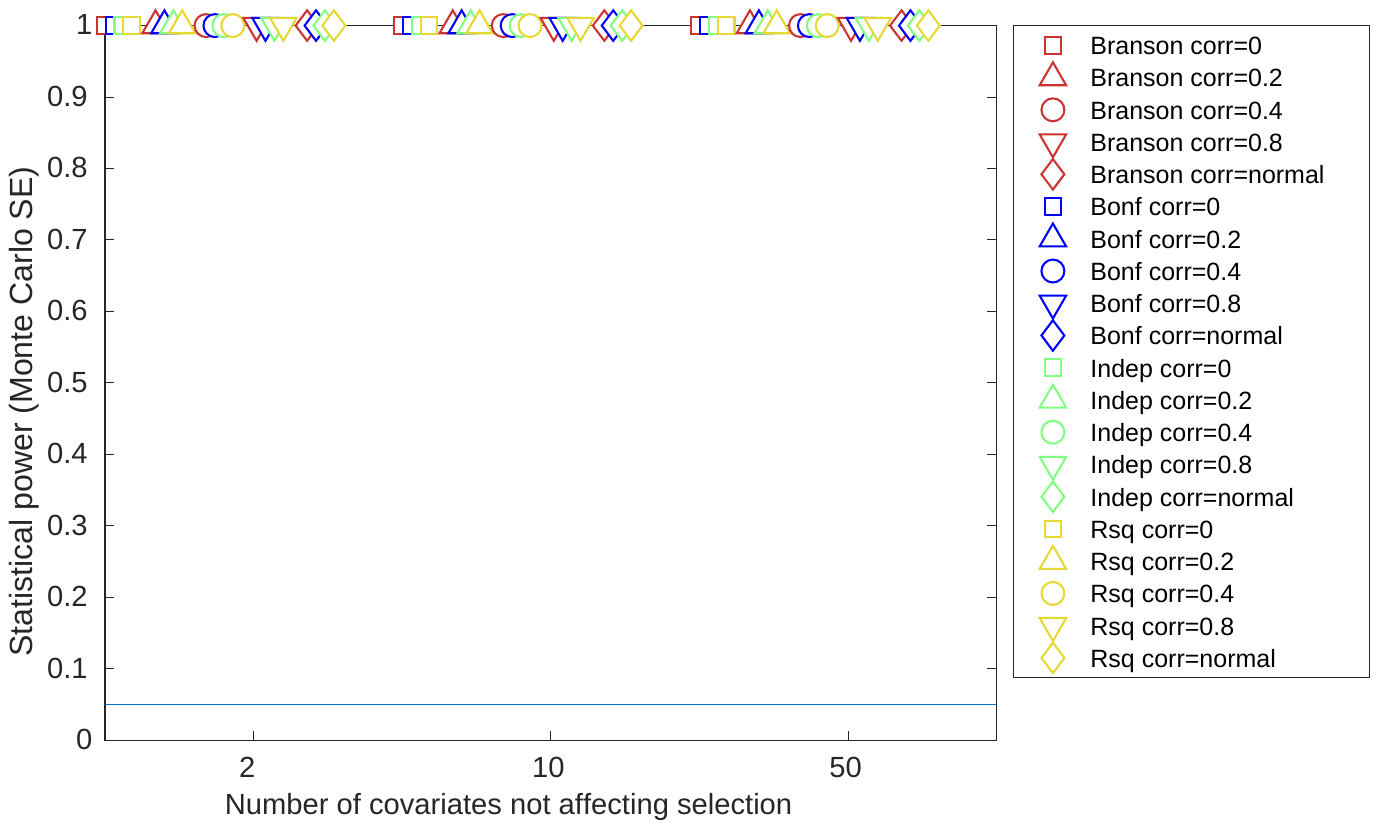 | 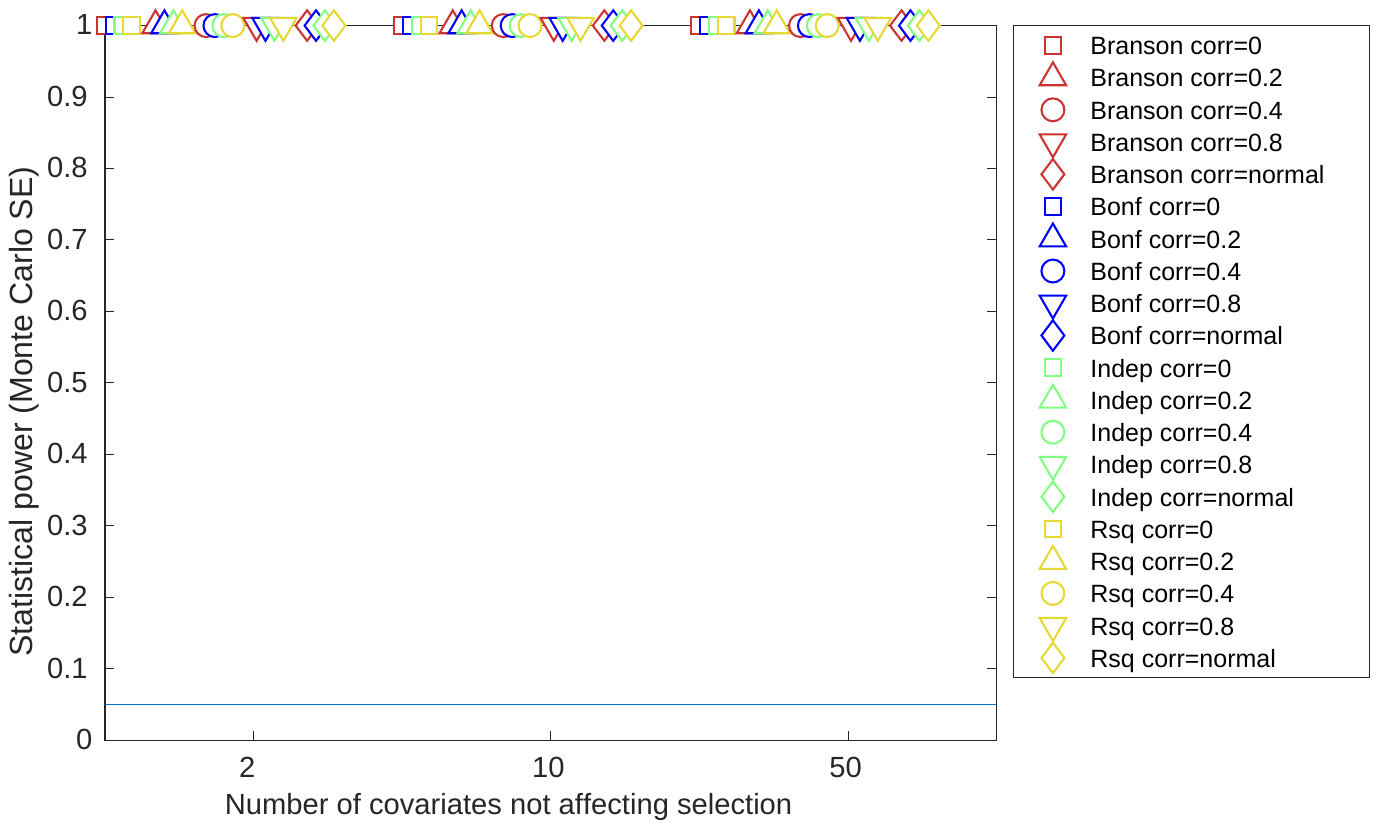 |

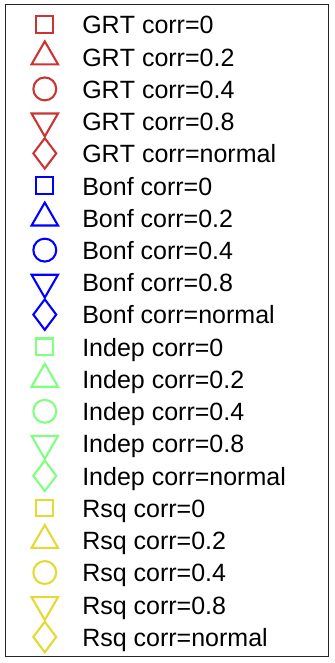
c) Instrument strength rsq=0.1, half covariates included in test

| **Total effect on selection** | $\boldsymbol{N}_{\boldsymbol{cs}}\boldsymbol{=2}$ | $\boldsymbol{N}_{\boldsymbol{cs}}\boldsymbol{=10}$ | $\boldsymbol{N}_{\boldsymbol{cs}}\boldsymbol{=50}$ |
| --- | --- | --- | --- |
| **R^2^=0.05** | 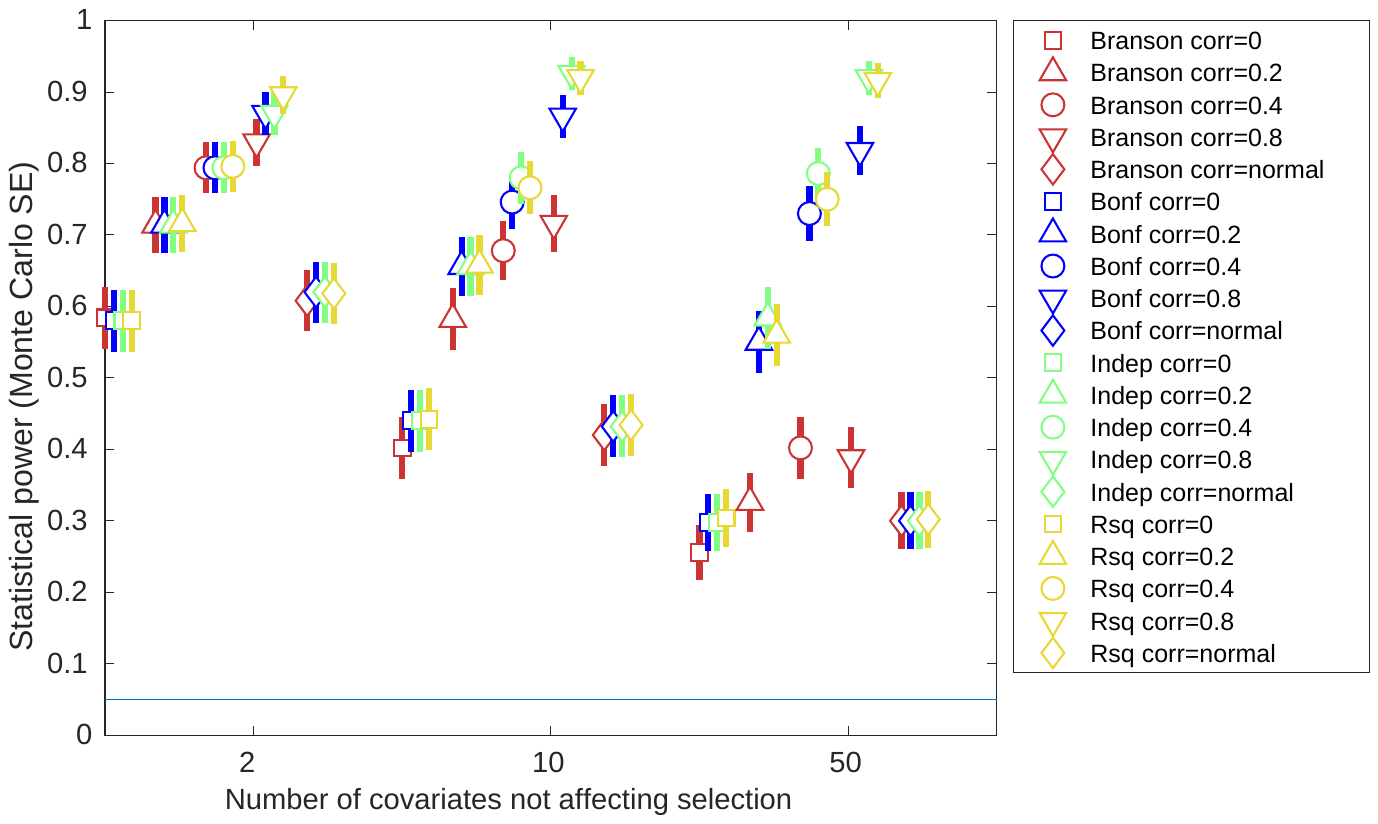 | 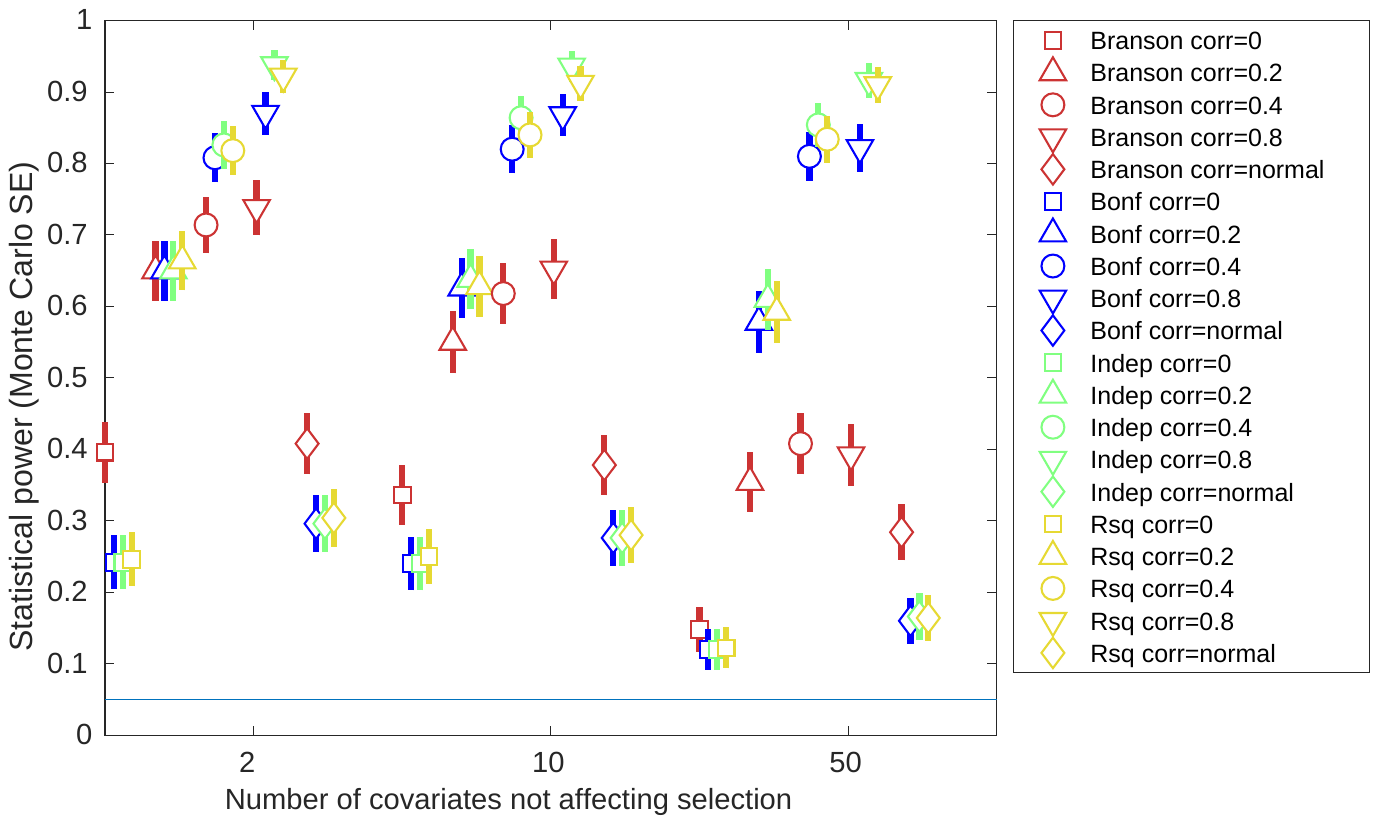 | 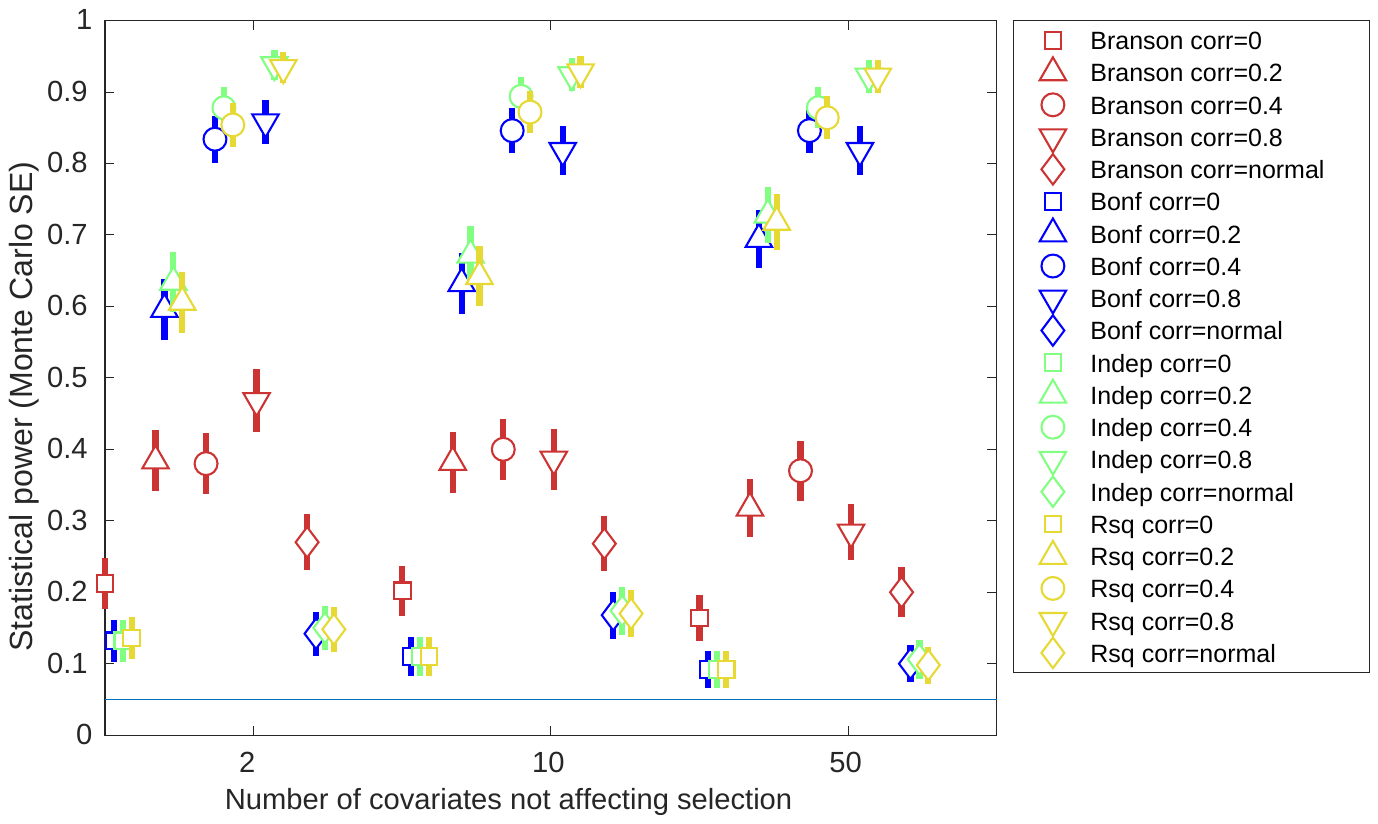 |
| **R^2^=0.1** | 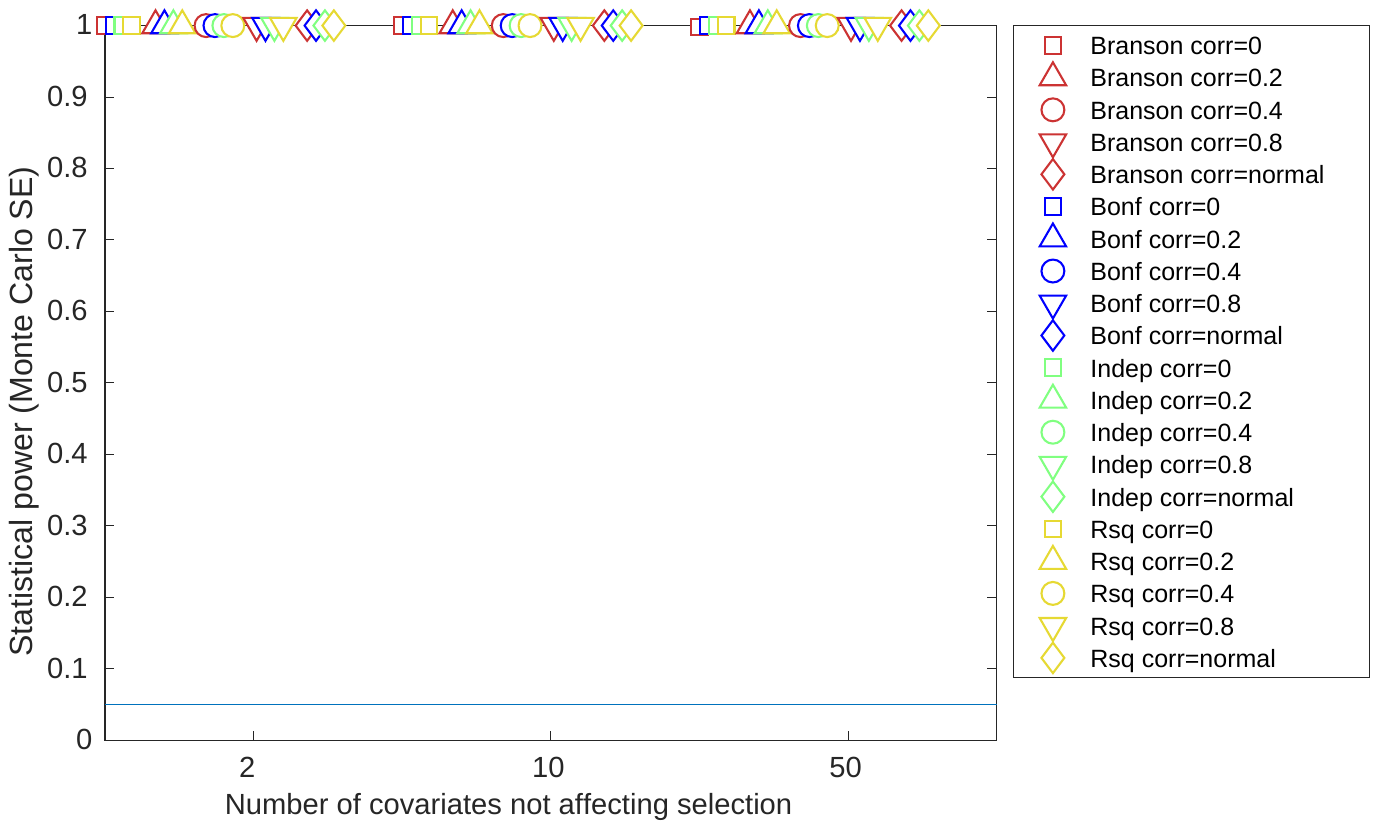 | 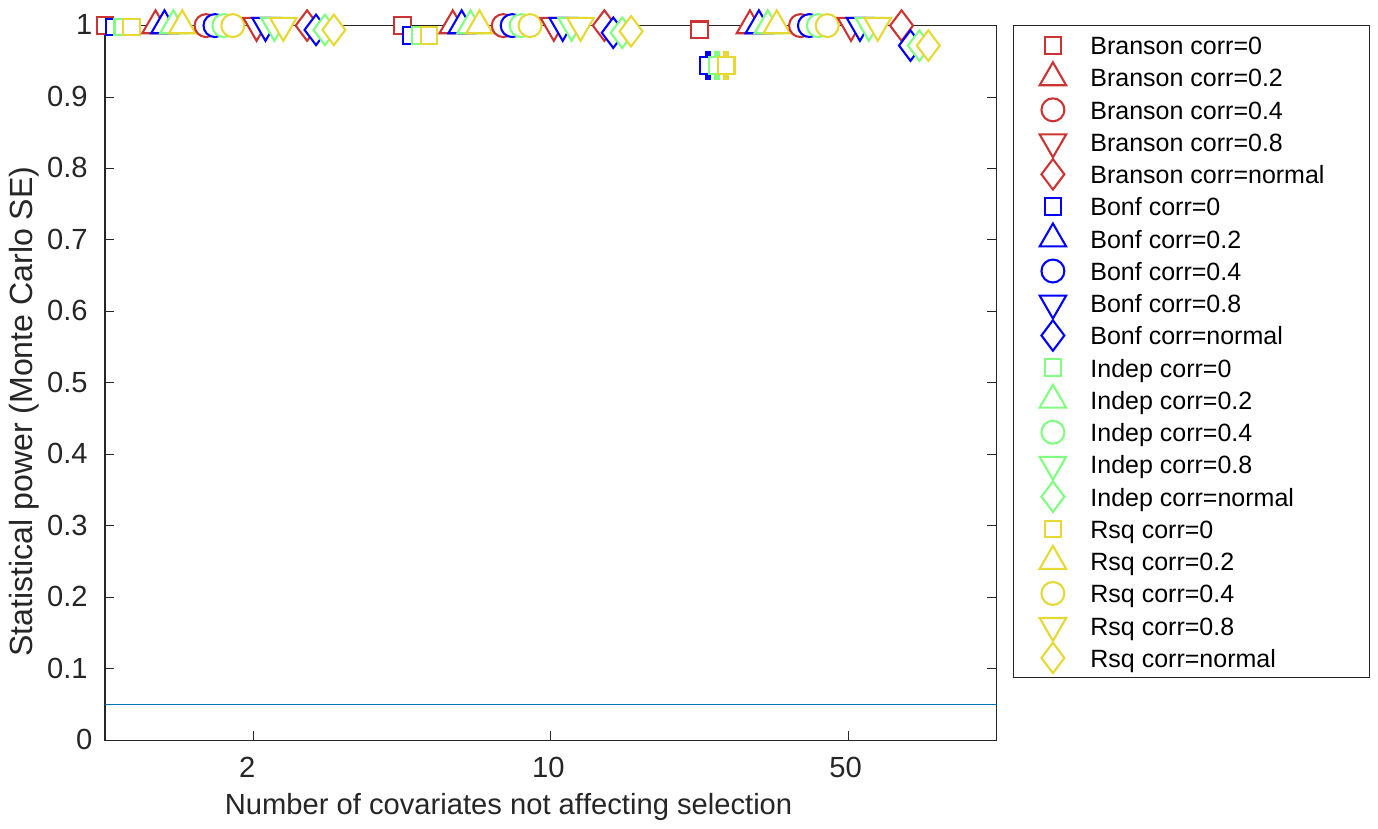 | 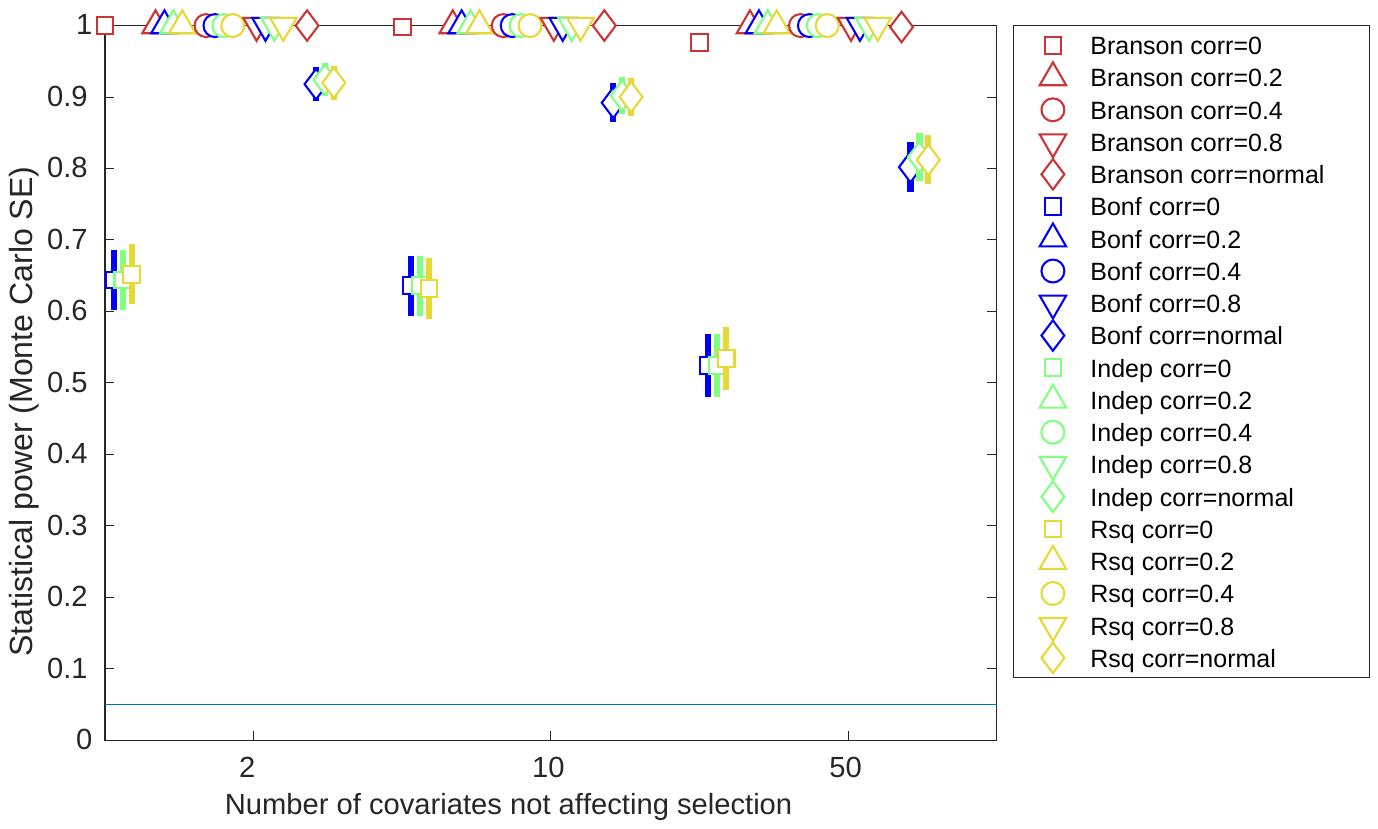 |
| **R^2^=0.2** | 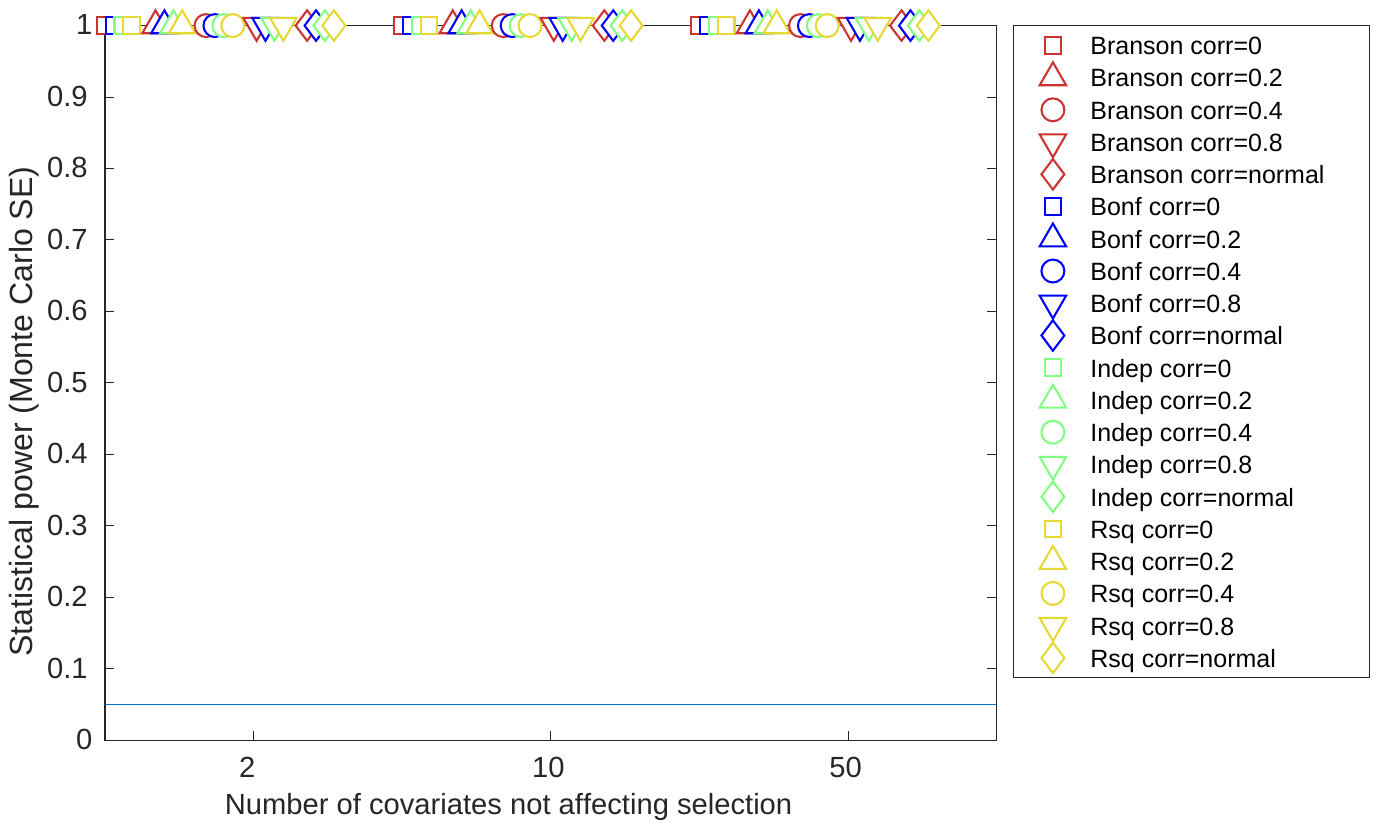 | 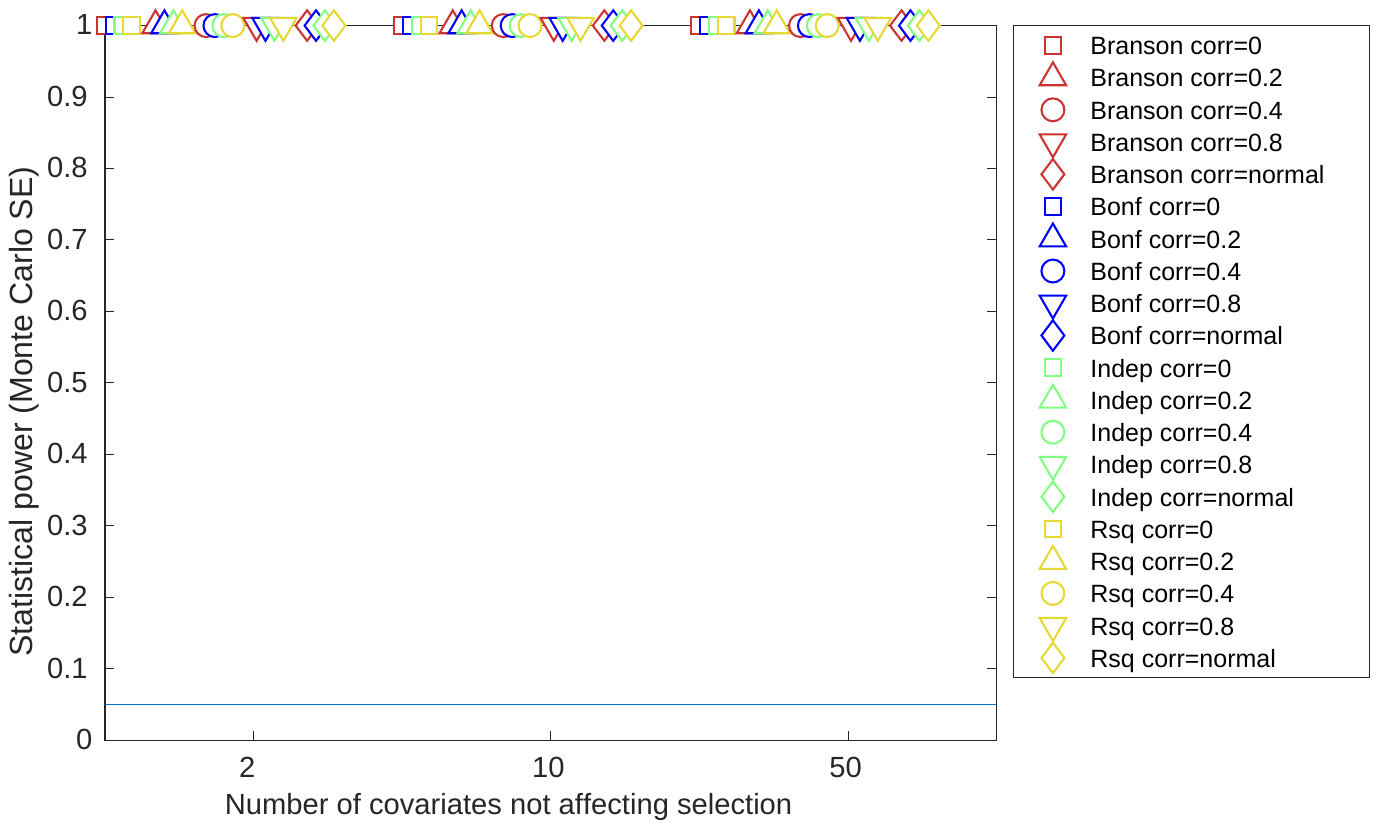 | 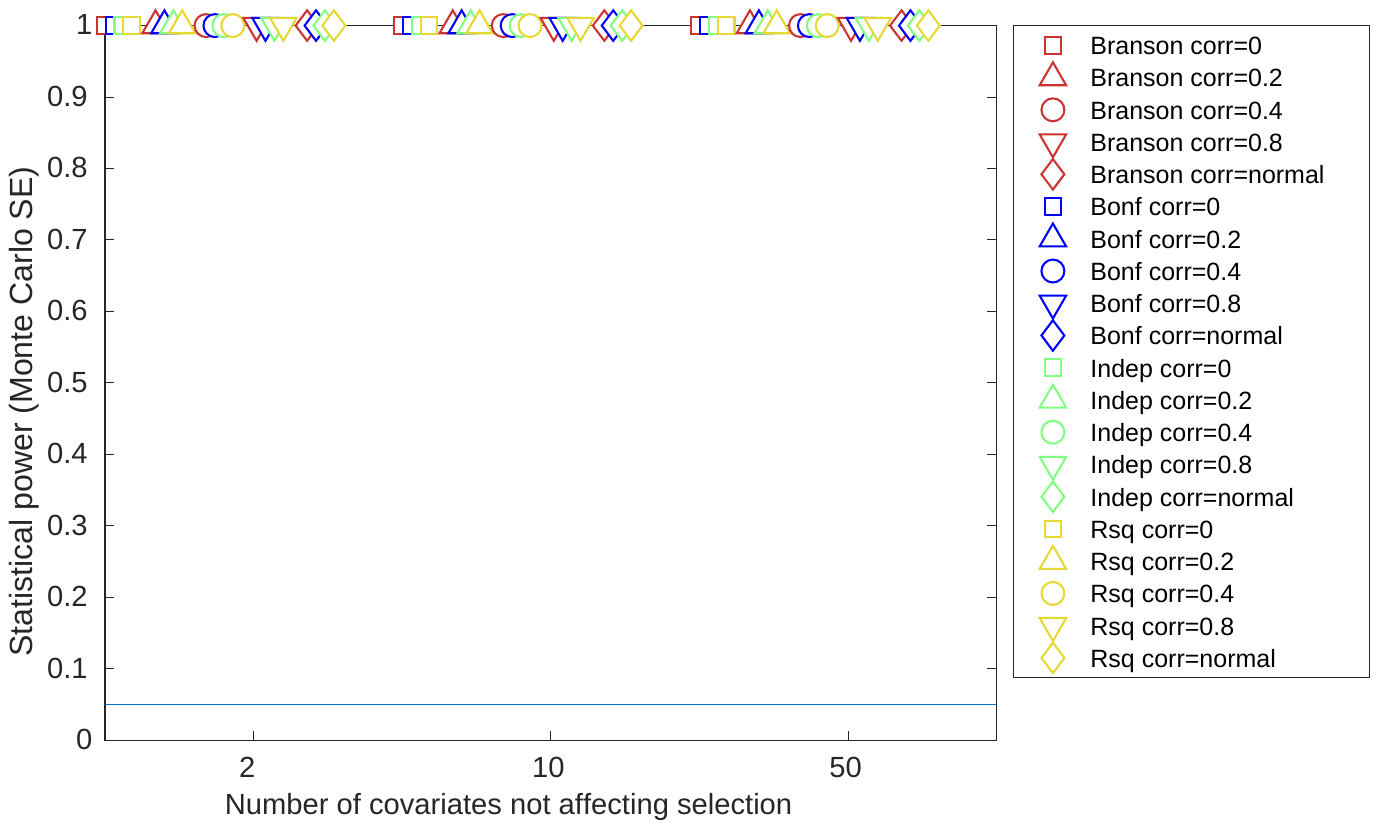 |

GRT: global randomization test; SE: standard error. Total effect on selection: the total effect of covariates C_S_ and X on selection S. $N_{cs}$**:** The number of Covariates affecting selection. Confidence intervals are +/- 1.96*MCSE (Monte Carlo standard error).

#### Supplementary figure 4: Results of selection bias simulations for instrument strength r^2^=0.05 and all covariates included in test, using the whole sample

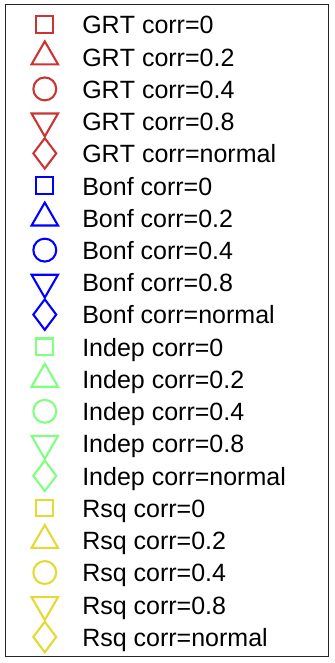

| **Total effect on selection** | $\boldsymbol{N}_{\boldsymbol{cs}}\boldsymbol{=2}$ | $\boldsymbol{N}_{\boldsymbol{cs}}\boldsymbol{=10}$ | $\boldsymbol{N}_{\boldsymbol{cs}}\boldsymbol{=50}$ |
| --- | --- | --- | --- |
| **R^2^=0.05** | 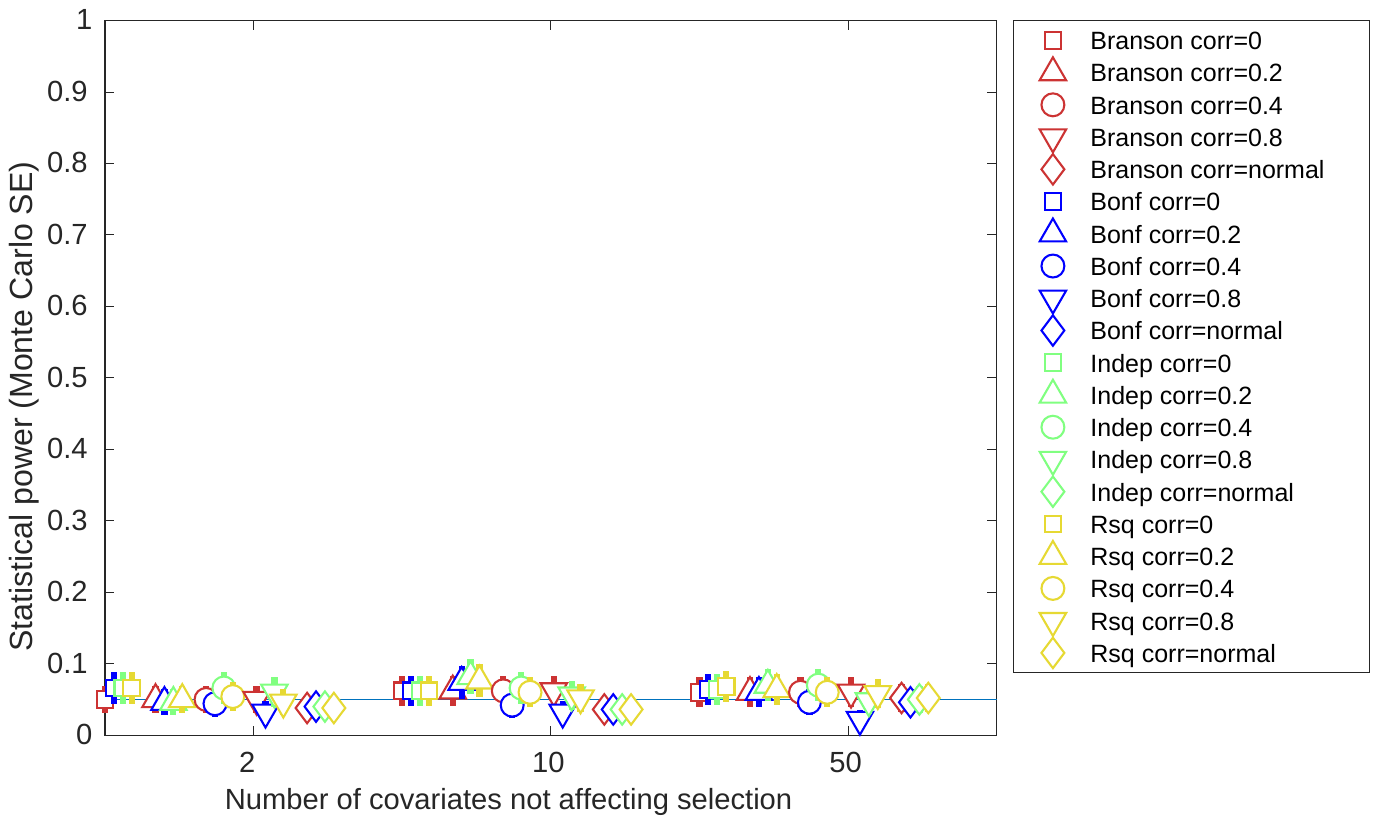 | 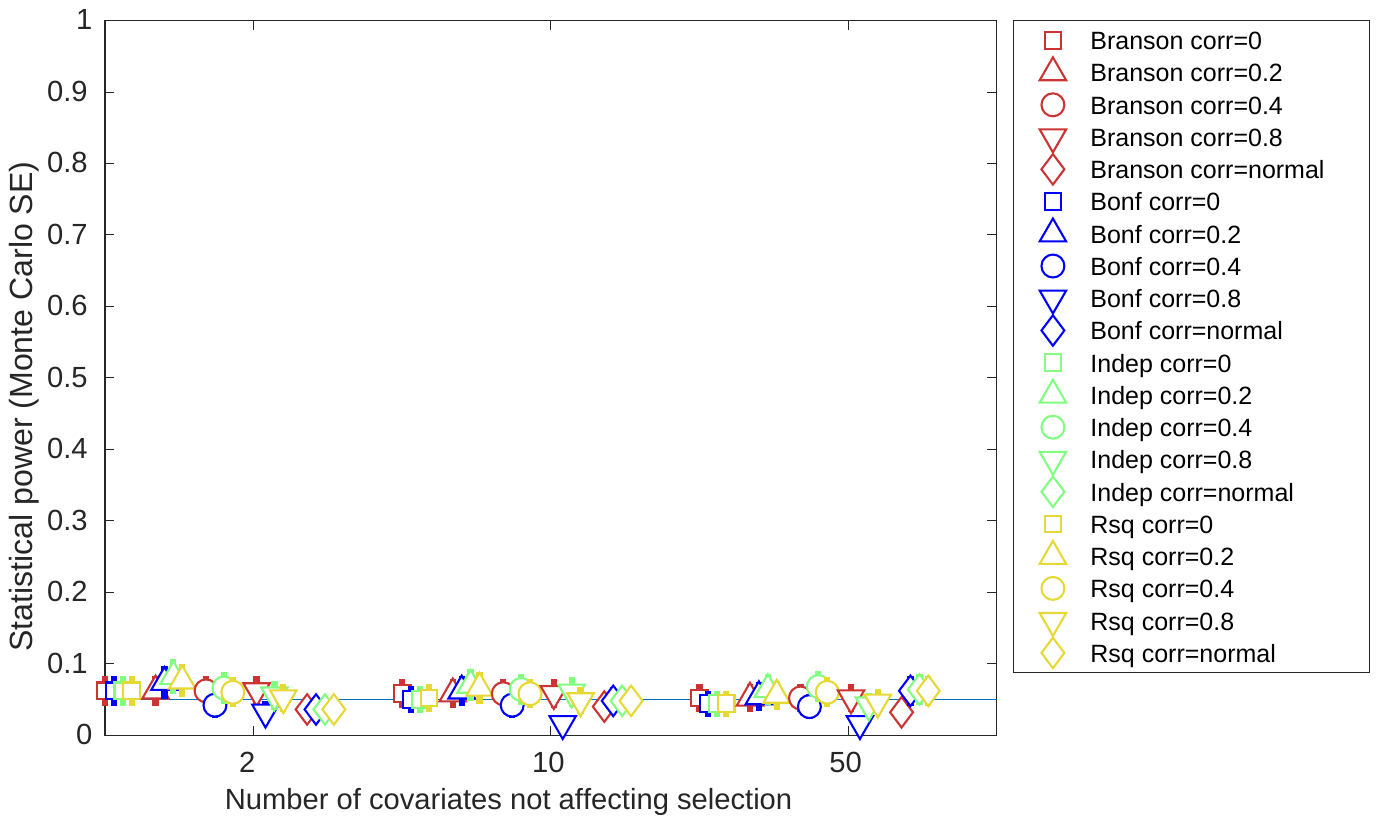 | 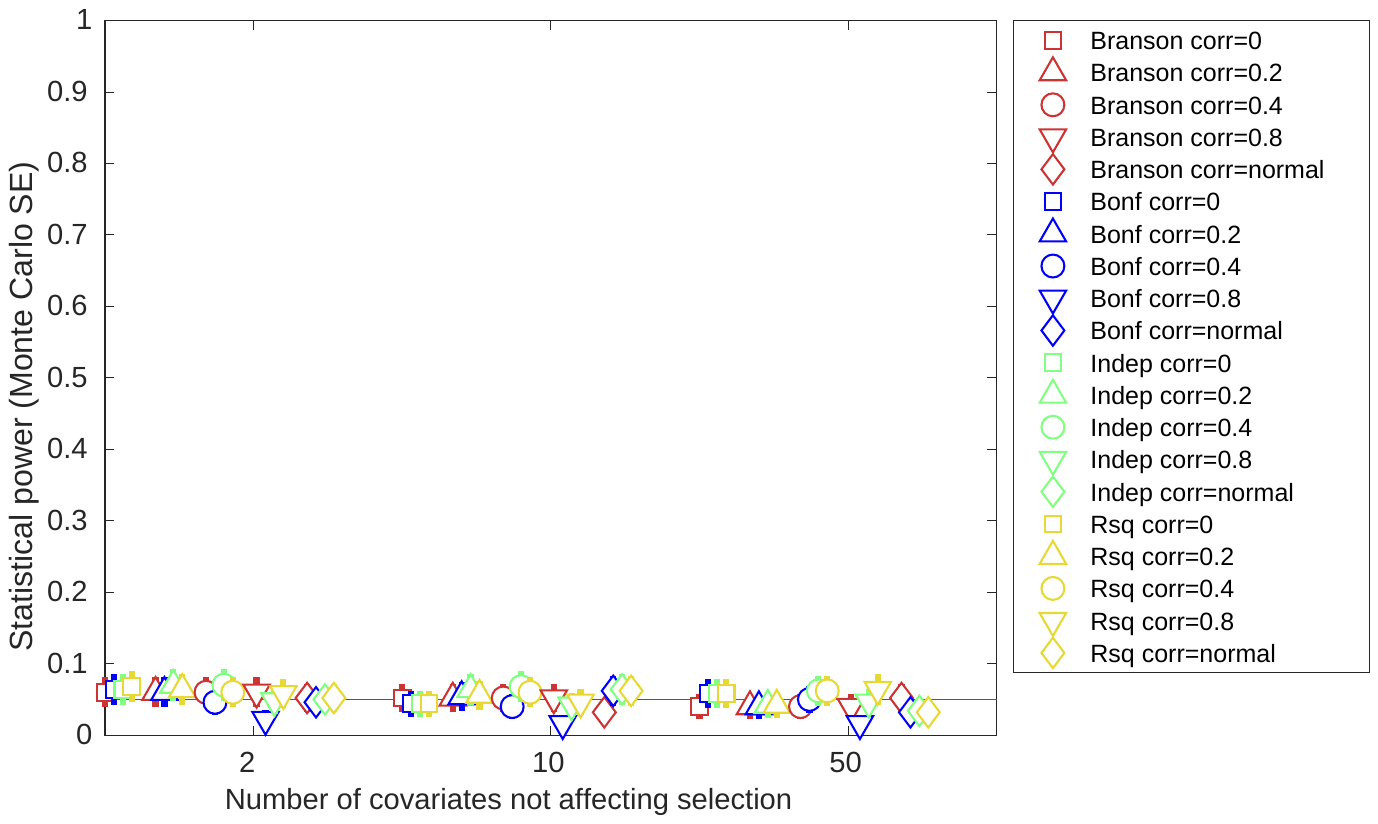 |
| **R^2^=0.1** | 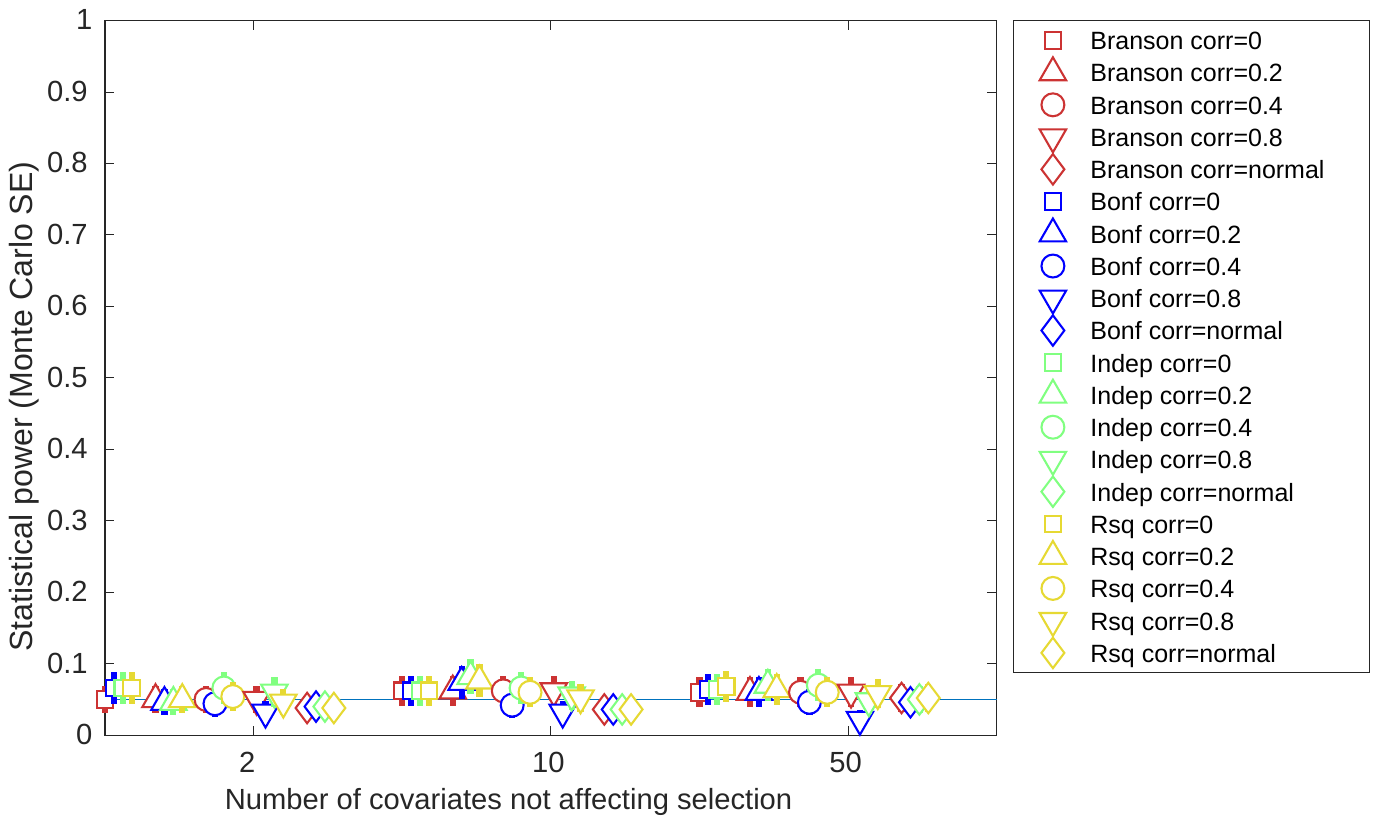 | 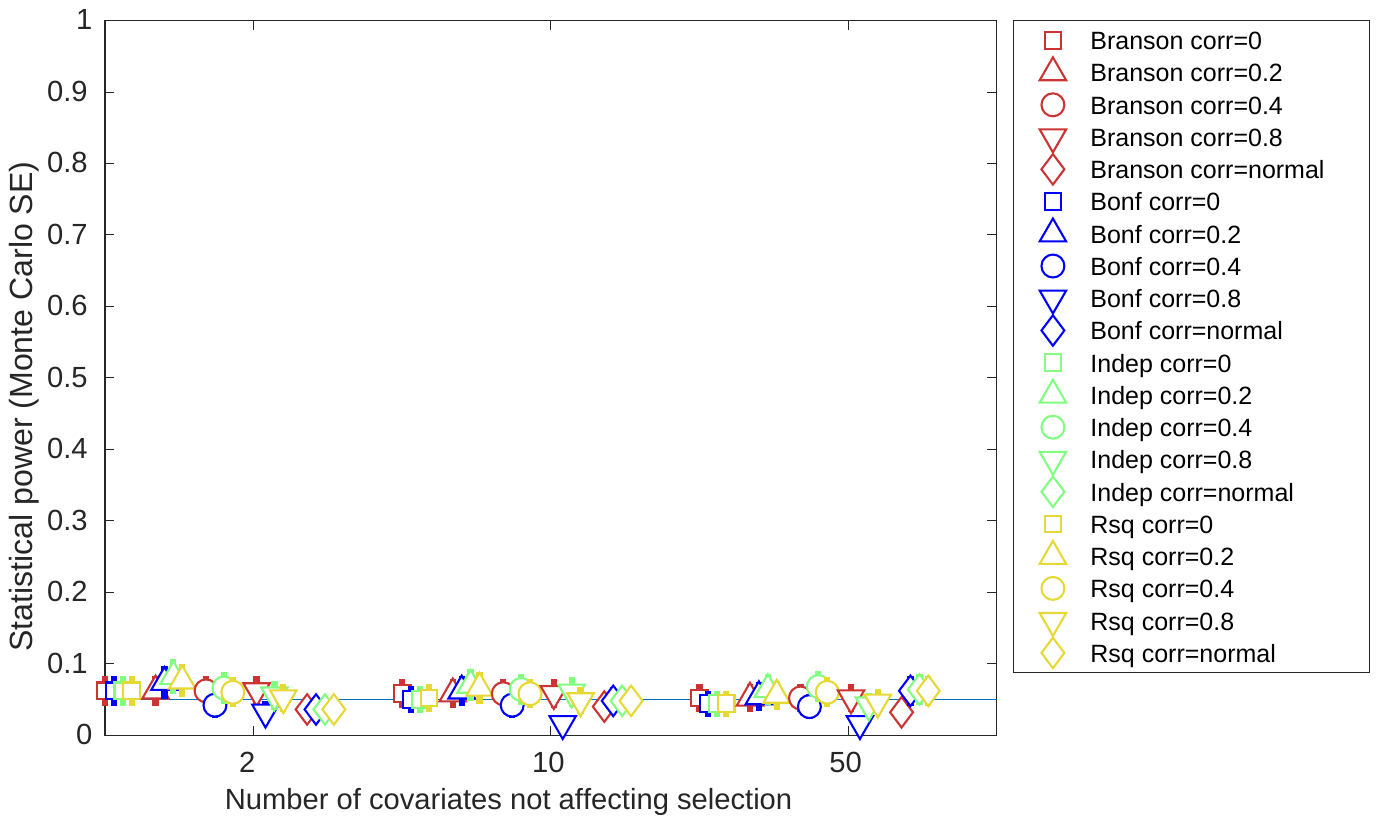 | 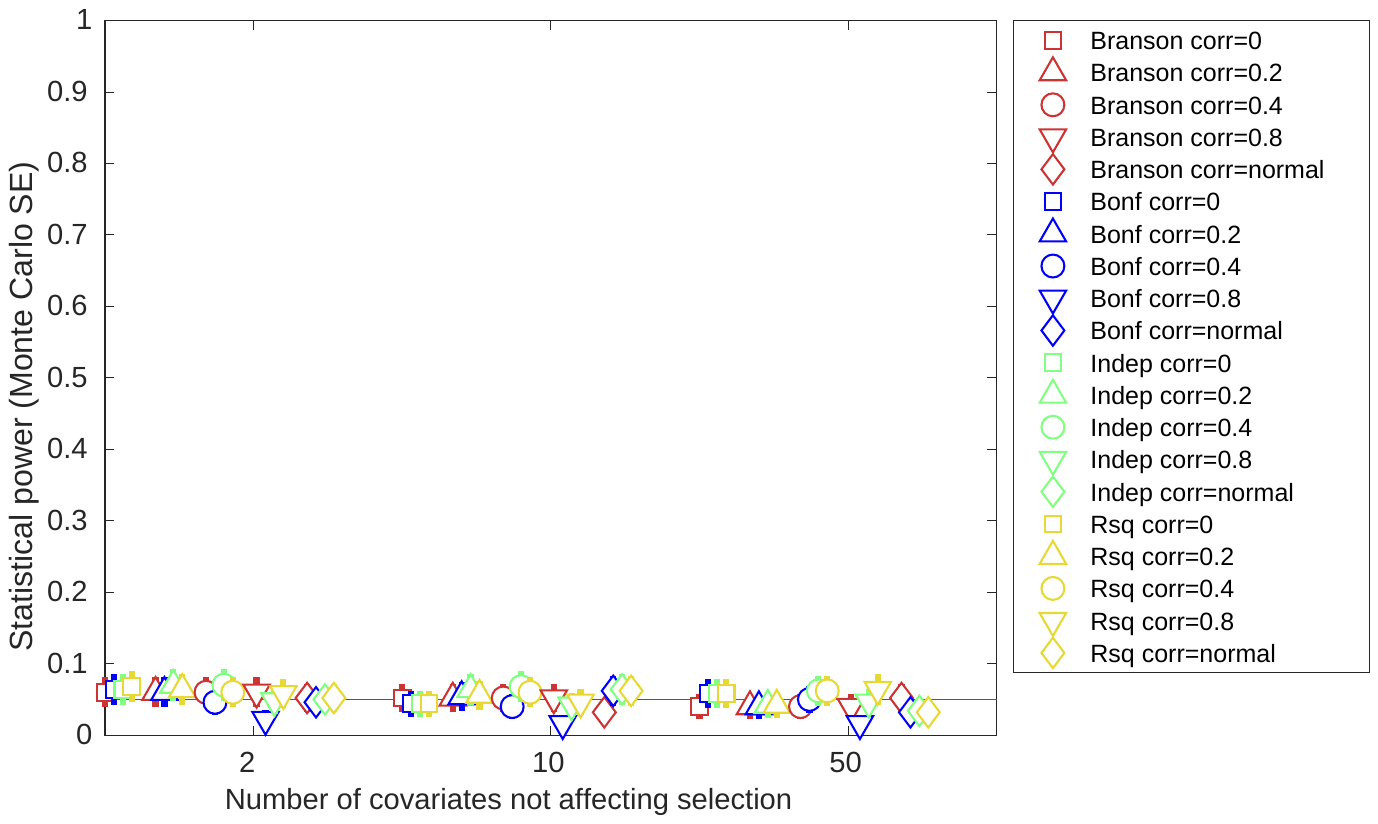 |
| **R^2^=0.2** | 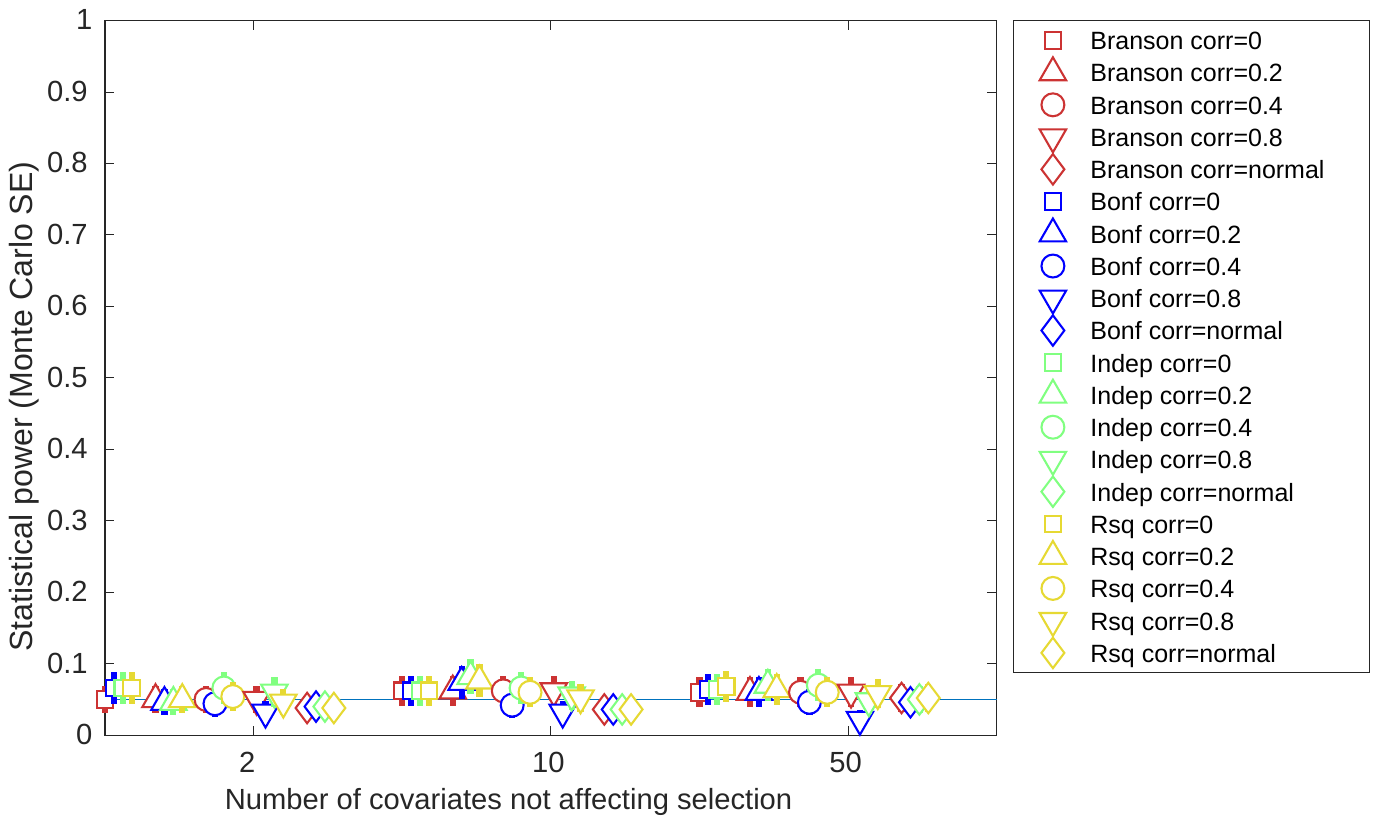 | 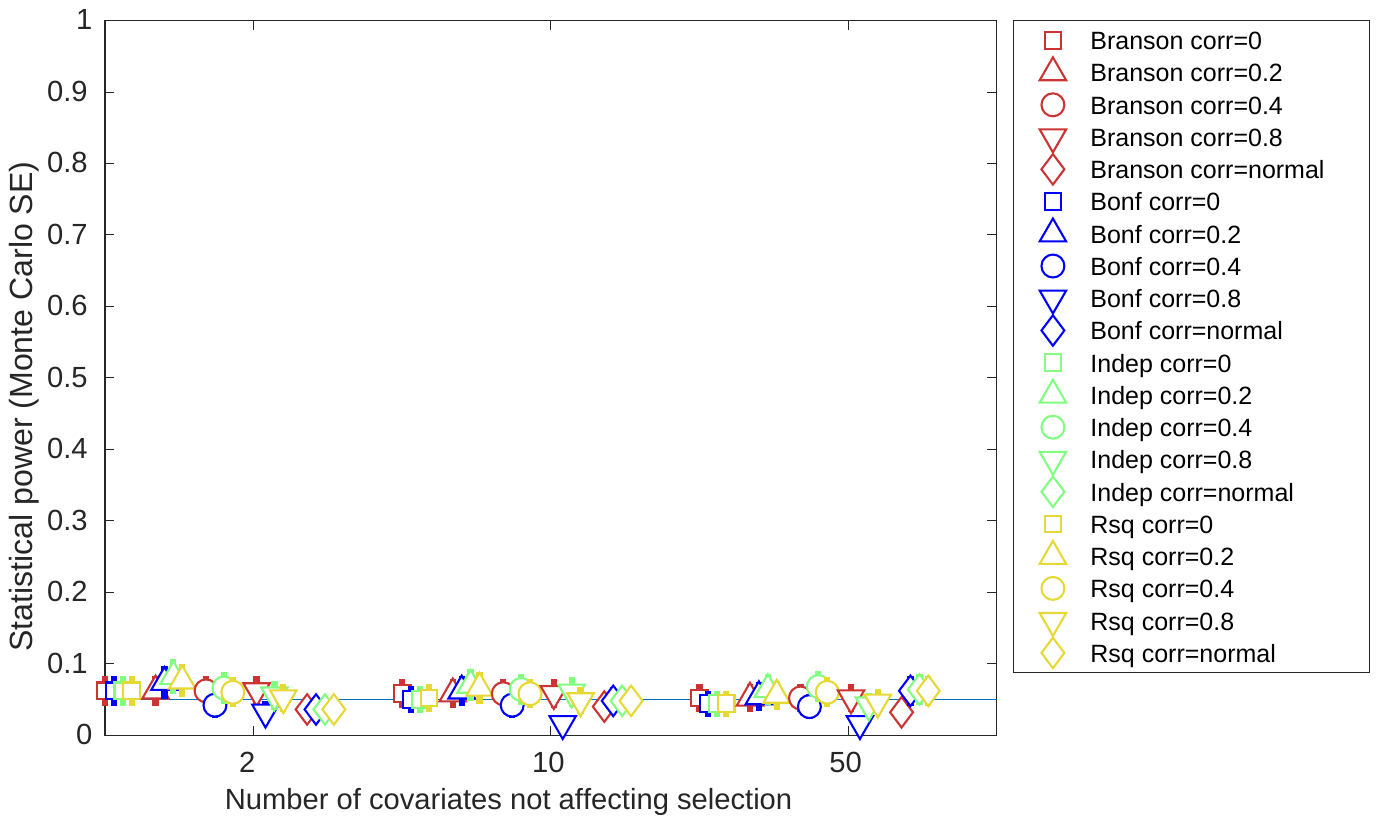 | 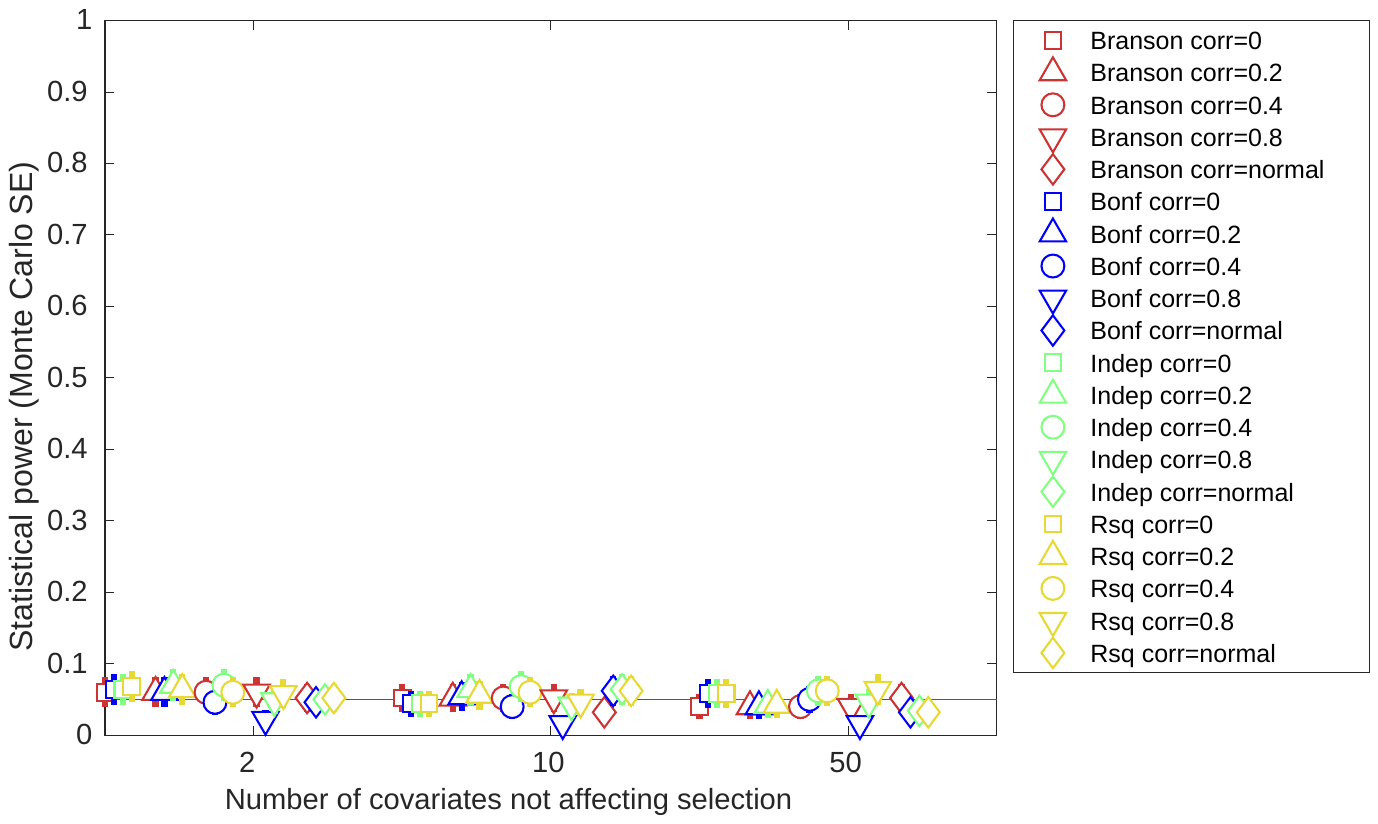 |

GRT: global randomization test; SE: standard error. Total effect on selection: the total effect of covariates C_S_ and X on selection S. $N_{cs}$**:** The number of Covariates affecting selection. Confidence intervals are +/- 1.96*MCSE (Monte Carlo standard error).

#### Supplementary figure 5: Covariate x Exposure interaction sizes for selection bias simulations with instrument strength r^2^=0.05

| **Total effect on selection** | $\boldsymbol{N}_{\boldsymbol{cs}}\boldsymbol{=2}$ | $\boldsymbol{N}_{\boldsymbol{cs}}\boldsymbol{=10}$ | $\boldsymbol{N}_{\boldsymbol{cs}}\boldsymbol{=50}$ |
| --- | --- | --- | --- |
| **R^2^=0.05** | 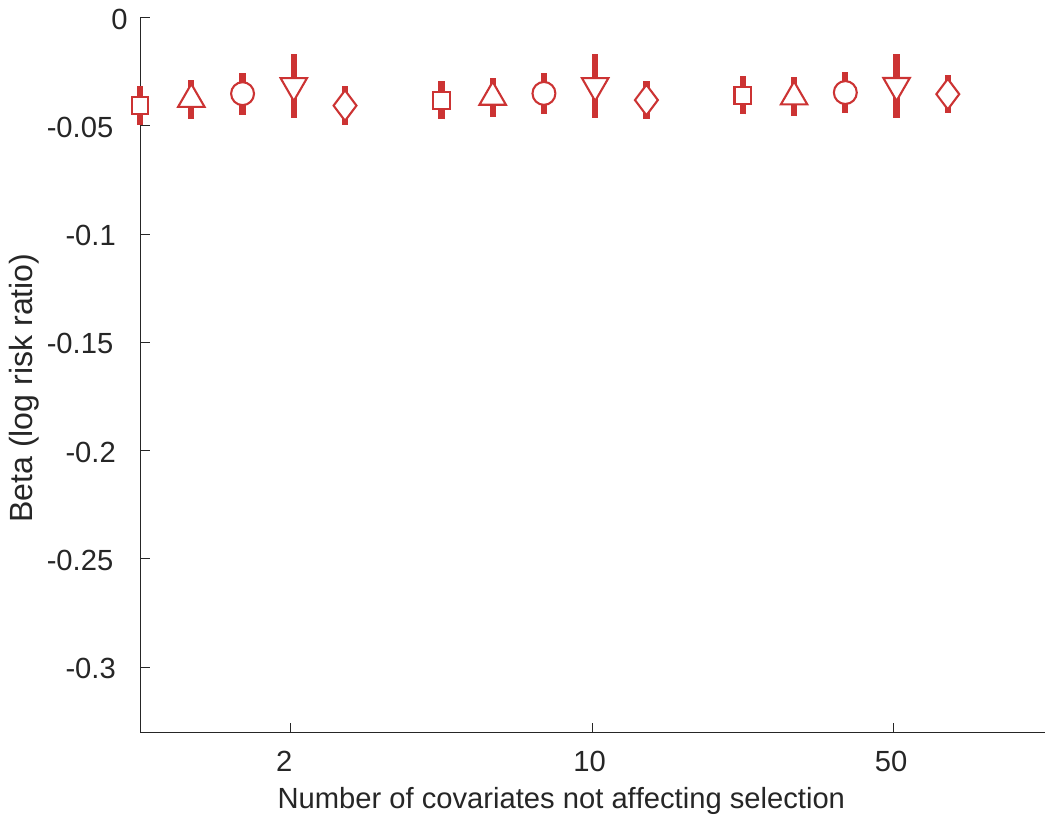 | 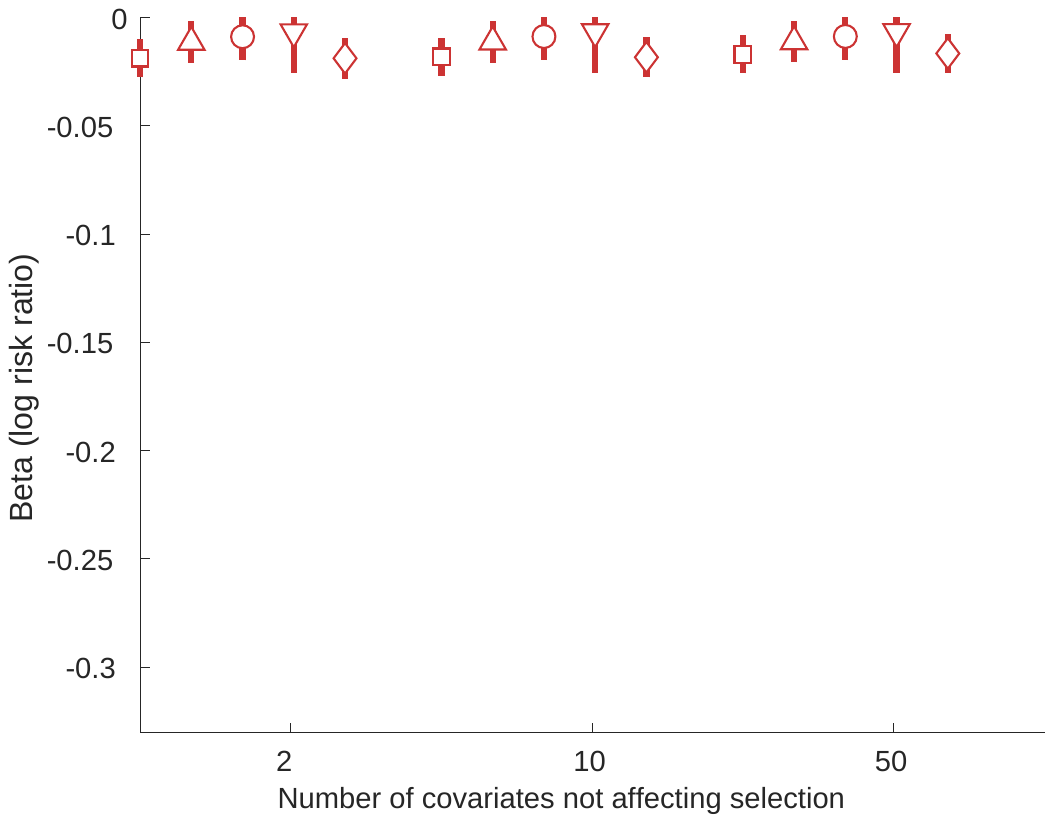 | 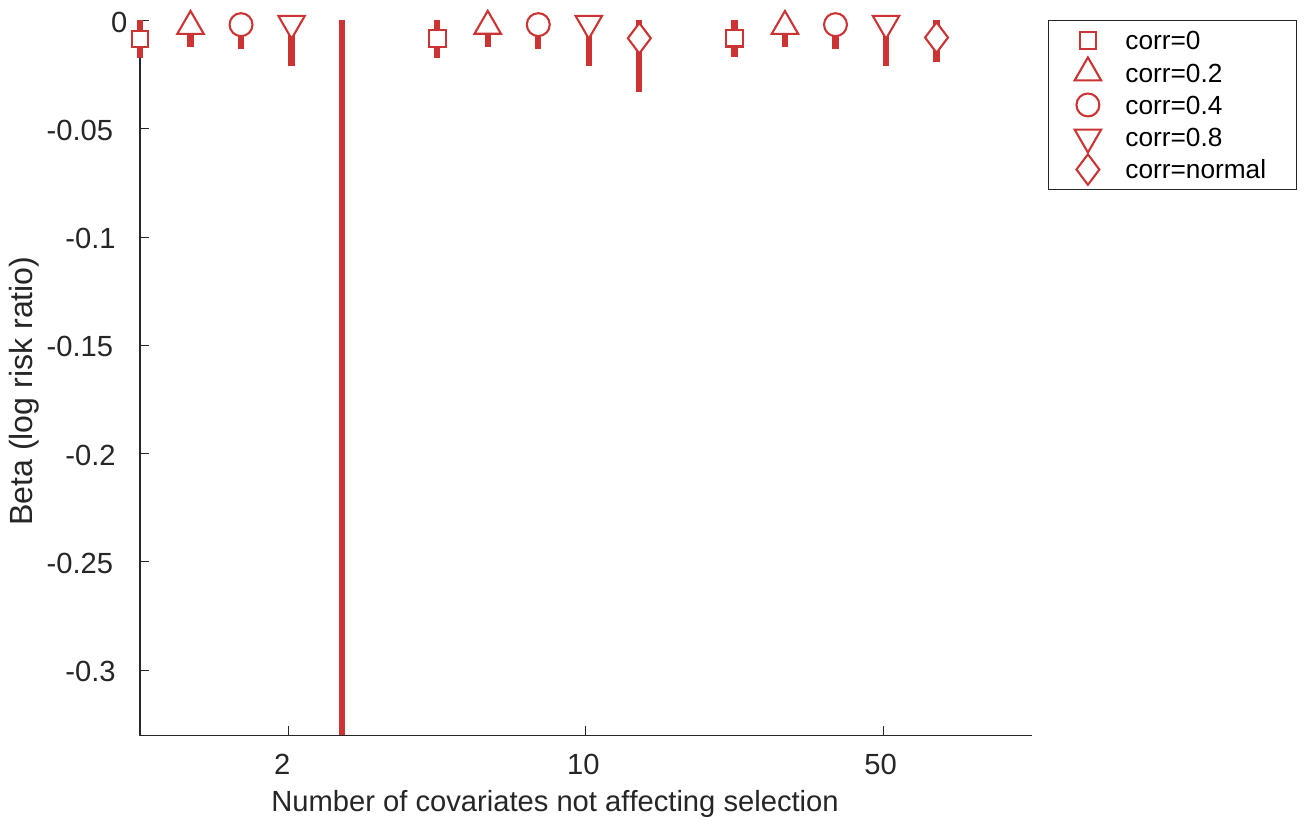 |
| **R^2^=0.1** | 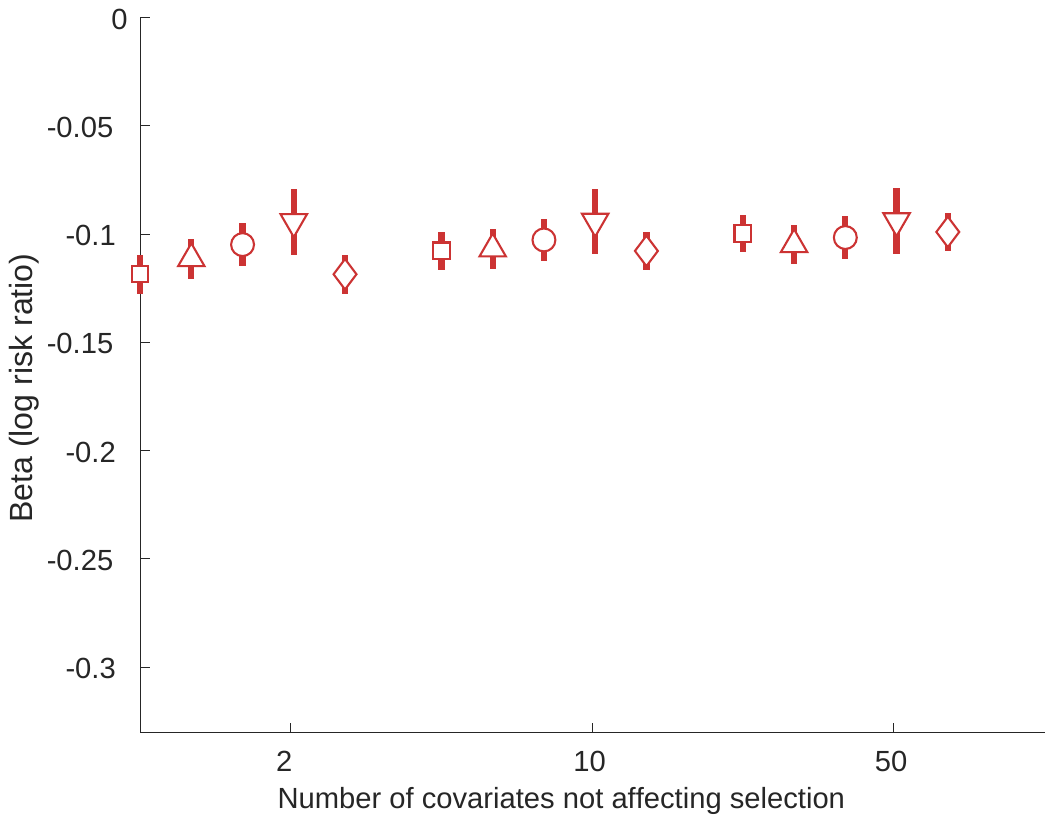 | 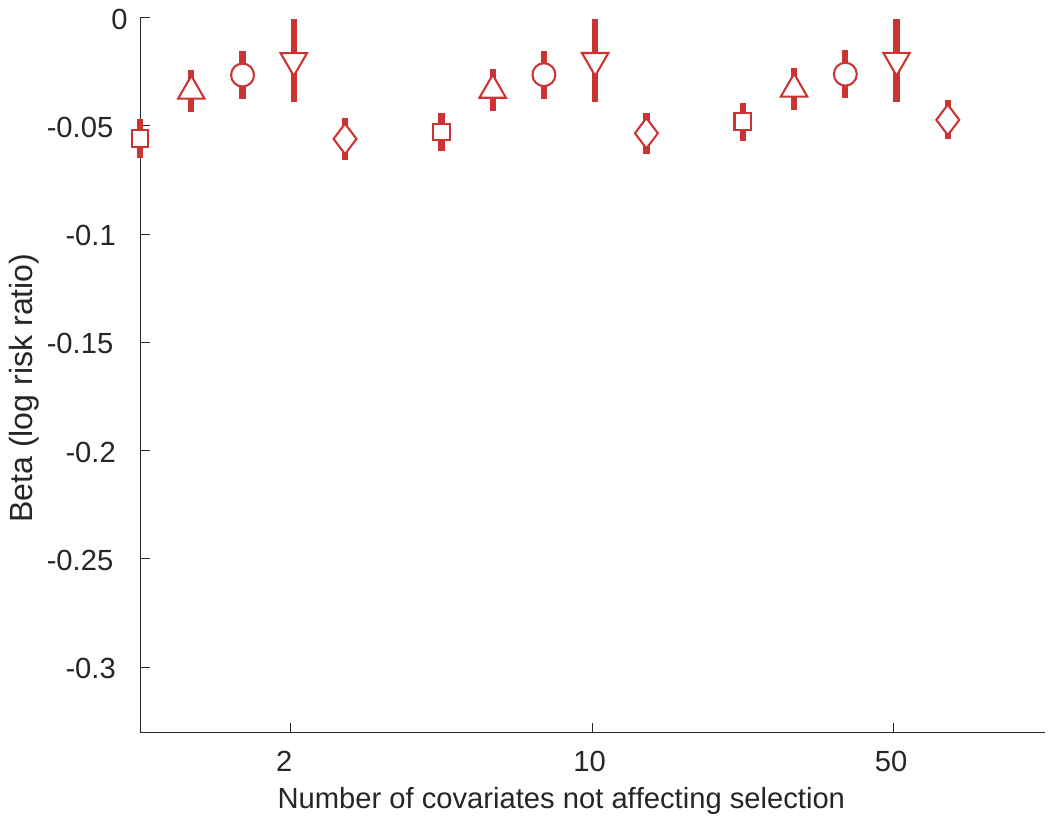 | 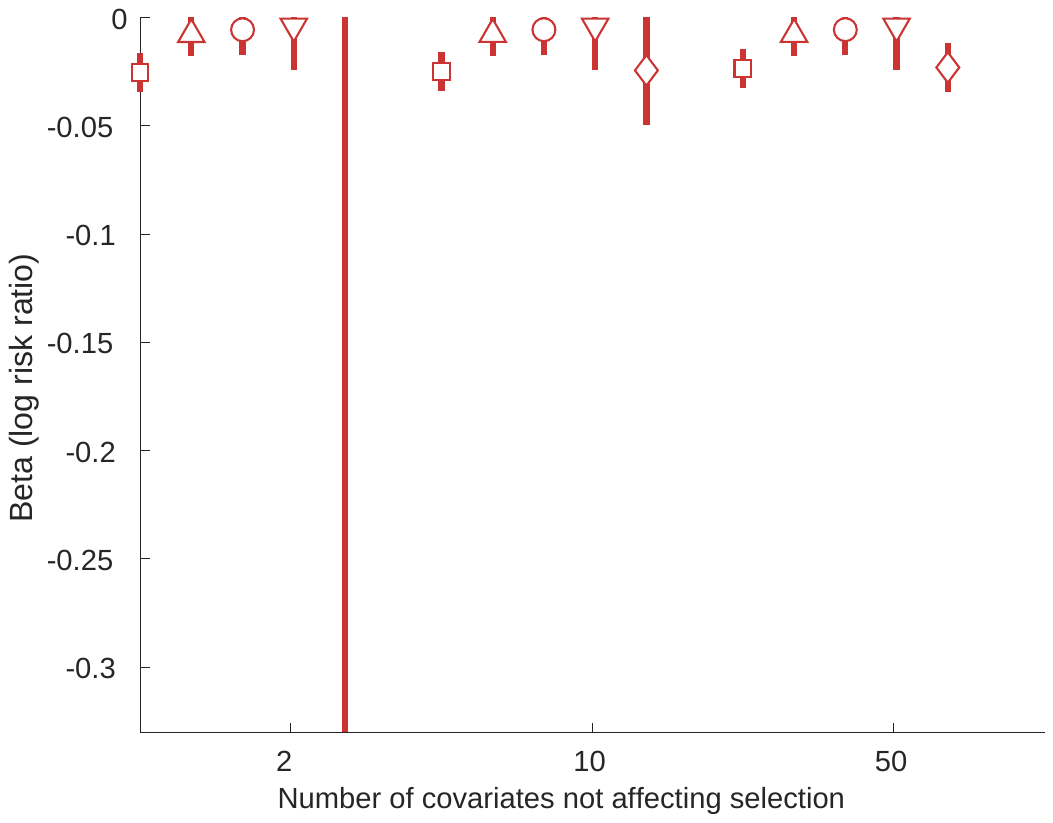 |
| **R^2^=0.2** | 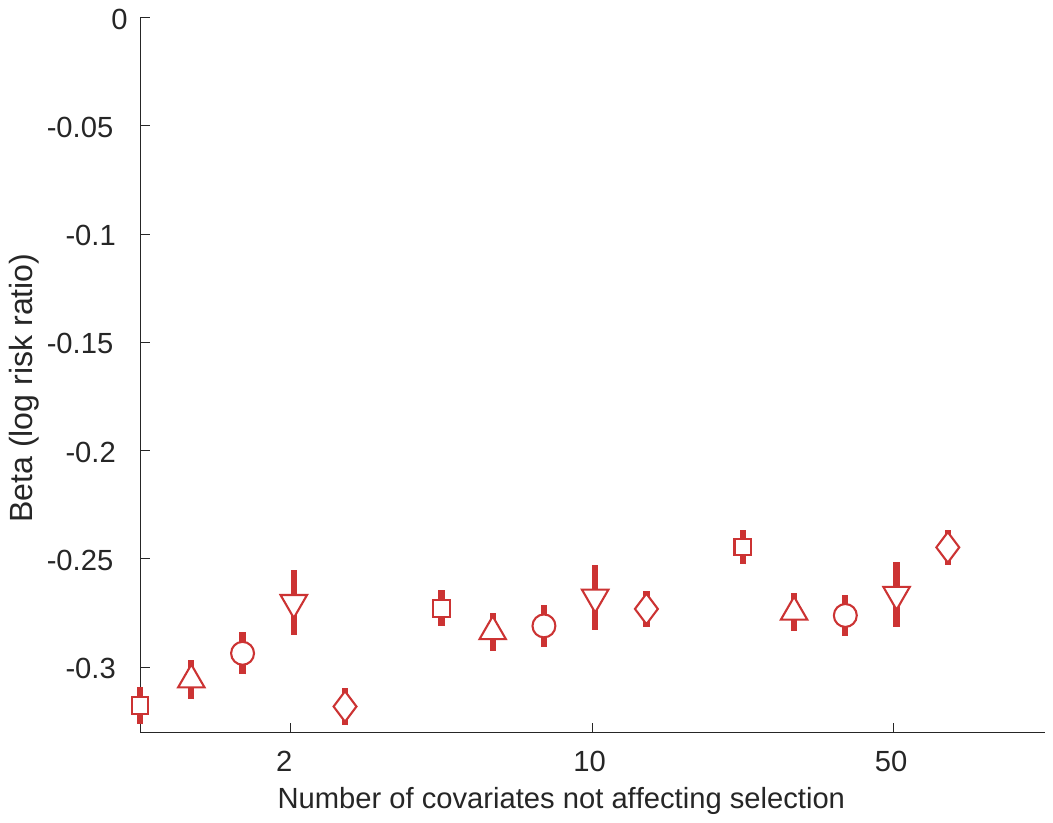 | 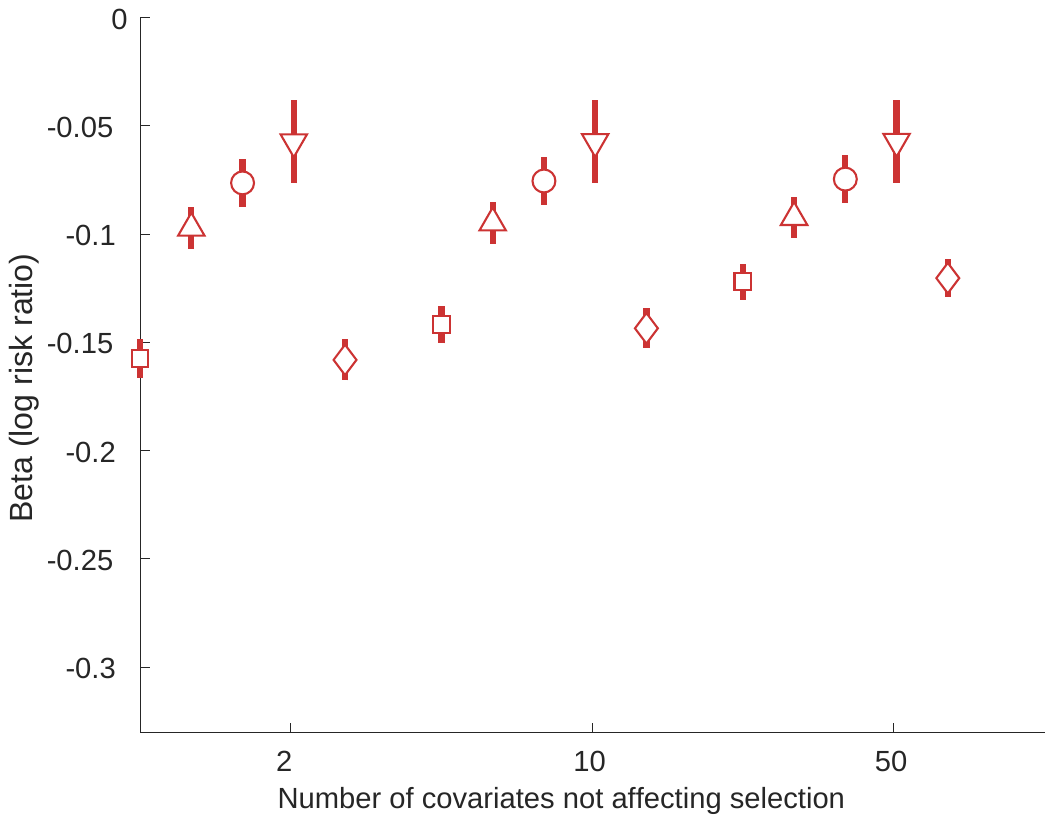 | 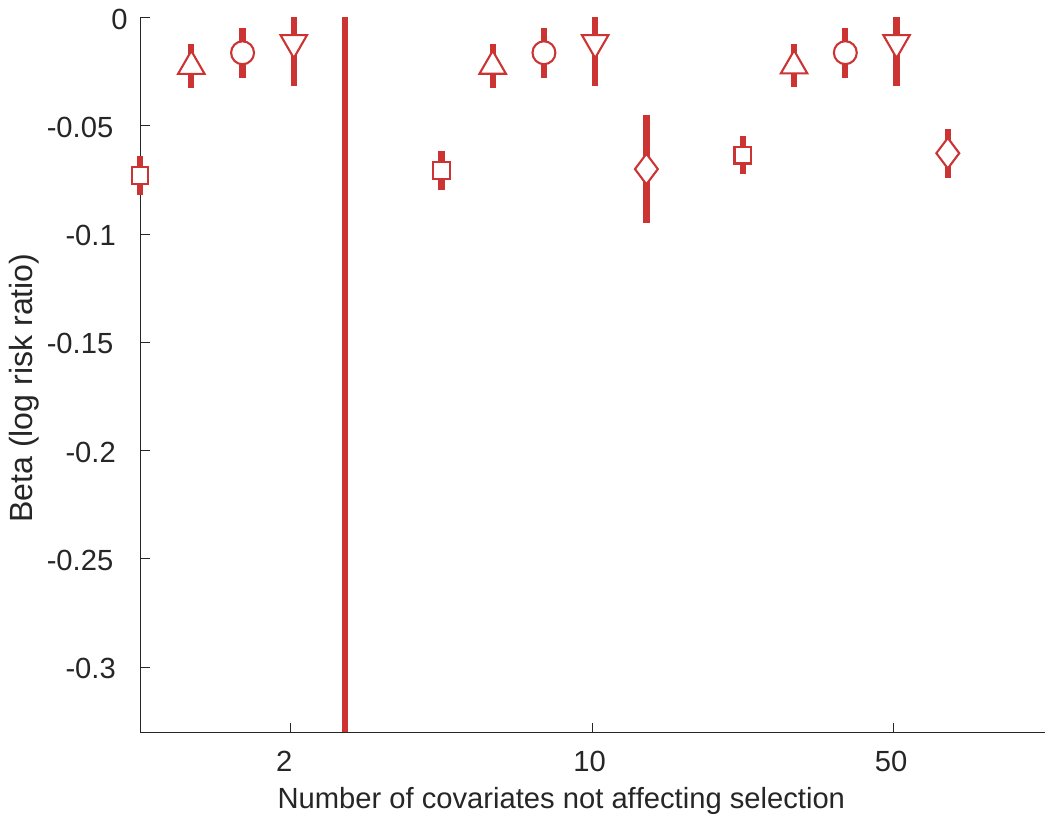 |

Average coefficient (1.96x model-based SE [i.e. average SE across all models]) for interaction terms in a poisson regression model estimating the association of covariates C_S_, and each of their interactions with exposure x, with selection. $N_{\mathrm{cs}}$**:** The number of Covariates affecting selection. Confidence intervals are +/- 1.96*MCSE (Monte Carlo standard error).

#### Supplementary figure 6: Results of horizontal pleiotropy simulations for non horizontally pleiotropic SNP (with effect of the horizontally pleiotropic SNP on the covariates set to r^2^=0.001

| a) SNP effect on each covariate R^2^= 0.001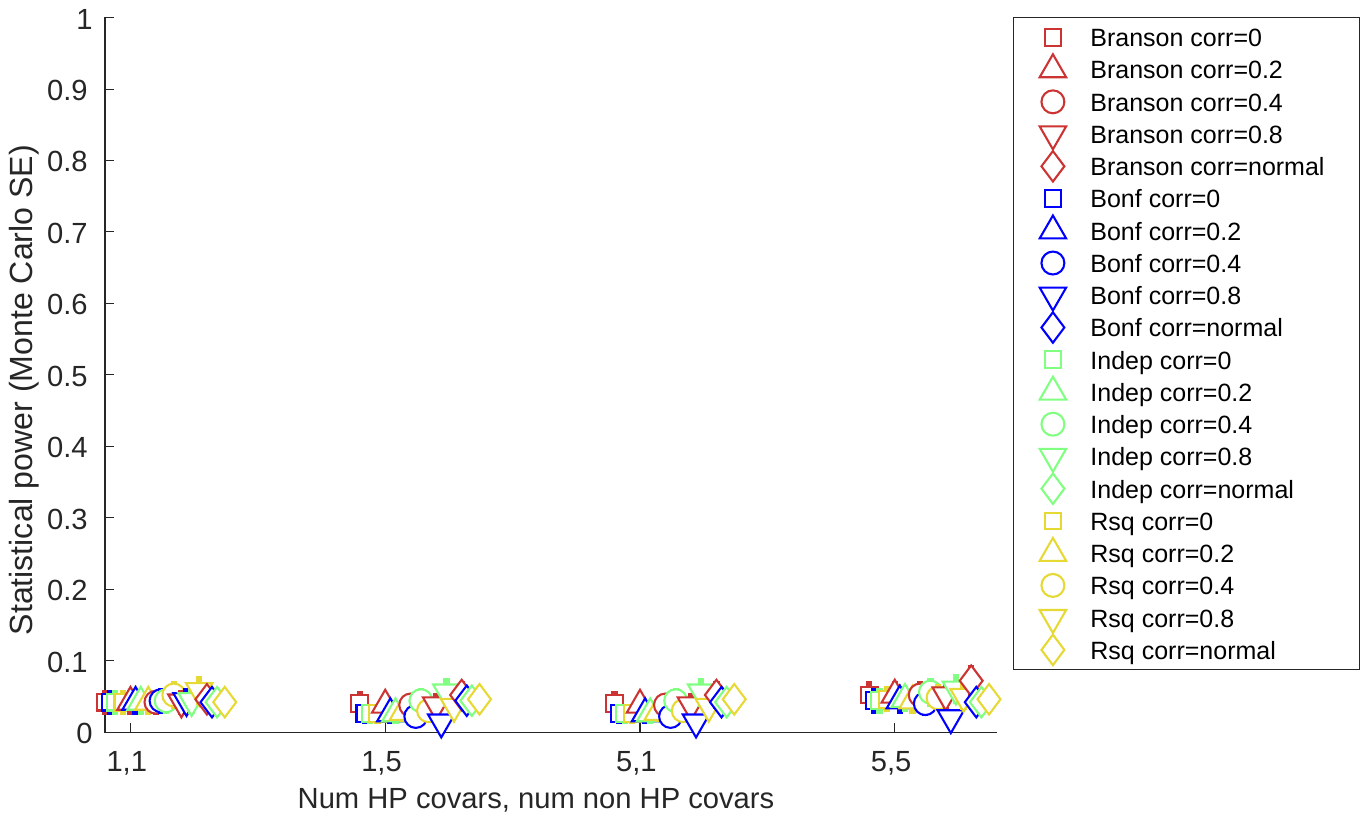 | b) SNP effect on each covariate R^2^= 0.005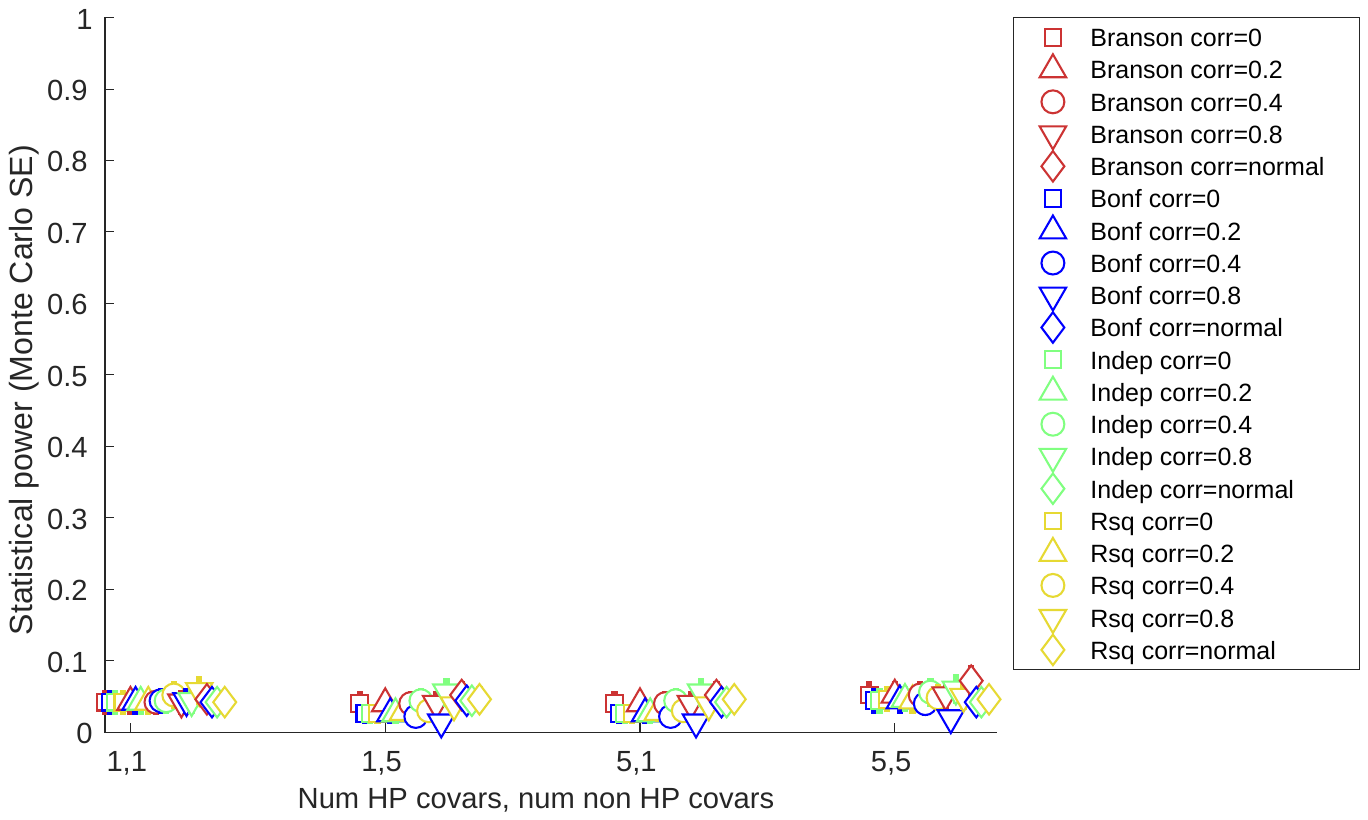 |
| --- | --- |
| c) SNP effect on each covariate R^2^= 0.01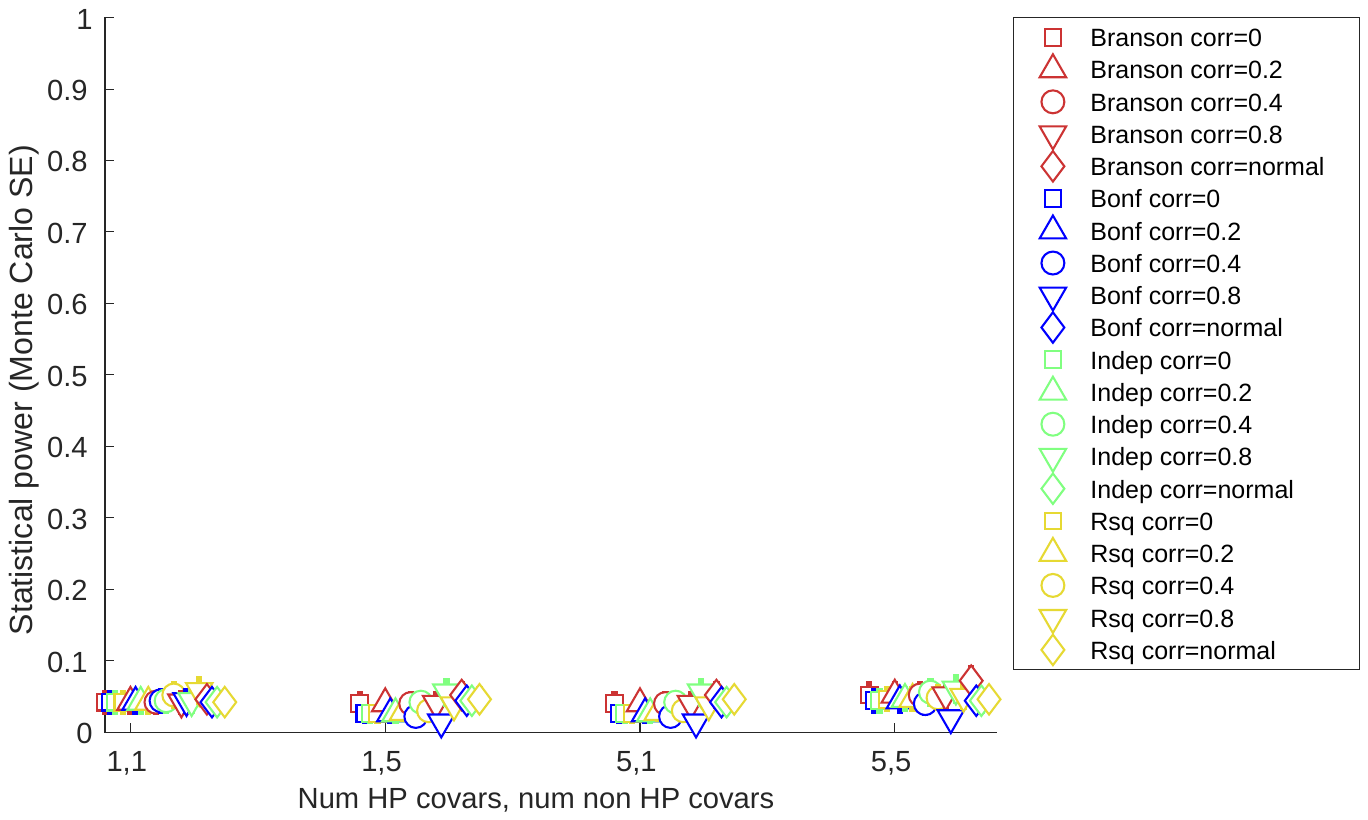 | 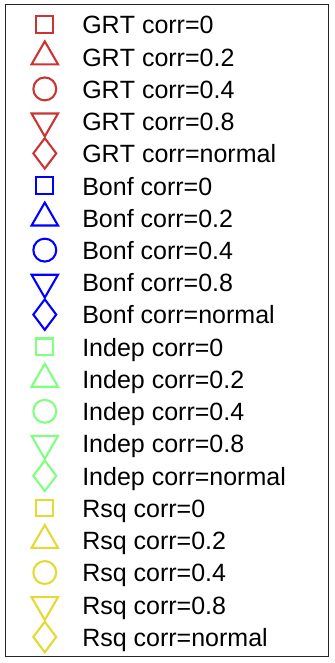 |

GRT: global randomization test; SE: standard error. Confidence intervals are +/- 1.96*MCSE (Monte Carlo standard error).

#### Supplementary figure 7: Results of horizontal pleiotropy applied example

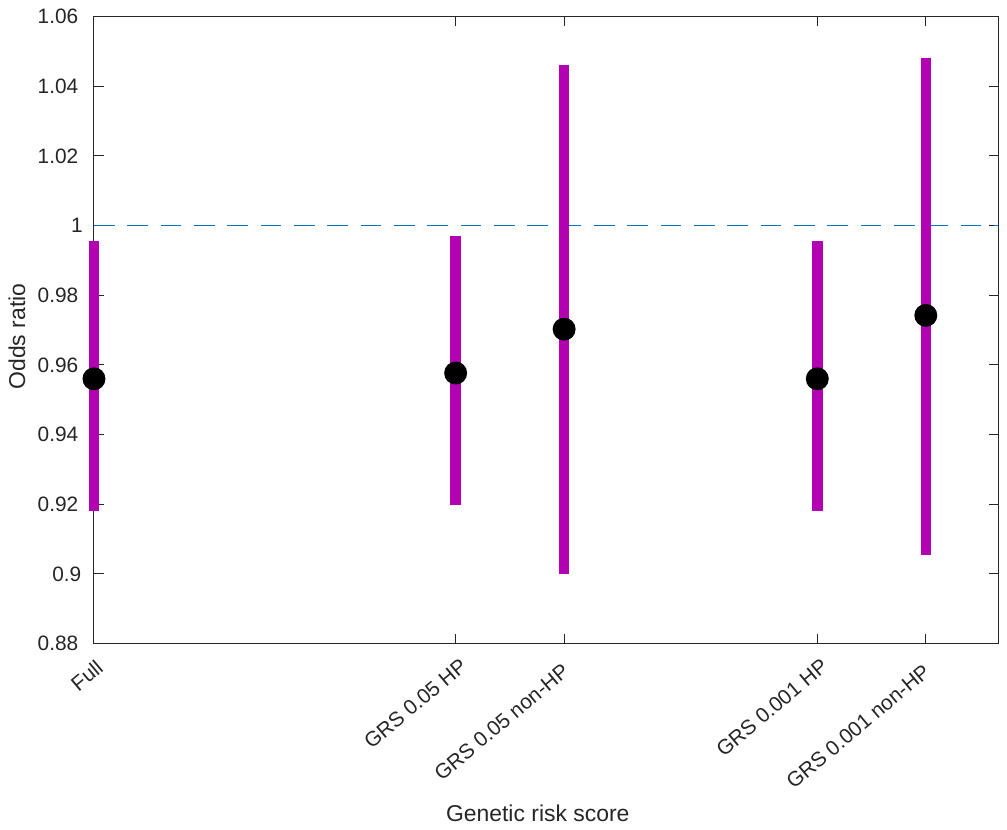

GRS: genetic risk score; HP: horizontal pleiotropy.

Full: GRS with all 58 CRP SNPs.

GRS 0.05 HP: GRS with the 51 SNPs found to be associated with the covariates set using a P=0.05 threshold.

GRS 0.05 HP: GRS with the 7 SNPs not found to be associated with the covariates set using a P=0.05 threshold.

GRS 0.001 HP: GRS with the 46 SNPs found to be associated with the covariates set using a P=0.001 threshold.

GRS 0.001 HP: GRS with the 12 SNPs not found to be associated with the covariates set using a P=0.001 threshold.

#### Supplementary figure 8: Relationship between number of tests and Bonferroni / independent test P value thresholds

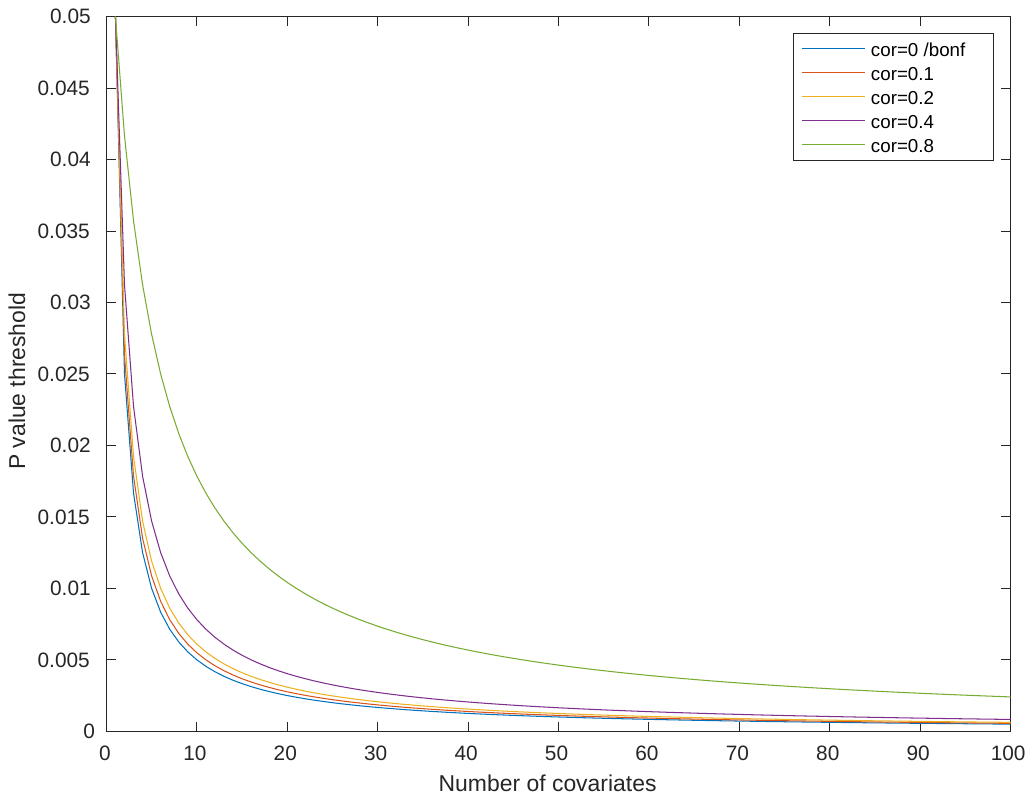

### SUPPLEMENTARY TABLES

#### Supplementary table 1: Correlations between covariates used to assess selection bias in MR applied analysis

|  | **Sex** | **Age** | **Height** | **Northing** | **Easting** | **Age left continuous full-time education** | **Townsend deprivation index** |
| --- | --- | --- | --- | --- | --- | --- | --- |
| **Sex** | 1 | 0.024 | 0.707 | 0.005 | -0.006 | 0.029 | 0.013 |
| **Age** |  | 1 | -0.107 | 0.022 | -0.002 | -0.187 | -0.091 |
| **Height** |  |  | 1 | -0.046 | 0.030 | 0.124 | -0.053 |
| **Northing** |  |  |  | 1 | -0.434 | -0.101 | -0.007 |
| **Easting** |  |  |  |  | 1 | 0.086 | 0.117 |
| **Age left continuous full-time education** |  |  |  |  |  | 1 | -0.072 |
| **Townsend** |  |  |  |  |  |  | 1 |

#### Supplementary table 2: Correlations between covariates used in horizontal pleiotropy applied example

|  | **P values** | | | | | | | | | | | | | |
| --- | --- | --- | --- | --- | --- | --- | --- | --- | --- | --- | --- | --- | --- | --- |
| **SNP** | **Smoking pack years** | **BMI** | **Weight** | **Leukocyte count** | **Albumin** | **Apol A** | **Apol B** | **Total chol** | **Glucose** | **HDL chol** | **Lipo A** | **SBP** | **DBP** | **Waist-hip ratio** |
| **Smoking pack years** | 1 | 0.146 | 0.152 | 0.176 | -0.078 | -0.123 | -0.019 | -0.081 | 0.090 | -0.150 | -0.006 | 0.091 | 0.024 | 0.263 |
| **BMI** |  | 1 | 0.834 | 0.155 | -0.147 | -0.271 | 0.079 | -0.049 | 0.160 | -0.347 | 0.009 | 0.188 | 0.276 | 0.434 |
| **Weight** |  |  | 1 | 0.101 | -0.068 | -0.380 | 0.044 | -0.116 | 0.142 | -0.449 | -0.002 | 0.173 | 0.284 | 0.598 |
| **Leukocyte count** |  |  |  | 1 | -0.040 | -0.099 | 0.015 | -0.040 | 0.047 | -0.140 | -0.009 | 0.063 | 0.050 | 0.140 |
| **Albumin** |  |  |  |  | 1 | 0.115 | 0.110 | 0.145 | -0.027 | 0.099 | -0.029 | 0.085 | 0.107 | 0.007 |
| **Apol A** |  |  |  |  |  | 1 | -0.027 | 0.319 | -0.064 | 0.919 | 0.007 | 0.029 | -0.035 | -0.392 |
| **Apol B** |  |  |  |  |  |  | 1 | 0.887 | -0.052 | -0.034 | 0.061 | 0.124 | 0.169 | 0.068 |
| **Total chol** |  |  |  |  |  |  |  | 1 | -0.093 | 0.335 | 0.054 | 0.105 | 0.131 | -0.110 |
| **Glucose** |  |  |  |  |  |  |  |  | 1 | -0.091 | -0.005 | 0.110 | 0.037 | 0.160 |
| **HDL chol** |  |  |  |  |  |  |  |  |  | 1 | 0.018 | -0.029 | -0.087 | -0.479 |
| **Lipo A** |  |  |  |  |  |  |  |  |  |  | 1 | 0.003 | 0.003 | -0.007 |
| **SBP** |  |  |  |  |  |  |  |  |  |  |  | 1 | 0.698 | 0.224 |
| **DBP** |  |  |  |  |  |  |  |  |  |  |  |  | 1 | 0.241 |
| **Waist-hip ratio** |  |  |  |  |  |  |  |  |  |  |  |  |  | 1 |

#### Supplementary table 3: P values for individual tests of covariates with SNPs for horizontal pleiotropy applied example

|  | **P values** | | | | | | | | | | | | | |
| --- | --- | --- | --- | --- | --- | --- | --- | --- | --- | --- | --- | --- | --- | --- |
| **SNP** | **Smoking pack years** | **BMI** | **weight** | **Leukocyte count** | **Albumin** | **Apol A** | **Apol B** | **Total chol** | **Glucose** | **HDL chol** | **Lipo A** | **SBP** | **DBP** | **Waist-hip ratio** |
| rs10512597 | 0.021 | 0.465 | 0.277 | <0.001 | 0.007 | 0.005 | 0.865 | 0.276 | 0.869 | 0.009 | 0.749 | 0.889 | 0.166 | 0.641 |
| rs1051338 | 0.885 | 0.410 | 0.753 | <0.001 | <0.001 | 0.025 | <0.001 | <0.001 | 0.022 | 0.070 | 0.157 | 0.655 | 0.165 | 0.572 |
| rs10521222 | 0.855 | 0.778 | 0.200 | 0.807 | 0.005 | 0.328 | 0.095 | 0.368 | 0.668 | 0.460 | 0.560 | 0.671 | 0.711 | 0.197 |
| rs10778215 | 0.897 | <0.001 | <0.001 | 0.625 | <0.001 | <0.001 | 0.462 | 0.943 | <0.001 | <0.001 | 0.128 | 0.882 | 0.084 | 0.012 |
| rs10832027 | 0.104 | <0.001 | 0.005 | 0.469 | <0.001 | 0.241 | 0.707 | 0.657 | 0.920 | 0.001 | 0.084 | <0.001 | <0.001 | <0.001 |
| rs10838687 | 0.841 | <0.001 | 0.149 | 0.022 | 0.001 | <0.001 | 0.094 | <0.001 | 0.001 | <0.001 | 0.905 | <0.001 | <0.001 | 0.978 |
| rs10925027 | 0.625 | 0.512 | 0.884 | <0.001 | <0.001 | 0.330 | 0.297 | 0.452 | 0.168 | 0.661 | 0.954 | 0.623 | 0.471 | 0.273 |
| rs11108056 | 0.826 | 0.080 | 0.867 | 0.002 | 0.437 | 0.910 | 0.774 | 0.432 | 0.156 | 0.912 | 0.164 | 0.838 | 0.121 | 0.102 |
| rs112635299 | 0.344 | <0.001 | <0.001 | 0.477 | <0.001 | 0.606 | 0.154 | 0.289 | 0.612 | 0.098 | 0.478 | <0.001 | <0.001 | 0.005 |
| rs1189402 | 0.645 | 0.016 | 0.033 | 0.835 | 0.004 | 0.145 | 0.001 | 0.015 | 0.864 | 0.359 | 0.510 | 0.143 | 0.571 | 0.101 |
| rs12202641 | 0.094 | 0.174 | 0.002 | 0.354 | <0.001 | <0.001 | <0.001 | <0.001 | 0.252 | <0.001 | 0.632 | 0.646 | <0.001 | 0.131 |
| rs1260326 | 0.806 | <0.001 | <0.001 | <0.001 | <0.001 | <0.001 | <0.001 | <0.001 | <0.001 | 0.124 | 0.237 | 0.255 | 0.394 | 0.346 |
| rs12960928 | 0.176 | <0.001 | <0.001 | 0.191 | 0.021 | <0.001 | 0.378 | 0.002 | 0.030 | <0.001 | 0.199 | 0.404 | 0.018 | <0.001 |
| rs12995480 | 0.003 | <0.001 | <0.001 | 0.028 | 0.018 | 0.014 | 0.615 | 0.717 | 0.144 | 0.007 | 0.338 | <0.001 | <0.001 | <0.001 |
| rs13233571 | 0.559 | <0.001 | <0.001 | <0.001 | <0.001 | 0.448 | <0.001 | 0.015 | 0.006 | <0.001 | 0.097 | 0.240 | 0.012 | 0.115 |
| rs13409371 | 0.895 | 0.369 | 0.461 | <0.001 | <0.001 | 0.020 | 0.002 | <0.001 | 0.880 | 0.453 | 0.244 | 0.165 | 0.386 | 0.311 |
| rs1441169 | 0.894 | 0.955 | 0.889 | 0.018 | 0.021 | 0.081 | 0.082 | 0.052 | 0.039 | 0.076 | 0.358 | 0.601 | 0.147 | 0.810 |
| rs1490384 | 0.107 | <0.001 | <0.001 | <0.001 | 0.753 | 0.860 | <0.001 | <0.001 | 0.003 | 0.027 | 0.914 | 0.720 | 0.015 | 0.006 |
| rs1514895 | 0.102 | <0.001 | <0.001 | 0.215 | 0.082 | 0.103 | 0.261 | 0.058 | <0.001 | 0.944 | 0.696 | 0.516 | 0.013 | 0.067 |
| rs1558902 | <0.001 | <0.001 | <0.001 | <0.001 | <0.001 | <0.001 | 0.004 | <0.001 | <0.001 | <0.001 | 0.336 | <0.001 | 0.002 | <0.001 |
| rs1582763 | 0.192 | 0.009 | 0.117 | <0.001 | 0.449 | 0.009 | 0.004 | 0.456 | 0.588 | 0.016 | 0.585 | 0.675 | 0.393 | 0.316 |
| rs1736060 | 0.690 | <0.001 | 0.002 | <0.001 | 0.566 | 0.007 | 0.022 | 0.592 | 0.864 | 0.449 | 0.281 | <0.001 | <0.001 | 0.005 |
| rs17658229 | 0.026 | 0.918 | 0.023 | 0.385 | 0.942 | 0.433 | 0.808 | 0.865 | 0.231 | 0.472 | 0.801 | 0.022 | <0.001 | 0.017 |
| rs178810 | 0.799 | <0.001 | <0.001 | <0.001 | 0.134 | 0.997 | 0.146 | 0.093 | 0.338 | 0.623 | 0.833 | 0.968 | 0.490 | 0.295 |
| rs1800961 | 0.111 | 0.893 | 0.316 | <0.001 | 0.019 | <0.001 | <0.001 | <0.001 | <0.001 | <0.001 | 0.388 | 0.114 | 0.784 | 0.0438 |
| rs1805096 | 0.462 | 0.459 | 0.813 | <0.001 | <0.001 | <0.001 | 0.593 | <0.001 | 0.530 | <0.001 | 0.236 | 0.783 | 0.898 | 0.120 |
| rs1880241 | 0.433 | 0.212 | 0.696 | <0.001 | <0.001 | 0.161 | 0.703 | 0.080 | 0.262 | 0.243 | 0.169 | <0.001 | 0.126 | 0.402 |
| rs2064009 | 0.484 | 0.001 | 0.002 | 0.300 | 0.012 | 0.002 | 0.088 | 0.588 | 0.061 | 0.014 | 0.153 | 0.313 | 0.164 | 0.028 |
| rs2239222 | 0.337 | 0.525 | 0.513 | 0.620 | 0.007 | 0.495 | 0.762 | 0.955 | 0.392 | 0.496 | 0.305 | 0.914 | 0.051 | 0.177 |
| rs2293476 | 0.917 | <0.001 | <0.001 | 0.290 | <0.001 | <0.001 | 0.203 | <0.001 | 0.085 | <0.001 | 0.190 | 0.888 | <0.001 | <0.001 |
| rs2315008 | 0.010 | 0.002 | <0.001 | <0.001 | 0.014 | <0.001 | 0.037 | 0.001 | 0.098 | <0.001 | 0.484 | 0.080 | <0.001 | 0.120 |
| rs2352975 | 0.559 | <0.001 | <0.001 | <0.001 | 0.994 | <0.001 | 0.013 | <0.001 | 0.021 | <0.001 | 0.256 | <0.001 | <0.001 | <0.001 |
| rs2710804 | 0.190 | 0.326 | 0.898 | <0.001 | <0.001 | 0.271 | 0.144 | 0.038 | 0.023 | 0.280 | 0.773 | 0.003 | 0.029 | 0.524 |
| rs2794520 | 0.818 | 0.315 | 0.488 | 0.674 | 0.393 | 0.540 | 0.014 | 0.007 | 0.346 | 0.376 | 0.969 | 0.402 | 0.518 | 0.929 |
| rs2836878 | 0.559 | 0.550 | 0.773 | <0.001 | <0.001 | 0.055 | 0.098 | 0.006 | 0.289 | 0.157 | 0.346 | 0.344 | 0.429 | 0.959 |
| rs2852151 | 0.880 | 0.137 | 0.290 | 0.018 | 0.895 | 0.626 | 0.025 | 0.166 | 0.953 | 0.326 | 0.769 | 0.051 | 0.212 | 0.479 |
| rs2891677 | 0.030 | 0.003 | 0.381 | <0.001 | 0.701 | 0.747 | 0.839 | 0.990 | 0.426 | 0.412 | 0.795 | 0.895 | 0.758 | 0.263 |
| rs340005 | 0.278 | <0.001 | <0.001 | 0.046 | <0.001 | <0.001 | 0.005 | 0.531 | <0.001 | 0.021 | 0.755 | 0.006 | 0.022 | 0.844 |
| rs4092465 | 0.947 | 0.580 | 0.224 | 0.175 | 0.251 | 0.251 | 0.423 | 0.597 | 0.966 | 0.504 | 0.346 | 0.453 | 0.312 | 0.106 |
| rs4129267 | 0.586 | 0.932 | 0.539 | 0.223 | <0.001 | <0.001 | 0.175 | <0.001 | 0.180 | 0.391 | 0.240 | 0.160 | 0.827 | 0.455 |
| rs4246598 | 0.864 | 0.103 | 0.056 | 0.743 | 0.325 | 0.169 | 0.001 | 0.005 | 0.095 | 0.699 | 0.774 | 0.796 | 0.106 | 0.846 |
| rs4420638 | 0.091 | <0.001 | <0.001 | <0.001 | 0.730 | <0.001 | <0.001 | <0.001 | 0.001 | <0.001 | 0.236 | 0.406 | 0.388 | <0.001 |
| rs469772 | 0.125 | 0.207 | 0.280 | 0.640 | 0.141 | 0.009 | 0.001 | <0.001 | 0.141 | 0.993 | 0.253 | 0.177 | 0.174 | 0.142 |
| rs4774590 | 0.164 | <0.001 | <0.001 | 0.108 | 0.273 | 0.385 | 0.100 | 0.118 | 0.018 | 0.277 | 0.279 | 0.666 | 0.216 | 0.017 |
| rs4841132 | 0.660 | 0.701 | 0.946 | 0.028 | <0.001 | <0.001 | <0.001 | <0.001 | <0.001 | <0.001 | 0.984 | 0.308 | 0.004 | 0.967 |
| rs6001193 | 0.255 | 0.393 | 0.169 | 0.160 | 0.107 | <0.001 | 0.125 | 0.751 | 0.671 | <0.001 | 0.011 | 0.268 | 0.037 | 0.010 |
| rs643434 | 0.008 | 0.947 | 0.823 | <0.001 | 0.002 | <0.001 | <0.001 | <0.001 | <0.001 | <0.001 | 0.004 | 0.637 | <0.001 | 0.780 |
| rs687339 | 0.010 | <0.001 | <0.001 | 0.113 | <0.001 | <0.001 | <0.001 | 0.221 | 0.012 | <0.001 | 0.007 | <0.001 | 0.011 | 0.001 |
| rs7121935 | 0.159 | 0.121 | 0.997 | 0.876 | <0.001 | 0.033 | 0.497 | 0.628 | 0.122 | 0.486 | 0.423 | 0.638 | 0.028 | 0.263 |
| rs7310409 | 0.371 | 0.819 | 0.342 | 0.007 | 0.003 | <0.001 | <0.001 | <0.001 | 0.765 | <0.001 | 0.630 | 0.007 | 0.004 | 0.541 |
| rs75460349 | 0.848 | 0.007 | 0.569 | 0.021 | 0.276 | <0.001 | <0.001 | <0.001 | 0.830 | <0.001 | 0.473 | 0.019 | 0.015 | 0.454 |
| rs7795281 | 0.056 | <0.001 | 0.002 | 0.424 | 0.980 | 0.593 | 0.379 | 0.179 | 0.024 | 0.734 | 0.708 | 0.529 | <0.001 | 0.067 |
| rs9271608 | 0.331 | 0.060 | 0.609 | <0.001 | 0.346 | <0.001 | 0.003 | 0.168 | <0.001 | 0.001 | 0.354 | 0.473 | <0.001 | <0.001 |
| rs9284725 | 0.985 | 0.795 | 0.119 | 0.185 | 0.045 | 0.146 | 0.473 | 0.444 | 0.366 | 0.523 | 0.205 | 0.139 | 0.340 | 0.087 |
| rs9385532 | 0.218 | <0.001 | <0.001 | <0.001 | 0.203 | 0.012 | <0.001 | <0.001 | 0.635 | 0.011 | 0.803 | 0.071 | 0.363 | 0.008 |
| rs9611441 | 0.195 | 0.183 | 0.265 | 0.635 | 0.150 | 0.544 | <0.001 | <0.001 | 0.166 | 0.646 | 0.351 | 0.072 | 0.445 | <0.001 |
| X17.58001690_GA_G | 0.085 | 0.175 | 0.318 | <0.001 | <0.001 | 0.047 | 0.206 | 0.148 | 0.134 | 0.617 | 0.539 | <0.001 | 0.006 | 0.550 |
| X3.47431869_GTCT_G | 0.126 | 0.057 | 0.386 | <0.001 | 0.014 | <0.001 | 0.384 | 0.144 | 0.252 | <0.001 | 0.433 | 0.691 | 0.412 | 0.005 |

#### Supplementary table 4: Results of horizontal pleiotropy applied example – identifying horizontally pleiotropic CRP SNPs

|  | **P value** | | | |
| --- | --- | --- | --- | --- |
| **SNP** | **Global Randomization test** | **R^2^ permutation test (r^2^perm)** | **Bonferroni corrected (test-Bonf) (14 tests) *** | **Independent (test-indep) (12.0 tests) *** |
| rs10512597 | <0.001 | <0.001 | <0.001 | <0.001 |
| rs1051338 | <0.001 | 0.012 | <0.001 | <0.001 |
| rs10521222 | **0.176** | **0.144** | **0.072** | **0.062** |
| rs10778215 | <0.001 | <0.001 | <0.001 | <0.001 |
| rs10832027 | <0.001 | 0.007 | <0.001 | <0.001 |
| rs10838687 | <0.001 | <0.001 | <0.001 | <0.001 |
| rs10925027 | <0.001 | <0.001 | <0.001 | <0.001 |
| rs11108056 | 0.015 | **0.108** | 0.023 | 0.020 |
| rs112635299 | <0.001 | <0.001 | <0.001 | <0.001 |
| rs1189402 | 0.040 | **0.101** | 0.020 | 0.017 |
| **rs**12202641 | <0.001 | <0.001 | <0.001 | <0.001 |
| rs1260326 | <0.001 | <0.001 | <0.001 | <0.001 |
| rs12960928 | <0.001 | <0.001 | <0.001 | <0.001 |
| rs12995480 | <0.001 | <0.001 | <0.001 | <0.001 |
| rs13233571 | <0.001 | <0.001 | <0.001 | <0.001 |
| rs13409371 | <0.001 | <0.001 | <0.001 | <0.001 |
| rs1441169 | 0.006 | **0.321** | **0.256** | **0.219** |
| rs1490384 | <0.001 | <0.001 | <0.001 | <0.001 |
| rs1514895 | <0.001 | <0.001 | <0.001 | <0.001 |
| rs1558902 | <0.001 | <0.001 | <0.001 | <0.001 |
| rs1582763 | 0.006 | 0.032 | <0.001 | <0.001 |
| rs1736060 | <0.001 | 0.005 | <0.001 | <0.001 |
| rs17658229 | <0.001 | 0.012 | <0.001 | <0.001 |
| rs178810 | <0.001 | 0.009 | <0.001 | <0.001 |
| rs1800961 | <0.001 | <0.001 | <0.001 | <0.001 |
| rs1805096 | <0.001 | <0.001 | <0.001 | <0.001 |
| rs1880241 | <0.001 | <0.001 | <0.001 | <0.001 |
| rs2064009 | <0.001 | **0.076** | 0.009 | 0.008 |
| rs2239222 | **0.161** | **0.171** | **0.096** | **0.083** |
| rs2293476 | <0.001 | <0.001 | <0.001 | <0.001 |
| rs2315008 | <0.001 | <0.001 | <0.001 | <0.001 |
| rs2352975 | <0.001 | <0.001 | <0.001 | <0.001 |
| rs2710804 | <0.001 | <0.001 | <0.001 | <0.001 |
| rs2794520 | **0.847** | **0.212** | **0.097** | **0.083** |
| rs2836878 | <0.001 | <0.001 | <0.001 | <0.001 |
| rs2852151 | **0.551** | **0.360** | **0.250** | **0.214** |
| rs2891677 | <0.001 | 0.002 | <0.001 | <0.001 |
| rs340005 | <0.001 | <0.001 | <0.001 | <0.001 |
| rs4092465 | **0.311** | **0.827** | **1.000** | **1.000** |
| rs4129267 | <0.001 | <0.001 | <0.001 | <0.001 |
| rs4246598 | 0.029 | **0.095** | 0.021 | 0.018 |
| rs4420638 | <0.001 | <0.001 | <0.001 | <0.001 |
| rs469772 | <0.001 | 0.017 | <0.001 | <0.001 |
| rs4774590 | **0.055** | **0.061** | 0.005 | 0.004 |
| rs4841132 | <0.001 | <0.001 | <0.001 | <0.001 |
| rs6001193 | <0.001 | 0.001 | <0.001 | <0.001 |
| rs643434 | <0.001 | <0.001 | <0.001 | <0.001 |
| rs687339 | <0.001 | <0.001 | <0.001 | <0.001 |
| rs7121935 | <0.001 | 0.002 | <0.001 | <0.001 |
| rs7310409 | <0.001 | <0.001 | <0.001 | <0.001 |
| rs75460349 | <0.001 | <0.001 | <0.001 | <0.001 |
| rs7795281 | <0.001 | <0.001 | <0.001 | <0.001 |
| rs9271608 | <0.001 | <0.001 | <0.001 | <0.001 |
| rs9284725 | **0.153** | **0.500** | **0.631** | **0.541** |
| rs9385532 | <0.001 | <0.001 | <0.001 | <0.001 |
| rs9611441 | <0.001 | 0.003 | <0.001 | <0.001 |
| X17.58001690_GA_G | <0.001 | <0.001 | <0.001 | <0.001 |
| X3.47431869_GTCT_G | <0.001 | <0.001 | <0.001 | <0.001 |

P values shown in bold are those >0.05, such that the SNP is not identified as being horizontally pleiotropic.
